# Field assessment of the burden and determinants of malaria transmission to inform tailoring of interventions (microstratification) in Ibadan and Kano metropolis: Study protocol

**DOI:** 10.1101/2023.01.20.23284766

**Authors:** Ifeoma D. Ozodiegwu, Akintayo O. Ogunwale, Olabanji Surakat, Joshua O. Akinyemi, Eniola Bamgboye, Adeniyi F. Fagbamigbe, Musa Muhammad Bello, Al-Mukhtar Y. Adamu, Perpetua Uhomobhi, Cyril Ademu, Chukwu Okoronkwo, Monsuru Adeleke, IkeOluwapo Ajayi

## Abstract

**Introduction:** Rapid urbanization in Nigerian cities may likely result in small-scale variation in malaria transmission with a higher malaria burden in informal settlements and slums. However, data is unavailable to quantify city-level variations in transmission risk and to inform selection of appropriate interventions. We are conducting field studies to understand how malaria risk varies at the smallest administrative units (wards) in two major Nigerian cities – Ibadan and Kano – to inform tailoring of interventions for cities in Nigeria’s National Malaria Strategic Plan.

**Methods and Analysis:** This is a mixed-method research design involving qualitative and quantitative research methods. We will carry out formative qualitative studies to inform sampling strategies for formal settlements, informal settlements and slums and the design of survey questionnaires. Cross-sectional surveys will be conducted in the wet and dry seasons across wards in the study cities and within different settlement types at the household and health facility levels to map all-age and under-five malaria prevalence and evaluate related risk factors. A 12-month longitudinal study among children under the age of 10 years will provide information on malaria seasonality in the two cities and related factors. Entomological surveys will be used to undertaken for eight months (four months in each season) per city to assess local transmission risk in both cities. Descriptive and regression analysis will be applied to study data to assess malaria prevalence, seasonality and related factors. Mathematical models of malaria transmission for each study ward will be developed based on study data to inform assessment of the impact of various intervention scenarios specified by the NMEP on malaria transmission.

**Ethics and Dissemination:** This protocol has been approved by Nigeria’s National Health Research Committee and relevant institutional Ethical Review Committees. Study results will be disseminated to key stakeholders and study communities.

**Strength and Limitations:** *Strengths:*

- Cross-sectional and longitudinal study sampling procedures and questionnaire design are enhanced by a strong formative research foundation
- Study design and site selection are informed by modeling and local knowledge
- The combination of human and vector studies will provide detailed information on malaria transmission risk and associated drivers

*Limitations:*

- Cross-sectional study may not be powered to measure all-age prevalence as sample size estimation is based on prevalence among children under the age of five years
- Study findings may not adequately measure transmission burden and risk factors in unsampled wards

## Introduction

Nigeria, in addition to being the greatest contributor to the global malaria burden, is rapidly urbanizing with the attendant challenges of overcrowding and environmental degradation, leading to growing concerns that malaria transmission may increase substantially in cities [1,2] Presently, a little over half of Nigerians reside in urban centers and it is projected that this number will rise to 70 percent in 2050 [3]. Infrastructural elements due to urban development are expected to reduce malaria transmission [4]. However, non-uniform infrastructure planning and provision within city neighborhoods is resulting in the expansion of informal settlements, slums, and urban farms with suitable habitats for vector breeding [5,6]. In addition, the discovery of *Anopheles Stephensi*, a malaria vector adapted to urban areas, in Nigeria may intensify transmission risk for urban residents [2,7–9]. Neighborhood-level differences in environmental suitability for mosquito breeding in cities will likely lead to small-scale geographic variation in malaria burden and these need to be accounted for during intervention planning.

Recognition of the implications of the spatial variation in malaria risk underlies the High Burden to High Impact (HBHI) response launched by the World Health Organization (WHO) in 2018 to reignite the pace towards achieving the Global Technical Strategy (GTS) targets of reducing malaria cases and deaths by 75 and 90 percent in 2025 and 2030 relative to 2015 levels [10,11]. High-burden malaria countries like Nigeria where declines in malaria burden have stagnated in recent years constitute a priority for HBHI-related activities. The second element of the HBHI response calls for high-burden countries to transition from a ‘one-size-fits-all’ approach to a more targeted response to enable the deployment of the most effective malaria control tools to areas where they will have maximum impact. The Nigerian Malaria Elimination Programme (NMEP) heeded this call and, in the 2021 – 2025 National Malaria Strategic Plan, interventions were tailored for each Local Government Area (LGA). Intervention tailoring is also needed within cities, particularly targeting informal settlements, slums and neighborhoods situated close to farms as residents may be at higher malaria risk as compared to planned settlements with high-quality housing and improved environmental conditions. WHO has recommendations for targeting malaria interventions to individual geographic areas taking into account local epidemiology, ecology, health system and socio-economic characteristics and these can be used to inform intervention decisions [12]. Selected intervention scenarios per locality can then be evaluated with mathematical models to understand their impact and inform the development of city-level intervention plans within national malaria strategic plans. However, data is lacking to facilitate these activities.

City-level variations in malaria burden and risk factors are not well-understood in Nigeria. National surveys do not have sufficient sample sizes necessary for capturing intra-urban malaria risk. While clinical malaria data encompasses malaria incidence in individuals living in cities, it is biased towards those that use public health facilities; socio-economic data, human behavior, information on place of residence and travel histories and frequency are typically not collected routinely; and it often lacks a well-defined catchment population [13]. These shortcomings of clinical malaria data preclude its use in understanding spatial distribution of malaria cases, deaths and risk factors in cities. In addition, data from entomological surveys are usually not specific to urban geographies [14]. Existing research also provides limited information on the burden and drivers of malaria risk in cities [15–19] These data and information gaps motivate the need for additional studies that elucidate the burden and drivers of malaria risk in urban Nigeria to inform city-level targeting of interventions.

### Conceptualizing the geographic scale of intervention targeting within cities

In urban areas, heterogeneity in epidemiological, entomological, health system and socio-economic determinants of malaria morbidity and mortality occur at fine spatial scales, such as neighborhoods, sub-districts, or wards, much smaller than the typical operational units of intervention targeting, states or local government areas. A cross-sectional survey of communities in Accra found that those located near urban agriculture had a higher prevalence of malaria [20]. Predictions of malaria parasite prevalence at a 10-meter grid level for the city of Dar es Salam found that higher malaria transmission risk occured along water channels and dense vegetation [21]. However, it is often not feasible for malaria control programs to identify non-administratively defined units for intervention targeting.

In Nigeria, wards are administratively defined units for political purposes, which may be sufficiently small enough to capture spatial variation in urban malaria risk and are easily identifiable by the state malaria programs during intervention planning and implementation. Therefore, this study will be conducted at the ward level to facilitate urban microstratification.

### Conceptualizing informal settlements and slums

Spatial variations in urban malaria risk emphasize the need to delineate the features of physical spaces that contribute to malaria transmission. Informal settlements or slums bear the highest malaria burden when compared to other urban localities [22]. The extant literature suggests that environmental conditions, human mobility patterns, and inadequate access to care in informal settlements and slums drive local transmission and facilitate the importation and adaptation of parasites to cities [22–28]. A clear definition of informal settlements and slums is critical to inform sampling strategies to adequately capture heterogeneities in urban malaria risk. While several definitions of informal settlements and slums abound [29–32], the definition by the United Nations (UN) Human Settlements Programme appear to be the most widely used [29,31,33,34]. According to the UN’s definition, informal settlements “are residential areas where 1) inhabitants have no security of tenure vis-à-vis the land or dwellings they inhabit, with modalities ranging from squatting to informal rental housing, 2) the neighborhoods usually lack, or are cut off from, basic services and city infrastructure and 3) the housing may not comply with current planning and building regulations and is often situated in geographically and environmentally hazardous areas” [34]. Slums are regarded as the ‘most deprived and excluded form of informal settlements [34] and more elaborate definitions by the UN emphasize the location of slums near or on most hazardous sites, and the issue of overcrowding within slum households [33]. What is clear from the UN’s definitions and others is that definitions of informal settlements and slums should be viewed from a perspective of human rights, basic infrastructure, structural quality of housing, and location.

Studies of areas identified as slums in Nigeria depict conditions that correspond to the UN’s definition of informal settlements and slums. A 2013 study of living conditions in Ajegunle, Ijora Oloye and Makoko, some of the largest slums in Lagos metropolis found that a third of residents resided in overcrowded households consisting of an average of five persons per room and the majority lacked access to basic amenities such as piped water, electricity supply and flush toilets [35]. Similarly, a study of slum conditions in Akure, the capital of Ondo state revealed poor housing and sanitary conditions and inadequate access to health facilities [35]. While these findings support the use of the UN’s definition of informal settlements and slums to guide the selection of study sites, local understanding of the features of informal settlements and slums may reveal additional information outside the indicators provided by the UN definition, hence, this study will rely on the opinions of key community stakeholders and professional experts to inform definitions of formal and informal settlements and slums.

### Objectives

To understand how to better tailor interventions in urban areas, we are collaborating with the NMEP to conduct human and entomological studies and use the corresponding data to construct a mathematical model of urban malaria transmission for Ibadan and Kano to inform the 2026 – 2030 National Malaria Strategic Plans. These cities were chosen due to their high malaria burden (at the state-level) and the potential cost-savings to the NMEP that would result from reducing malaria transmission. Data collected from these field studies will provide insight into how to design and implement high-impact intervention-tailoring approaches in other urban settings across Nigeria. Our human studies will involve qualitative, cross-sectional and longitudinal approaches while entomological studies will be strictly longitudinal. All study types will be conducted in both Ibadan and Kano. The overarching aims of the study are to evaluate the following: 1) ward-level prevalence of malaria, 2) malaria seasonality, 3) risk factors for malaria prevalence and incidence, and 4) risk of local transmission as assessed by entomological indicators. Qualitative assessments will be used to define formal, and informal settlements and slums in study wards, and to inform survey development and study planning for both the cross-sectional and longitudinal studies. Cross-sectional studies conducted at the household-level and health facilities will provide outputs for the estimation of ward-level malaria prevalence and for identification of associated key risk factors. Longitudinal studies will inform the estimation of malaria seasonality and identification of risk factors for formal, and informal settlements and slums. Entomological studies will inform the understanding of vector species composition, larval habitats and transmission rates (Figure 1).

**Figure 1:**
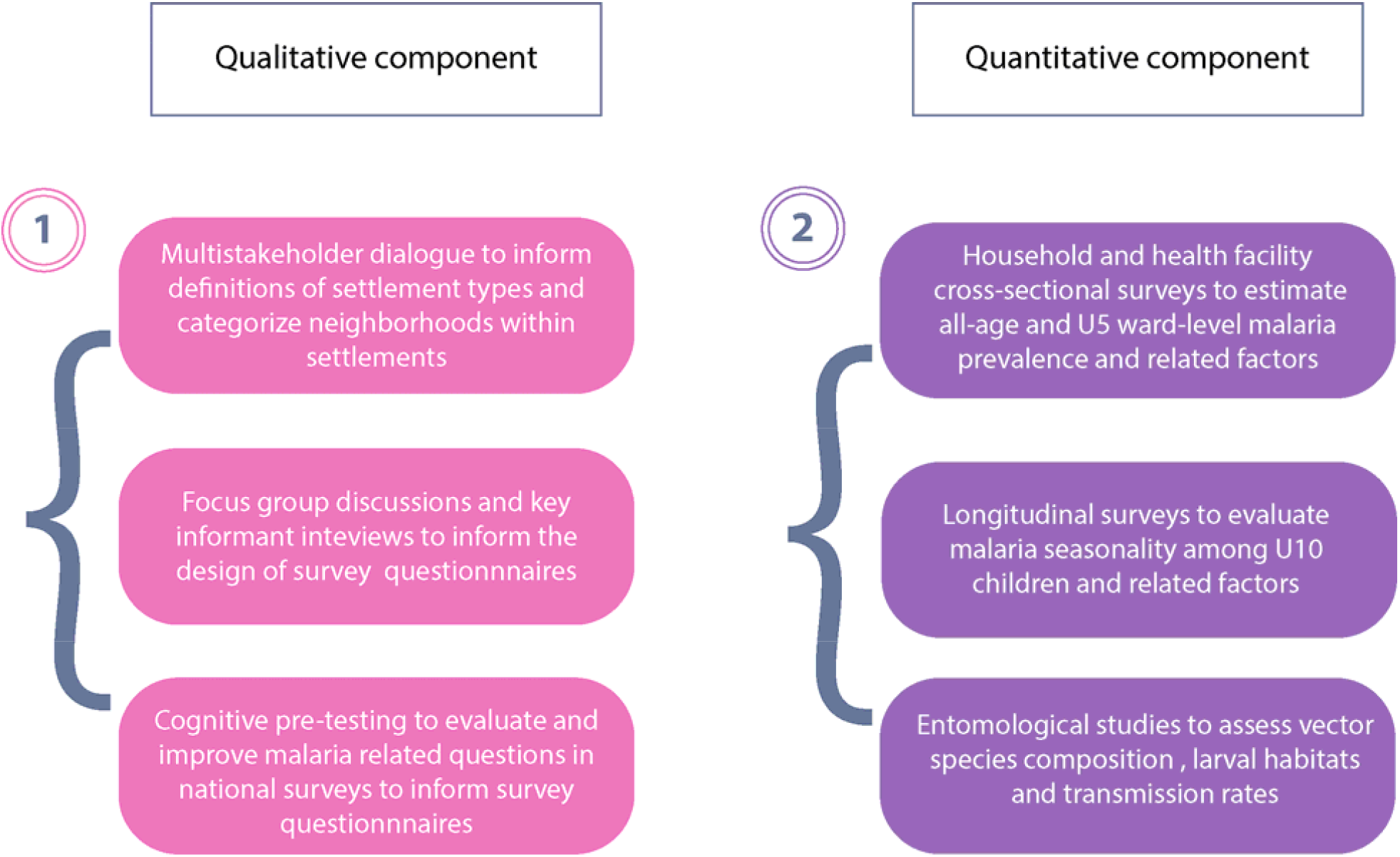
Study approach, types, and objectives. U5 is abbreviation for children under the age of five years and U10 means children under the age of 10 years

Data from these field studies will be used for calibration of a ward-level mathematical model for projecting the impact of various intervention scenarios on malaria burden indicators for individual wards in Kano and Ibadan metro areas. This will aid the NMEP to have a clear understanding of the implications of different intervention scenarios including the impact of deprioritizing vector control tools in city wards with low transmission burden. Our mathematical modeling results will support the NMEP in identifying optimal strategies for achieving malaria control and elimination goals.

## Methods

### Description of study location and study ward selection process for the prevalence, longitudinal and entomological surveys

Ibadan and Kano metro areas are the study locations. Ibadan metro area is located in Oyo state and has five LGAs further subdivided administratively into 59 wards (Figure 2A). Ward population sizes for Ibadan are presented in Figure 2B. The mean population size across all 59 wards in Ibadan is 15,408 (standard deviation (SD) – 12,747) [36] mean population density is 12,665 individuals per square kilometer (SD – 5,802) [37]. Kano metro area is located in Kano state and comprises six LGAs, which are subdivided administratively into 66 wards (Figure 3A). Ward population sizes for Kan are depicted in Figure 3B. The mean population size across all 66 wards in Kano is 47,497 (SD – 58,981) [36] and mean population density is 23,401 individuals per square kilometer (SD – 17, 313) [37].

**Figure 2:**
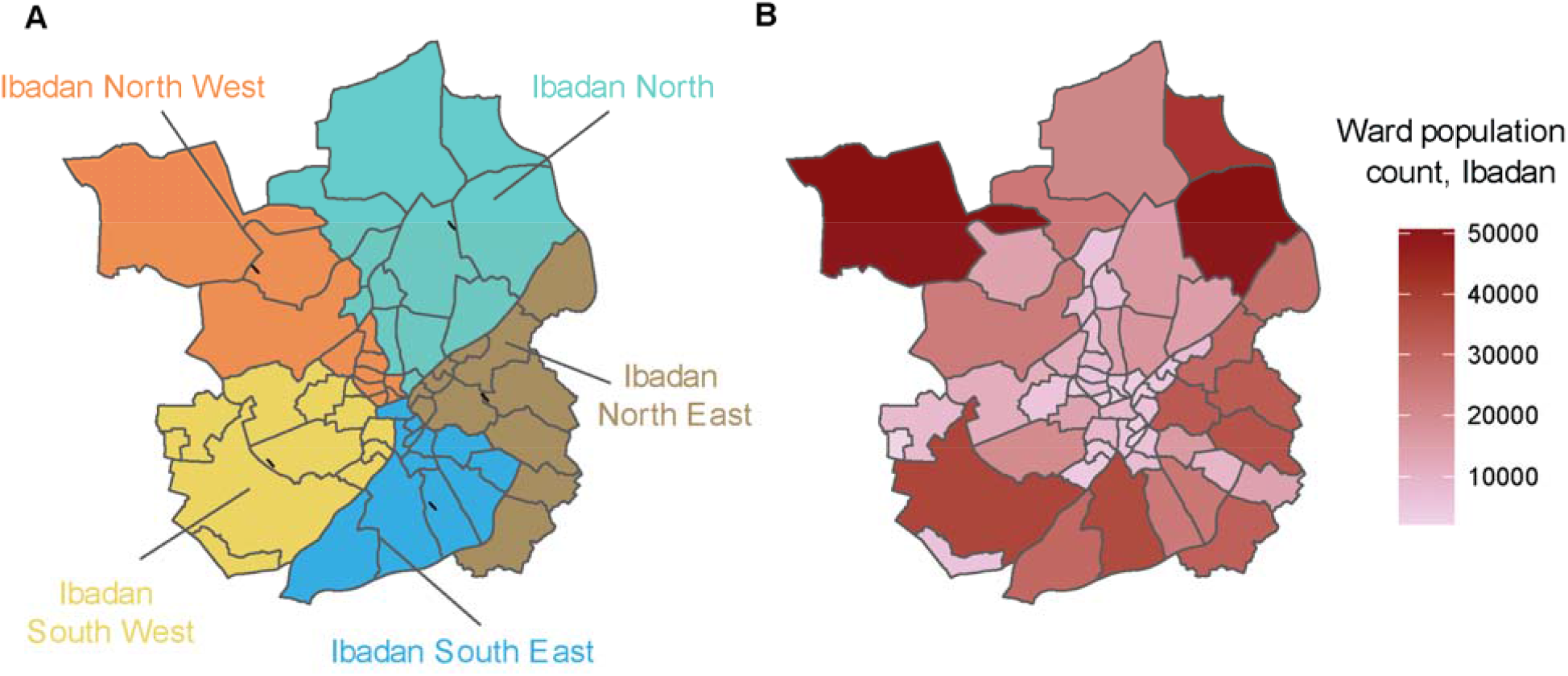
Administrative subdivisions of Ibadan metro. A) Ward boundaries are colored by their corresponding LGAs, B) Ward population count

**Figure 3:**
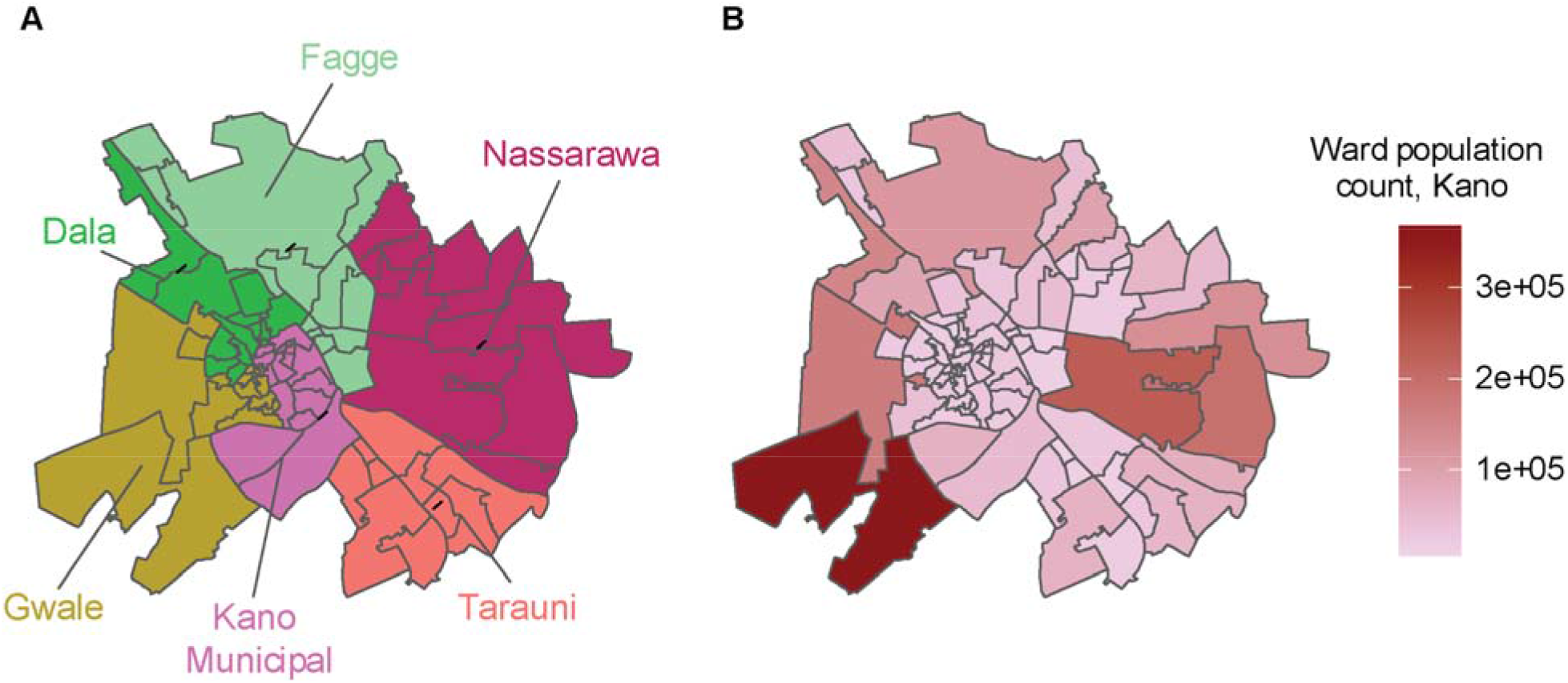
Administrative subdivisions of Kano metro. A) Ward boundaries are colored by their LGA, B) Wards in Kano population count

### Overview of the study ward selection process

Given the large number of wards in LGA in Ibadan and Kano metro, we developed a process to choose a selection of representative wards from which we can extrapolate prevalence, seasonality, and entomological data to unsampled wards. Model-based clustering was used to classify wards in Ibadan and Kano metropolis into groups with similar characteristics. Site visits were conducted for at least one ward per cluster to confirm the clustering results and inform the final ward selection. Figure 4 depicts the ward selection process for Ibadan. A similar figure for Kano is available in the Supplement.

**Figure 4:**
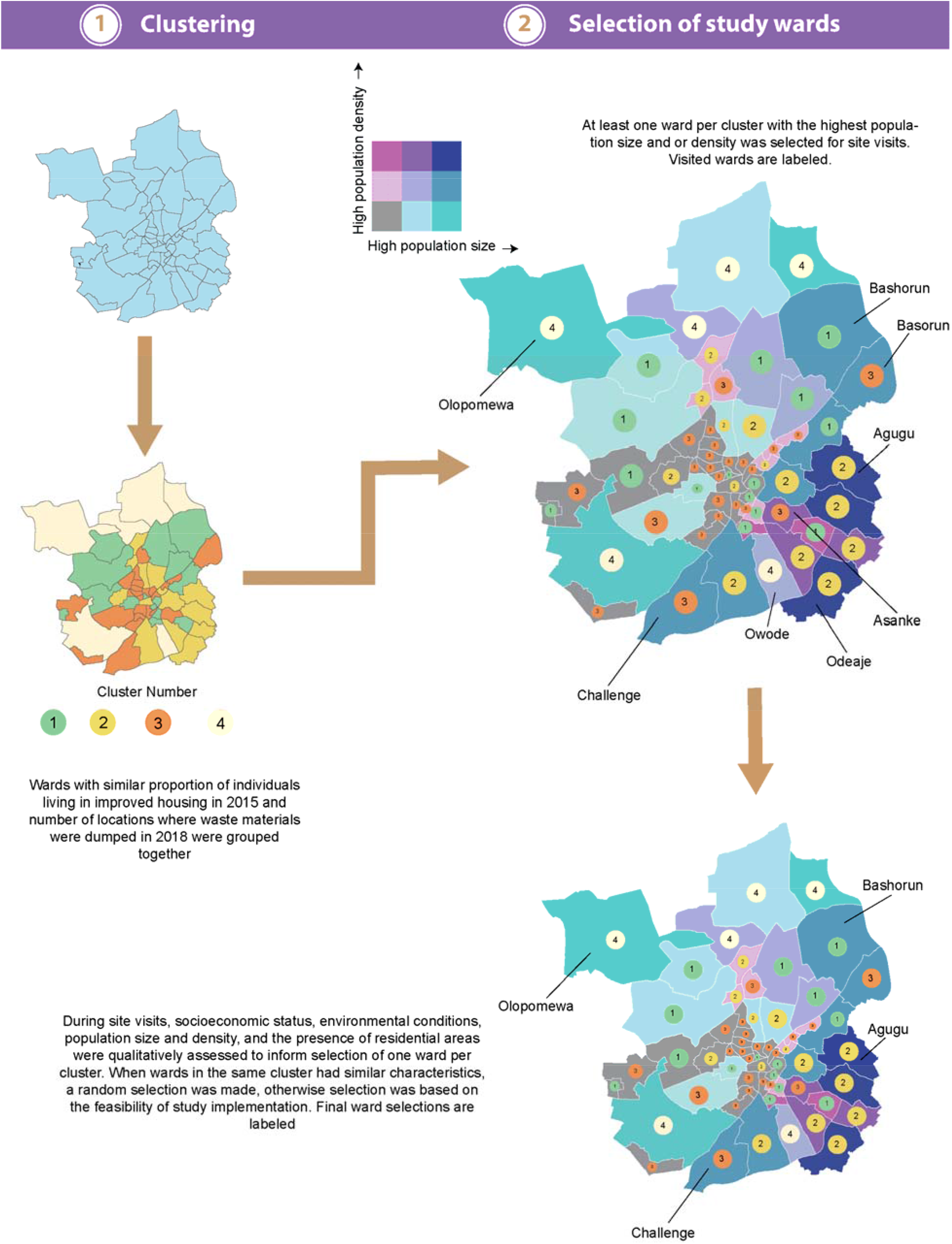
Overview of methods used to select study locations in Ibadan metropolis for the household, longitudinal and entomological surveys. Wards visited for ground truthing and final ward selections are labeled. Similar methods were used for ward selection in Kano metropolis

### Clustering

Estimates of malaria prevalence and seasonality and entomological parameters for each ward are essential for mathematical models to capture small-scale spatial variations in malaria transmission to provide projections of their impact, given an intervention scenario. However, generating such measures of transmission would be infeasible given limited resources. Clustering methods can be used to identify wards with similar socioeconomic and environmental characteristics to inform the extrapolation of transmission metrics from sampled to unsampled areas. We applied model-based clustering with variable selection as an empirical approach to classify wards into clusters [38]. Intial variables considered for clustering in Ibadan and Kano were ward-level data on population density [39], mean k-complexity (a measure of settlement informality) [40], enhanced vegetation index [41], measures of settlement type classification (received through email communication from Gates Foundation GIS Team), and proportion of individuals living in improved housing in 2015 [42]. An additional variable included in the clustering model solely for Ibadan was the number of locations where waste materials were dumped in 2018 [43]. After variable selection, data on the proportion of individuals living in improved housing in 2015 [42] and the number of locations where waste materials were dumped in 2018, was used to group wards in the Ibadan metro are into four clusters. Similarly, wards in the Kano metro area were aggregated into five clusters using data on the proportion of individuals living in improved housing in 2015 [42]. The selection of cluster partitions were based on Bayesian Information Criteria (BIC) values, that is those with the lowest BIC values were selected. Following the clustering procedure, one to three potential study locations, with either the largest population size and/or population density, per cluster were selected for site visits to inform decisions on the final selection of one study location per cluster (Table 1). Where one ward had both the highest population size and density, the ward with the next highest population size was chosen. Population size and density were chosen as the selection criteria for study wards on the assumption that selected areas will comprise individuals of different socioeconomic status and this diversity will result in the generation of representative estimates of transmission within their cluster partition.

**Table 1:** Potential and final study wards

| Cluster number | Ibadan metro area* | Kano metro area* |
| --- | --- | --- |
| 1 | Owode, **Olopomewa** | **Dorayi**, Goron Dutse |
| 2 | **Challenge**, Basorun, Asanke | **Fagge D2**, Kwachiri |
| 3 | Ode Aje, **Agugu** | **Tudun Wazurchi**, Sani Mainagge |
| 4 | **Bashorun** | **Gobirawa**, Kofar Ruwa |
| 5 |  | **Zango**, Shahuchi |

\*final study wards are in bold

### Site visits

Due to funding limitations, only one ward could be selected per cluster. Therefore, the study team conducted site visits to confirm the clustering results and inform final ward selections by assessing whether selected wards had residential areas and qualitatively comparing location characteristics within clusters where two or more potential study wards were selected during the clustering procedure. Eight potential study wards (Figure 4 and Table 1) were visited in the Ibadan metro area and ten wards were visited in the Kano metro area. The study team captured pictures of residential houses and roads in each area, and qualitatively assessed and compared socioeconomic status, environmental conditions, and population size and density for wards in the same cluster, where two or more wards were selected. Reports from the site visits in the Ibadan metro area, for instance, suggested that wards in clusters 2 were similar with respect to housing quality and road and neighborhood infrastructure while wards grouped into clusters 1 in Ibadan metro area were noted to be vastly different. To elaborate, most neighborhoods in Olopemewa had tarred roads in contrast to Owode. For Kano, clustering methods captured similarities in housing quality in wards in cluster 1 but not in cluster 3-housing infrastructure in Dorayi and Goron Dutse were similar but in Tudun Wazurchi, they were of lower quality than those in Sani Mainagge. Additionally, individuals residing in Tudun Wazuruchi were regarded as having a low-income status compared to those in Sani Mainagge. Given the varying performance of the clustering methods for different clusters, decisions on study wards in clusters with dissimilar housing and environmental conditions relied on the the opinion of the field team on ease of study implementation. The selected study wards in Ibadan were four - Olopomewa, Challenge, Agugu and Bashorun while in Kano metro area they were five - Dorayi, Fagge D2, Tudun Wazurchi, Gobirawa, and Zango (Table 1). The final selected study locations represent a diversity of environmental and living conditions in both Ibadan and Kano metropolis. Site visit notes and pictures for each ward are available upon reasonable request to the authors.

### Qualitative study methods

#### Defining formal and informal settlements and slums, and categorizing neighborhoods by settlement types using Multi-stakeholders’ Dialogues (MSDs) to inform sampling plans

Multi-stakeholders’ dialogues will be held to define formal and informal settlements and slums and to categorize neighborhoods by settlement types to inform the sampling strategy within selected wards. Multi-stakeholders’ dialogues or roundtables are structured processes used to bring stakeholders together to develop a shared understanding of issues, evidence and plans of action [44]. One MSD will be conducted per city. The MSDs will use a participatory community mapping approach. During a roundtable discussion, participants will reach a consensus on the characteristics of different settlement types. Each participant will then be given a form and asked to provide general impressions of the study wards and to record the number of communities/neighborhoods that can be described as formal settlements, informal settlements, or slums. Subsequently, participants familiar with or residing in each of the study wards will be asked to sketch a community map with detailed descriptions of various settlement types and to present the final product to the full group of participants for feedback. To facilitate the map sketching process, participants will be provided with a validated map of each ward showcasing major landmarks and streets.

A total of 10 participants will be recruited for the MSDs in Ibadan and Kano, respectively. Participants will include a Town Planner or a Quantity Surveyor from the State Ministry of Lands and Housing, a Building/Structural Engineer from State Ministry of Works, a Surveyor from the Office of the Surveyor-General, a Statistician/Demographer or Population Expert from State Bureau of Statistics and an academic with expertise in Urban Planning or Geography. Other categories of participants that will be included in the MSDs are two stakeholders who work as Estate Managers/Agents and two community members residing in a formal and informal settlement as well as one intra-city experienced commercial taxicab or shuttle driver who is familiar with various areas within the metropolis. The recruitment of pertinent stakeholders for the MSDs will be facilitated using purposive and snowballing procedures. Community gatekeepers and heads or senior officials in various relevant ministries and institutions will be consulted and engaged for the purpose of recruiting distinguished and well-informed individuals for the MSDs. In addition to the 10 participants that will be involved in each of the MSDs, a stakeholder from the state level/office of NMEP will be involved in the dialogue as a non-participant observer. The Supplement includes a matrix showing details of the description of the participant types and how each participant will be sampled, the study guide and a sample of the informed consent form for the MSDs. The MSDs will be audio-taped and led by a rapporteur and two facilitators (moderators) who are experts in qualitative research and community engagement.

#### Focus group Discussions and Key Informant Interviews to inform the design of human survey questions

In Ibadan and Kano metro areas, Focus Group Discussions (FGDs) and Key Informant Interviews (KIIs) will be conducted among purposively selected participants spread across various communities using guides comprising open-ended questions and prompts (See Supplement pages 16 - 43) with goal of generating information to guide the design of the quantitative study. The FGDs and KIIs will be designed to explore the following: 1) community management of suspected malaria infections, 2) malaria health-seeking behaviors, 3) how to best support study participants to recall whether and what malaria medications were used for treatment after a suspected or confirmed malaria episode, and 4) methods employed by community members to protect themselves from malaria. As shown in Table 2, the FGDs will target community members comprising caregivers of under-fives, adult male and adult female groups, one each in formal, informal settlements, and slums (Areas designated as formal, informal settlements and slums will be based on the products of the MSDs). Each FGD group will comprise 8 – 12 participants. A minimum of eleven FGDs each will be conducted in Ibadan and Kano, respectively, of which two FGDs will be used to pre-test the study instruments. Data will be collected up till a point of saturation. For each FGD, participants will be recruited purposively and with the help of community gatekeepers or community liaison officers. Community gatekeepers and/or community liaison officers will also be identified through a community engagement process.

**Table 2:**
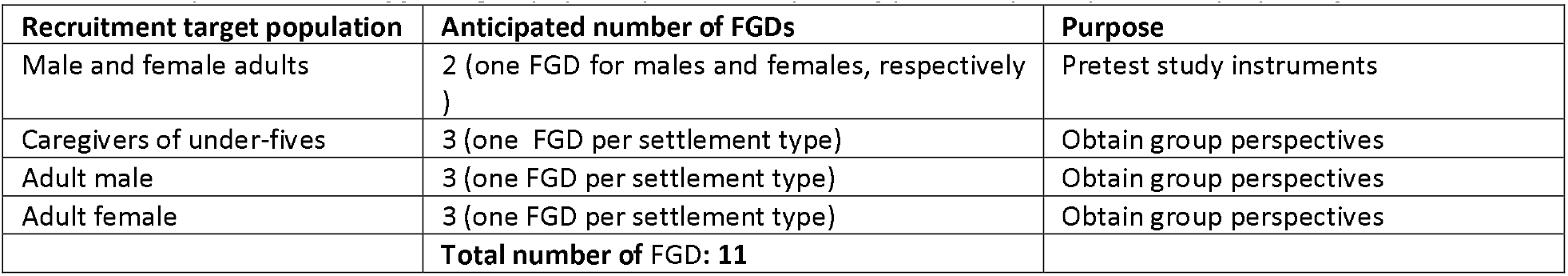
Anticipated number of focus groups per city and description of potential participants and purpose of each discussion

| Recruitment target population | Anticipated number of FGDs | Purpose |
| --- | --- | --- |
| Male and female adults | 2 (one FGD for males and females, respectively) | Pretest study instruments |
| Caregivers of under-fives | 3 (one FGD per settlement type) | Obtain group perspectives |
| Adult male | 3 (one FGD per settlement type) | Obtain group perspectives |
| Adult female | 3 (one FGD per settlement type) | Obtain group perspectives |
|  | **Total number of FGD: 11** |  |

The KIIs will target community, formal health sector and informal health sector stakeholders in the different settlement types in Ibadan and Kano. Table 3 presents information on the recruitment targets and anticipated number of KIIs for each group. Key informants will be recruited purposively from various areas and settlement types within the study sites. A minimum of twenty KIIs will be conducted in addition to two KIIs, which will be used to pretest the study instruments. The final number of FGDs and KIIs will depend on our team’s assessment of data saturation. A separate KII guide will be used for each of the three categories of the key informant interviewees, namely, community stakeholders, health workers in formal healthcare sectors and healthcare providers in informal sectors.

**Table 3:** Description of key informant interview types, recruitment targets and proposed number of interviews per city

| Key informant interview (KII) target stakeholder | Recruitment target population | Anticipated number of KIIs | Purpose |
| --- | --- | --- | --- |
| Health sector stakeholders | Formal health worker and informal health worker | 2 KIIs | To pretest study instruments |
| Community stakeholders | Community/opinion leaders, traditional leaders (one community leader from each of the three types of settlements) | 4 KIIs | To obtain key stakeholders' perspectives |
| Formal health sector stakeholders | Heads of primary health care centres, PHC coordinators, LGA medical officers, LGA Roll- | 10 KIIs | To obtain key stakeholders' |
|  | back Malaria Focal Persons, Public and private sector pharmacists and doctors, and State Malaria Programme Officers |  | perspectives |
| Informal health sector stakeholders | Patent Medicine Vendors, drug peddlers/hawkers, traditional/herbal healers (two separate KIIs from each of the typologies of informal health workers across the three different settlement types) | 6 KIIs | To obtain key stakeholders' perspectives |
|  |  | **Total number of KIIs: 22** |  |

The KIIs and FGDs will be conducted by a team of two trained Research Assistants (RAs) comprising a moderator and a recorder/note-taker and will be supervised by field supervisors, a quality assurance manager, and the social scientist/qualitative research expert on the research team. All KIIs and FGDs will be audio-taped. Study RAs will be university graduates with experience in conducting qualitative research and will be fluent in English language as well as Yoruba and Hausa for Ibadan and Kano study sites respectively. The selection of RAs will be gender sensitive. The FGDs and KIIs will be conducted in locations where the privacy of participants can be guaranteed.

#### Cognitive pretesting of questions from the Nigerian Demographic and Health Surveys to inform the design of human survey questions

The Demographic and Health Survey questionnaire comprise validated malaria illness-related, health-seeking, and financial status-related questions that may be useful for the study. To assess its utility among the study population, recall and comprehension of pertinent questions will be assessed among mothers of under-five children using cognitive pretesting (see Supplement pg 51 for guide). Cognitive pretesting, also known as Cognitive Interview (CI), will allow the study team to generate suggestions for improving assessed questions in the quantitative surveys.

A minimum of four caregivers of under-five children will be purposively selected across Ibadan and Kano metro areas for the CIs. Each CI will be facilitated by a moderator and a note-taker. The CIs will be conducted to a point of data saturation.

### Cross-sectional study methods

#### Household surveys

To assess ward-level malaria prevalence among all-ages, and children under the age of five years as well as related factors (socio-economic, behavioural, intervention coverage, mobility), household-based surveys of individuals of all ages, including pregnant women, from the different settlement typologies will be conducted in the wet season (May – August) and dry seasons (November – February) in the study wards in Ibadan and Kano. The same households will be sampled in the dry and wet seasons. Survey instruments are presented in the Supplement (page 57), and will be revised based on the results of the qualitative study. A positive Rapid Diagnostic Test (RDT) will be used to detect the presence of malaria infection. Individuals with fever (axillary temperature 37.50C) and a positive RDT test will be given a treatment dose of artemether lumefantrine or referred to the health facility. Severe malaria, severe anemia, non-malarial illnesses, or illnesses judged to require more than oral antimalarial treatment will be referred to local health facilities. Blood spot samples will be collected on filter paper and stored for further analysis using polymerase chain reaction (PCR). The sampling frame will be extracted from the 2006 National Population and Housing Census of the Federal Republic of Nigeria (NPHC), conducted by the National Population Commission (NPC). Each ward is subdivided into census enumeration areas (EAs) and the EA will serve as the Primary Sampling unit (PSU). The number of households and population for each EA will be based on data from the 2021 National Population Commission Household survey.

Initial household and individual-level sample size estimation: Survey sample size estimation was based on cluster sampling methodology. Malaria prevalence estimates of children under the age of five years from all urban clusters in Oyo and Kano state in the 2018 NDHS (29% and 27% prevalence, respectively) were used to approximate ward-level prevalence estimates in all ages and children under the age of five years in Ibadan and Kano, respectively. Expected prevalence values were combined with a design effect (*Deft*) of 1.2, a relative standard error (α) of 0.05, and an assumption of a 90% response rate at the household (*R*_*h*_) and individual levels *(R*_*i*_) to estimate the number of households per study ward as 1,741 in Ibadan and 1,923 in Kano. The formula below was used for the household sample size estimation.

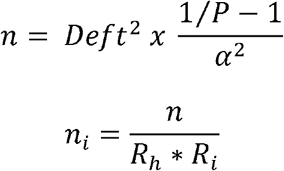

Where *n* is the household interview sample size without accounting for non-response, and *n*_*l*,_ is the household interview sample size after adjustment for non-response

Rapid diagnostic tests are expected to be administered to five or fewer individuals per household depending on the household size. If the overall average number of individuals per household is assumed to be 3.9 persons in Ibadan and 5.7 persons in Kano, the number of individuals expected to be available for RDT, after accounting for a 90% response rate, is estimated to be 6,111 per ward in Ibadan and 7,835 per ward in Kano.

#### Final sample sizes after sample allocation

To allocate household interview sample size estimates of 1,741 and 1,923 per ward in Ibadan and Kano respectively, we will stratify communities into formal, informal and slum settlements using the criteria for defining settlement type obtained from the MSDs. Subsequently, we will select 35 settlements (enumeration areas (EAs)) from each study ward in both Ibadan and Kano. Within each ward, the 35 EAs will be selected proportional to the ratio of formal settlements to informal settlements to slum settlements. Dividing the total household sample size (1,741 and 1,923 respectively in Ibadan and Kano) by the 35 enumeration areas to be selected, we estimate that 50 households (after rounding to the nearest whole number) will be interviewed per enumeration area in Ibadan and 55 in Kano. This implies that a total of 1,750 and 1,925 households will be sampled per ward in Ibadan and Kano respectively. The RDTs expected to be administered per study ward, per season in Ibadan (under the same assumption of 3.9 persons per household in Ibadan and 5.7 persons in Kano), after adjustment for a 90% response rate, will then be 6,142 in Ibadan and 8,663 in Kano. Across all four wards in Ibadan and for each season, 7,000 households are expected to be sampled and 24,568 RDTs are expected to be administered while in Kano, 9,625 households are expected to be sampled and 43,315 RDTs administered in all five wards in Kano and for each season.

#### Household sampling strategy

In the study wards in Ibadan and Kano metropolis, study households will be randomly selected and household heads or representatives will be interviewed. RDTs will be administered to all individuals in the household when the number of household members is five or fewer. When there are more than 5 household members, they will be stratified into five age groups (0 - 5, 6 - 10, 11 - 17, 18 - 30, and >30), and one member from each group will be tested. In households where individuals are missing in a particular age category and household members are more than five, this will be compensated for by testing an additional household member in the youngest age category. Household residents tested for malaria using RDTs will also provide blood spot samples. RDTs and blood spot sample collection with filter paper will be performed by research assistants/laboratory technologists who will undergo a two-day training workshop on survey procedures and a refresher course on sample collection procedures and malaria diagnosis with RDT. Participants will provide a single blood sample for the malaria RDT and collection with filter paper. RDT results will be read as per the manufacturer’s instructions and recorded separately on the patient’s questionnaire. Test results will be passed to the health care provider to use in managing the patient. Blood spot results will be used for later confirmation of RDT results with PCR at the conclusion of field work. Data and location data will be collected via interviewer-administered questionnaires using GIS-enabled Android tablets. This will enable online and real-time access to collected data.

#### Computation of sampling weights

Sampling weights will be applied because of the non-proportional allocation of the sample size to the different wards and the potential differentials in response rates across the wards. The sampling weight per individual will be the inverse of the first-stage sampling probability of an EA from a specific ward and the inverse of the second-stage sampling probability of the household.

#### Health facility surveys

To support the mapping of malaria prevalence among children under the age of five years within unsampled wards in Ibadan and Kano metro area, Health Facility Surveys (HFS) will be conducted among pregnant women using survey instruments presented in the Supplement (pages 74-75). These instruments will be revised based on the results of the qualitative study. We will recruit pregnant women attending antenatal care (ANC) for the first time during their first pregnancy and residing in any ward in Ibadan and Kano metropolis. We are recruiting women during their first pregnancy (primigravidae) as this was found to have the best correlation with malaria prevalence in children under the age of five compared to all gravidae and multigravida [45]. Those presenting with emergency conditions will be excluded. Similar to the cross-sectional survey, a positive RDT test will be used to detect the presence of malaria infection and dried blood spot samples will be collected on filter paper for PCR. Those testing positive for malaria will be treated according to national guidelines. Ward residence data will be collected for each respondent to facilitate mapping of malaria prevalence in unsampled wards.

##### Health facility selection

Using data for first-time ANC attendees in 2021 from the Nigeria District Health Information System (DHIS2), as provided by the NMEP, one non-tertiary health facilities with the largest ANC attendance was purposively selected for the HFS per LGA in Ibadan and Kano metro. The study locations were selected from LGAs in Ibadan and Kano metro rather than wards because the catchment area for health facilities is typically at the LGA-level. The HFS will be conducted monthly for three months in each health facility during the wet and dry seasons respectively.

##### Sample size estimation and allocation

Due to a paucity of literature on malaria prevalence among pregnant women residing in Ibadan and Kano metropolis, the ward-level sample size was estimated based on a proportion of 50% positivity among new ANC attendees using a similar method as the NDHS surveys [46]. A design effect of 1.5 was used to account for the clustering of pregnant women in a health facility and RSE was kept at 5%. The same prevalence was assumed for each month of the year. Therefore, the sample size was initially calculated per month using the formula below to get an estimate of 900 ANC attendees per city.

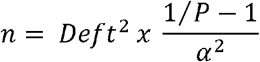

Where: *P*=assumed malaria prevalence among pregnant women; α = RSE and *Deft* = Design effect

Because the target population of new ANC attendees per month in Ibadan and Kano was less than 900, the estimated sample size was further adjusted for the population size using the average number of ANC attendees between July and September from data provided by the NMEP from routine surveillance. The final sample was then calculated using the finite population correction formula below.

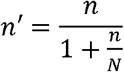

Where: *n* = estimated sample size; *N* = Average of total ANC attendance and *n’* = final sample size estimate

Following adjustment for 85% response rates, the sample size per month for Ibadan and Kano was computed as 406 and 775 new ANC attendees, respectively. The overall anticipated sample size for six months (3 months per season) in Ibadan is 2,436 in Ibadan, and 4,650 in Kano. Detailed sample size allocation for Ibadan and Kano metro are shown in Tables 4 and 5. Sample size allocation for each health facility was proportional to its average ANC attendance from July – September 2021 (Table 4 and 5).

**Table 4:**
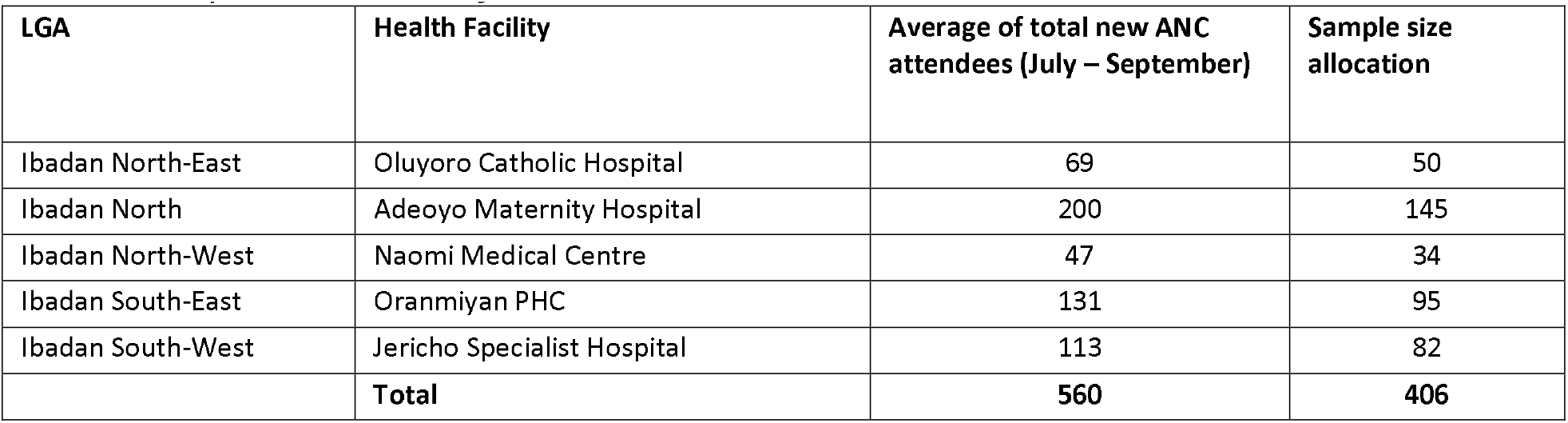
Sample size allocation for HFS in Ibadan metro

| LGA | Health Facility | Average of total new ANC attendees (July – September) | Sample size allocation |
| --- | --- | --- | --- |
| Ibadan North-East | Oluyoro Catholic Hospital | 69 | 50 |
| Ibadan North | Adeoyo Maternity Hospital | 200 | 145 |
| Ibadan North-West | Naomi Medical Centre | 47 | 34 |
| Ibadan South-East | Oranmiyan PHC | 131 | 95 |
| Ibadan South-West | Jericho Specialist Hospital | 113 | 82 |
|  | **Total** | **560** | **406** |

**Table 5:**
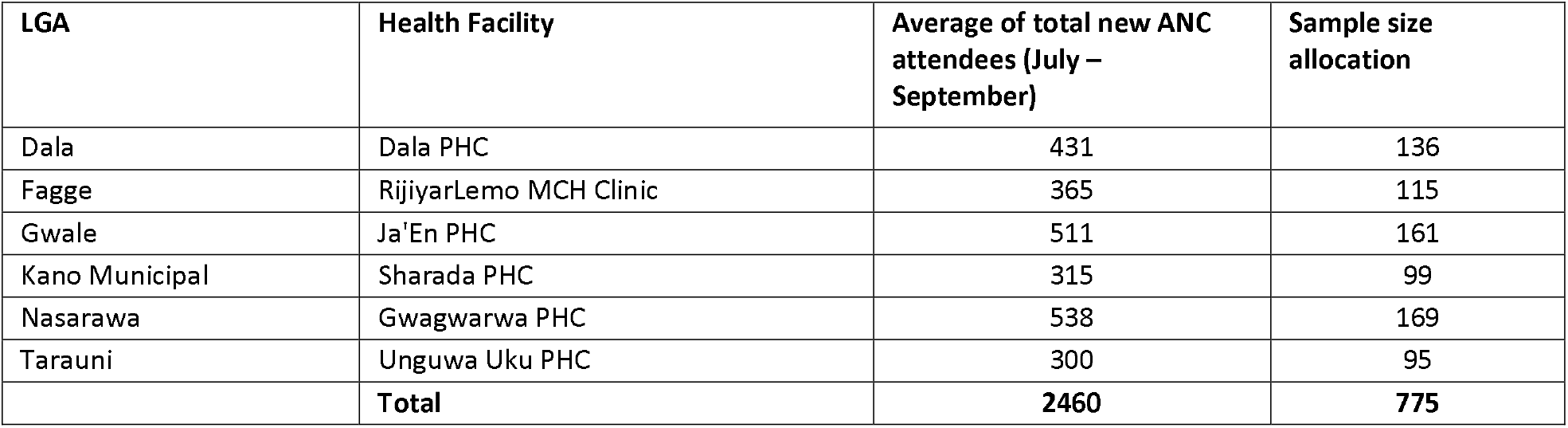
Sample size allocation for HFS in Kano metro

| LGA | Health Facility | Average of total new ANC attendees (July – September) | Sample size allocation |
| --- | --- | --- | --- |
| Dala | Dala PHC | 431 | 136 |
| Fagge | RijiyarLemo MCH Clinic | 365 | 115 |
| Gwale | Ja'En PHC | 511 | 161 |
| Kano Municipal | Sharada PHC | 315 | 99 |
| Nasarawa | Gwagwarwa PHC | 538 | 169 |
| Tarauni | Unguwa Uku PHC | 300 | 95 |
|  | **Total** | **2460** | **775** |

##### Sample collection

All new ANC attendees at the facility will be interviewed and screened for malaria using RDT until the required number per health facility and month is achieved. The data to be collected will be similar to the information collected in the household cross-sectional surveys. New ANC attendees will be screened for malaria with RDT and interviewed in a specially designated area/room (that can guarantee confidentiality of information) in the health facility by research assistants/laboratory technologists who will undergo a two-day training workshop on survey procedures and a refresher course on blood sample collection and malaria diagnosis using RDT. Each pregnant woman will provide a single finger-prick blood sample for the malaria RDT and additional confirmation with PCR using blood spot samples at the conclusion of the study. The RDT results will be read as per the manufacturer’s instructions and recorded separately on the patient’s questionnaire. Test results will be passed to the health care provider to use in managing the patient. Data will be collected via interviewer-administered questionnaires using GIS-enabled Android tablets.

### Longitudinal surveys

To estimate malaria seasonality and related factors, a sample of children, 0 – 10 years, will be followed for 12 months and data will be collected using survey instruments in the Supplement (page 95). Sample size: We used an open-source sample size calculator provided by OpenEpi [47] for cohort studies (https://www.openepi.com/SampleSize/SSCohort.htm) to estimate the sample size per ward. An initial sample size of 228 per ward was estimated, based on Fleiss et al. [48]. All parameters used as input for the sample size calculation are provided in Table 6. Adjusting for 20% attrition, the minimum sample size was computed as 285 children per ward. Two wards that have a good mix of formal and informal settlements from the selection made for the cross-sectional survey will be purposively selected in Ibadan and Kano. The total sample size for the two study wards each in Ibadan and Kano will be 570 children.

**Table 6:** Parameters used for computing longitudinal study sample size

| Parameter | Value (source) |
| --- | --- |
| Two-sided significance level | 95% |
| Power | 80 |
| Ratio of unexposed to exposed in sample | 1.0 |
| Percent of unexposed with outcome (percent of ITN users with malaria) | 30.33% (based on malaria incidence estimates from the WHO Observatory [49]) |
| Risk Ratio | 0.49 [50] |

#### Participant selection and field methods

Participants will be selected/recruited at the community level. A household listing will be carried out and used as a sampling frame for a random selection of eligible households with mother-child pairs.To be eligible for enrollment, the mother-child pair should be residing in the household for the next year. Two individuals residing within each study ward will be recruited and trained as Community Monitors (CM) to identify eligible study participants, conduct interviews, and administer RDT tests on the study participants. Additionally, a supervisor will be trained to oversee the activities of the CMs in each ward, and monitor data collection and follow-up. The study will be explained to mothers/caregivers of children and those who give informed consent will be enrolled. Participating mothers/caregivers and the selected children will be visited at home once in a month during the one-year follow-up and will be encouraged to visit the CMs any time a child enrolled in the study has a case of suspected uncomplicated malaria.

At the initial visit, the CMs will collect information regarding the participant’s family, including basic demographic information and information and the use of malaria prevention practices. Participants will also be informed to report to the CMs in their ward whenever their under-five child develops a febrile illness. At each monthly visit, the CMs will administer a standardized questionnaire, collecting information regarding illnesses that had occurred since the last visit, symptoms experienced, use of health-care facilities, and use of medicines and vector control tools. In addition, information on travel history and the cost of treatment will be collected. Anthropometric measurements will also be done. Axillary temperatures and finger or heel prick (neonates) blood samples will be collected for RDT to determine parasitemia and blotted samples for PCR on filter paper will be obtained for confirmation of cases detected using RDT. Any recruited child with documented fever (axillary temperature 37.5° C) and malaria parasitemia judged to be uncomplicated malaria will be given a treatment dose of artemether lumefantrine or referred to the health facility. Severe malaria, severe anemia, non-malarial illnesses, or illnesses judged to require more than oral antimalarial treatment will be referred to local health facilities. To reduce loss to follow-up, we shall have the telephone numbers of the mother/caregiver as well as that of a very close person/another household member whom we can call to check his/her whereabouts. During non-scheduled visits with the CM, that is when the child has a case of suspected uncomplicated malaria, similar protocols as in the monthly visits will be followed.

### Entomological surveys

To understand the larval habitats, indoor and outdoor transmission rates, species composition and dynamics, biting rates and inoculation rates of *Anopheles* vectors, entomological surveys will be conducted for four months in the dry and wet season, respectively.

#### Selection of the study sites

Due to funding limitations and because we do not expect vector dynamics to vary widely across wards with similar environmental conditions, three wards each will be selected in Kano and Ibadan from the study ward selections in Table 2 informed by the outcome of the MSDs. In each ward, a site for indoor and outdoor mosquito collection will be established in accordance with NMEP protocol for entomological surveys. Households selected for the entomological study will be from those enrolled in the longitudinal study. Study households will be mobilized and asked to complete informed consent forms (See Supplement page 102).

#### Indoor and outdoor mosquito collection with Centers for Disease Control (CDC) light trap

CDC light traps will be set-up indoor and outdoor in the selected house in each site. The trap will be suspended at 1.5m towards the leg of an adult sleeping inside an untreated bednet. The traps will be operated between 6pm to 6 am for three consecutive days per month in accordance with NMEP protocol for routine indoor and outdoor collection of mosquitoes using CDC light traps. The cups of the traps will be collected hourly and mosquitoes by the traps will be killed by chloroform and recovered into well labeled cups for identification.

#### Collection of indoor resting mosquitoes using Pyrethrum Spray catches

Pyrethrum Spray Catch (PSC) will be conducted monthly in ten randomly selected houses in each site. This is to collect indoor resting mosquitoes. The method involves spreading a white cloth to cover all the space in the selected household after which an insecticide is sprayed by the Entomologist. The mosquitoes that fall on the cloth will be recovered into the petri-dishes using forceps.The petri-dishes will be labelled and transported into the laboratory for identification.

#### Collection of Environmental data

On each trapping day, a digital device (Thermo Pro, remote sensor 433MHz wireless-Model TP-60) will be used to collect indoor and outdoor temperature as well as relative humidity. The data will be recorded in hardcopy forms.

#### Management and identification of collected mosquitoes

Mosquitoes collected in the field (Indoor, outdoor and PSC) will be transported into the laboratory and sorted based on an hourly catch. The female *Anopheles* mosquitoes will be identified using keys described by Cooetze and Gillies [51] and Gillet [52]. The number of each species of female *Anopheles* mosquitoes identified will be recorded. Each female *Anopheles* mosquitoes will be identified will be coded and preserved in silica-filled 18uardian18 tube (1.5ml) and transported to the NMEP-accredited Molecular Biology Laboratory at Osun State University for molecular identification of species, blood meal source and sporozoite rate using detailed protocols provided in the Supplement (pages 106 – 111).

#### Prospection of the breeding sites and duration

Larval sampling will be carried out in all selected wards in Kano and Ibadan metropolis in both the wet and dry seasons. The microhabitats of potential breeding sites such as puddles, vehicle tires, septic tanks, gutters, tire tracks, wells and run-offs will be prospected. Prospective breeding sites will be selected and sampled using standard dippers [53]. Larvae encountered will be transferred to the collection bowl and taken to a laboratory.The following list describes the procedure for mosquito larval collection at breeding sites:

1. Larval sampling will be done between 0700 to 0900 hours. Water bodies such as swamps, ditches, streams, water puddles, pools, rice fields, drainage, tire tracks, domestic container, tree holes, domestic well, footprint etc. will be located.
2. The dipper will be lowered gently at an angle of about 45° to enable water and nearby larvae to flow into the dipper.
3. The content of the dipper will be emptied into a container and transported to the laboratory. Five dips will be taken per breeding sites in accordance with standard protocol as published in previous studies [45-47].
4. For each sampling operation, the following will be recorded: a) Geographic location (GPS coordinates), b) Name of locality, c) Type of breeding sites (permanent, semi-permanent or temporary), d) Source of the water (rain, river, lagoon or man-made), e) Nature of water collection (puddle, rice-fields, etc.), and f) Number of larvae per dip.

#### Larval identification and species characterization

The larvae will be reared to emerge into adults and then identified morphologically using standard protocols for female *Anopheles* mosquitoes. Molecular identification will be carried out as described in the Supplement (Pages 106 – 111).

### Public Involvement

This study is conducted at the request of the NMEP and in partnership with them to generate evidence to inform intervention tailoring in urban areas in the 2026 – 2030 National Malaria Strategic Plan. Prior to the start of the study, the study team met with the NMEP to discuss and refine the study design and review evidence to inform selection of the study sites. Plans are in place to enable representatives of the State and National Malaria Elimination Programme to serve as observers during data collection.

Mobilization meetings will be held with the key stakeholders before the commencement of the cross-sectional, longitudinal and entomological studies, including: 1) The National Coordinator, Head, Integrated Vector Management, and Head of Case Management of NMEP 2) Malaria Programme Managers in Kano and Oyo States, 3) LGA Malaria Coordinators in the Selected LGAs, and 4) community leaders of the selected communities and households. The study protocols will be explained to all stakeholders including the households to be used for the study.

### Data analysis plan

#### Qualitative data

Data processing will start with the verbatim transcribing of tape recordings of the MSDs, FGDs, KIIs and Cis immediately after the data collection to avoid loss or omission of important details. Results of the MSDs, FGDs as well as the interviews (KIIs and Cis) conducted in Yoruba and Hausa, will be translated into English to ascertain the quality of the data. All transcribed notes will be audited and validated by project team members. Validated transcribed notes will be entered into the computer using NVIVO version 12 Pro. Inductive-dominant coding approach will be used to drive the coding process [35]. Parent nodes (primary codes) and child nodes (secondary codes) will be generated based on the content of the data. The codes will be linked appropriately to the corresponding quotations. Memos will be created as necessary and linked with appropriate codes and quotations. The generated codes and the quotations will be reviewed and critiqued carefully by the data analyst and experienced qualitative research experts on the project team.

Thematic content analysis will be performed, and generated themes will be based on the (a) content of the study instrument (b) sample quotes from transcripts; and (c) peer review and reflections (contributions from members of the project team). Based on the step-by-step approach of thematic analysis identified by Nowell et al [36], the verbatim transcript of each of the MSDs, FGDs and interviews (KIIs and Cis) will be carefully read, examined and juxtaposed theme by theme to identify relevant texts, repeating words, similar phrases and divergent opinions. For each theme, common and peculiar trends, as well as similar and divergent opinions, will be noted. Themes will be developed and revised iteratively. Additionally, the manifest and latent contents of the MSDs, FGDs, KIIs and Cis data will be explored. A summary of findings will be written, and samples of appropriate verbatim quotes will be provided. Findings of the qualitative data will be used to guide the development of questionnaires for the cross-sectional and longitudinal studies.

#### Hoousehold, health facility and longitudinal surveys

Descriptive analysis of de-identified data from the household and health facility surveys will be conducted to generate initial estimates of malaria prevalence by dry and wet season for sampled and unsampled wards in Ibadan and Kano metro. Spatial interpolation methods will be applied to generate smoothed estimates for all wards borrowing information from areas with more data, that is, wards sampled in the household surveys, to generate estimates in areas with fewer data points, that is unsampled wards with malaria test data from the health facility surveys. Additional analysis will be conducted to estimate bed net use, bed net coverage and mobility metrics at the ward-level. Regression models will additionally be used to examine the association between observed malaria prevalence and individual and community-level factors.

Descriptive analysis of de-identified data from the longitudinal surveys will be conducted to generate monthly malaria test positivity rates by RDT among children 0 – 10 years for Ibadan and Kano metro area. Regression-based methods for repeated measures will be used to examine the association between individual and community factors and monthly malaria test results

Entomological data will be analyzed to evaluate the following: 1) habitat occupancy, 2) types of *Anopheles* found and the nature of habitats utilized by each species in each site in both seasons, 3) larval density, 4) man biting rate, 5) human blood index, 6) indoor resting density, and 7) Sporozoite rate. The Supplement (page 118) provides a detailed description for how these entomological variables will be assessed. In each study location, *Anopheles* habitat will be geo-referenced and a habitat distribution map for each area will be created.

Ward-level mathematical models will be developed using the above entomological parameters and calibrated to all-age prevalence and incidence data from the field study, to estimate the impact of intervention scenarios specified by the NMEP on future transmission dynamics.

## Data Availability

All data produced in the present study are available upon reasonable request to the authors

https://dhsprogram.com/

## Ethics and dissemination

This protocol has been approved by Nigeria’s National Health Research Committee and Ethical Review Committees of Oyo and Kano States Ministry of Health, and Northwestern University. Study participants will be asked to provide either written or verbal informed consent prior to being recruited into the study. Separate informed consent forms are provided for each of the components of the study including qualitative studies, household surveys, health facility surveys, longitudinal surveys, and entomological surveys (see Supplement). The informed consent forms will be made available in English as well as major local languages of the study sites (Yoruba and Hausa). In addition, during community mobilization, information about the study will be presented to the communities and permission will be obtained from the communities’ gatekeepers. They will also be provided information on the process of informed consent and examples of the informed consent forms will be kept with the communities for review. For children 0-10 years enrolled in the longitudinal study, written informed consent will be sought from their parent/20uardian. If the parent/guardian accepts participation on behalf of the child, he/she will be asked to sign two copies of the written informed consent. One copy of the consent form will be given to the parent/guardian, and another kept by the investigator (household survey) / health worker in the facility (health facility survey), a photocopy of which will be kept in the investigator’s office. Assent will be obtained from children selected to participate in the study in addition to the parental consent.

Confidentiality will be ensured to protect the identification and other personal information provided by the participants. Codes will be used to label data from various sources. We will explain clearly to participants that information they supply will be properly stored and kept in encrypted files. The data will be restricted to only authorized persons and stored only for a maximum of 5 – 7 years after which data will be destroyed after the data would have been published.

Participants will be given incentives to compensate for the time they spent in participating in the study as well as a transport fee if applicable. We will express clearly to the participants how we appreciate their time and efforts. Where necessary and where participants request, we will offer free of charge supports services such as referral advice and counselling to participants after their participation in the study.

An exit briefing/dissemination meeting will be held with all the relevant stakeholders and communities where the studies are conducted within four weeks upon completion of field work and analysis of the data. The dissemination meeting will also be used to educate the selected communities on the transmission of malaria, health and socio-economic impacts and various methods to protect individuals and communities against malaria. We will also inform participants about how the findings of the study will be disseminated (e.g., reports, conference presentations, journal articles).

## Authors’ contributions

IDO conceptualized the study and its design, identified funding for the study, and led writing of the study protocol. IA, MA, AOO, OS, JA, AF, MB, and AYA led study design, including sample size estimation, and co-wrote the protocol. EB developed the quantitative questionnaire and supported study design. PU, CA, and CO provided feedback on the study design and review feedback for the protocol.

