## Supplementary Material for "Field assessment of the burden and determinants of malaria transmission to inform tailoring of interventions (microstratification) in Ibadan and Kano metropolis: Study protocol"

*Denotes first authors

To whom correspondence should be addressed:

### Study ward selection process

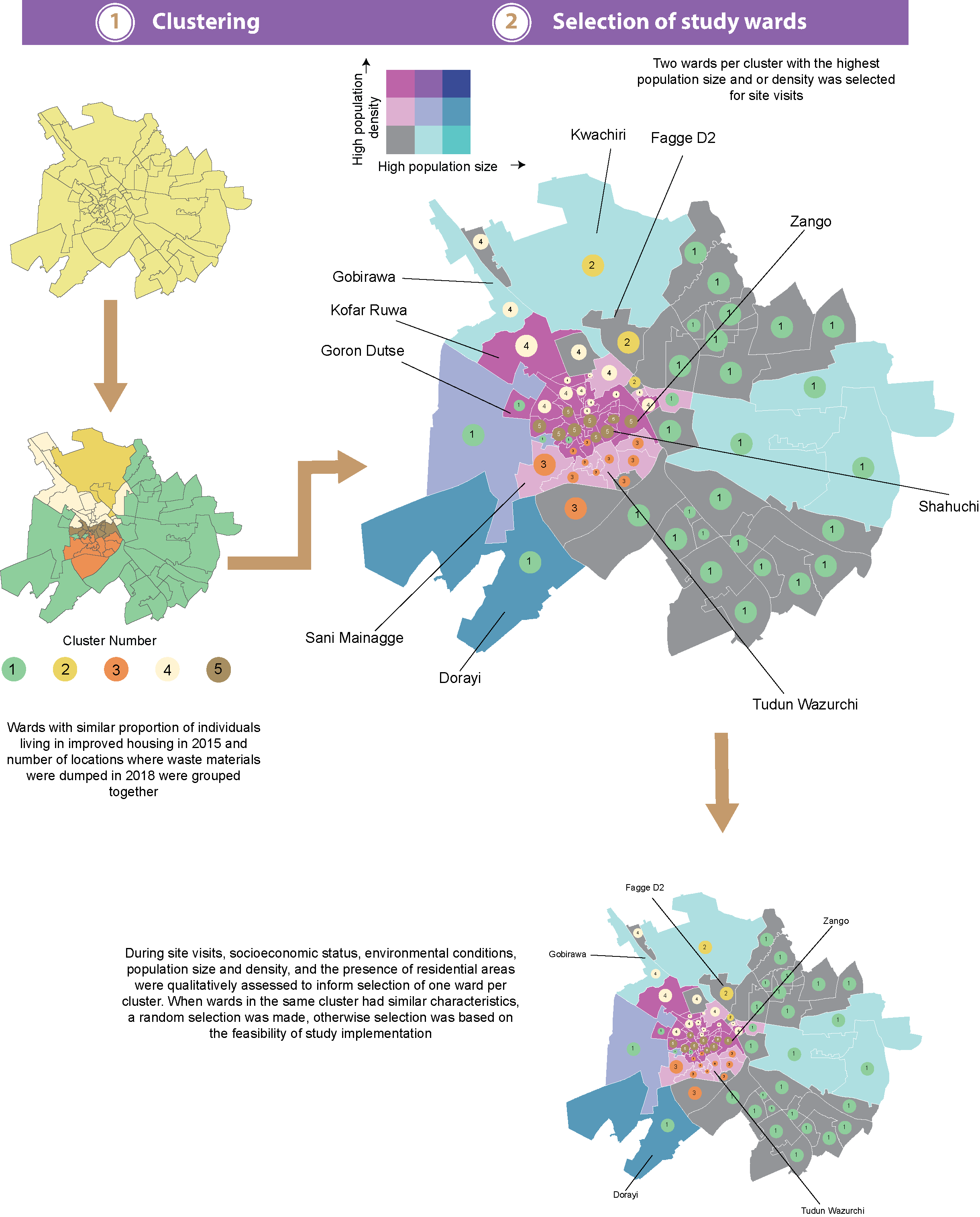

*Figure 1: Overview of the site selection process for Kano*

*Table 1: Field observations from site visits*

|  | **Ibadan metro area** | | **Kano metro area** | |
| --- | --- | --- | --- | --- |
| **Cluster #** | **Ward** | **Site impressions of residents, environmental and living conditions** |  | **Site impressions of environmental and living conditions** |
| 1 | Olopomewa | Roads are tarred and housing infrastructure consists of well-built, fenced and painted cement homes. An abundance of supermarkets, pharmacies and recreational areas were noted. Residents were observed to be high and middle socioeconomic class | Dorayi | Densely populated ward with modern well-built housing, tarred streets surrounded by congested and poorly accessible settlements with lower quality housing. According to locals, the ward used to have a major dumpsite and was recently redeveloped to contain modern housing. Residents of this ward were observed to be mostly of low socioeconomic status |
| 1 | Owode | Most roads are untarred, and this ward abuts multiple slums. Inhabitants are mainly in the low and middle socio-economic class and most houses are used for residential purposes. Site observers observed the ward to have medium population density | Goron Dutse | Wards is part of ancient Kano city, extends beyond the city walls and was perceived to densely populated. Housing quality is varied. Some areas had newer high-quality housing situated in neighborhoods with tarred streets. However, majority of the settlements comprised of congested low-quality housing, which had street access |
| 2 | Challenge | Roads are untarred and gutters are overgrown with plants. Housing infrastructure is a mix of ageing old-style housing architecture with modern and newly built cement housing. Inhabitants were perceived to be middle-income | Fagge D2 | Densely populated ward with medium to poor quality housing quality. Streets are tarred and have good vehicular access that links most neighborhoods. The socioeconomic status of individuals residing in this ward were perceived to vary from low to medium. |
| 2 | Basorun | Roads are roughly tarred and housing is a mix of aged housing together with new housing styles. Ward is home to a popular market where staple foods are sold. Most residential houses abuts shops. Ward was observed to have medium population density and inhabitants were perceived to be of middle income. | Kwachiri | Sparsely populated commercial hub with very few residential houses. It is home to a major mechanic village/garage in Kano and serves as a transit hub for Hajj pilgrims |
| 2 | Asanke | Roads leading to individual homes are untarred but ward has access to major tarred roads that run through the city. Ward population size and density was perceived be high. Houses were aging old-style architecture. |  |  |
| 3 | Agugu | Densely populated ward with residents considered to be low-income. Has poor road network and existing roads are untarred and contain puddles of water. Housing infrastructures are dilapidated cement homes | Tudun Wazurchi | Many roads within this ward have poor vehicular transport accessibility. Major means of transport is on foot. Housing quality is of medium to poor quality, and it is densely populated. Most households were perceived to be in the lowest socioeconomic position. |
| 3 | Ode Aje | Roads are untarred with the exception of one road that provides access to major road networks and another road to provides access to minor road network. Housing is aged and dilapidated. Inhabitants are perceived to be of low income and population density was perceived to be high. | Sani Mainagge | Residents were perceived to be of higher socio-economic status. Housing quality is high as depicted by the many new modern builds. Most roads are well tarred. Ward appears to be sparsely populated |
| 4 | Bashorun | Ward with good road network. Inhabitants were perceived to consist of mostly individuals of high socio-economic status. Neighborhood has several pharmacies, supermarkets and recreational centers like hotels. | Gobirawa | Densely populated ward with medium to poor quality housing. Streets are untarred but are wide enough to enable vehicular transport access across neighborhoods. The socioeconomic status of residents was perceived to vary from low to medium |
| 4 |  |  | Kofar Ruwa | Wards has few residential areas. It is a commercial village comprising of mechanic villagese (garages), commercial parks and building materials shops |
| 5 |  |  | Zango | Housing types range from medium to poor quality. Major roads leading to the ward are tarred. The socioeconomic status of households were perceived to vary from low to moderate. |
|  |  |  | Shahuchi | Housing quality in this ward varied from modern builds to dilapidated structures. Ward was perceived to be densely populated. Ward is the location of a major tertiary health care center. |

**Multistakeholder Dialogue Instruments**

#### Matrix showing suggested participants to be involved in the multi-stakeholders’ dialogues in each site

| **S/N** | **Category of Participant** | **Number** | **Institution/ Affiliation** | **Remarks** |
| --- | --- | --- | --- | --- |
| 1 | Town Planner/ Quantity Surveyor | 1 | Ministry of Lands and Housing | An experienced stakeholder (participant) to be nominated by the Commissioner/Permanent Secretary in the Ministry |
| 2 | Building Engineer/Structural Engineer/Architect | 1 | Ministry of Works and Transport | An experienced stakeholder (participant) to be nominated by the Commissioner/Permanent Secretary in the Ministry |
| 3 | Land Surveyor/Geographic Information System Expert | 1 | Surveyor General Office | An experienced stakeholder (participant) to be nominated by the Surveyor General/Permanent Secretary in the Surveyor General Office |
| 4 | Statistician/Demographer/  Population expert | 1 | State Bureau of Statistics/National Population Commission office at the state level | An experienced stakeholder (participant) to be nominated by the Statistician General/Director in the Agency |
| 5 | Geographer/Urban Regional Planning expert (Academic) | 1 | University of Ibadan/Bayero University | To be nominated by head of department or recruited through snowballing procedure |
| 6 | Estate Manager (Professional)  (Residing or working within formal settlement) | 1 | Private Practice/Community | To be nominated by community gatekeeper(s) or recruited through snowballing procedure |
| 7 | Estate Agent  (Residing or working within informal settlement) | 1 | Private Practice/ Community | To be nominated by community gatekeeper(s) or recruited through snowballing procedure |
| 8 | Intra-City Transport Worker (Taxicab driver)/Thrift collector residing in informal settlement/slum | 1 | Community | To be nominated by community gatekeeper(s) or recruited through snowballing procedure |
| 9 | Community Member (Residing in formal settlement) | 1 | Community | To be nominated by community gatekeeper(s)/Landlord Association or recruited through previous contact. The individual should preferably be an indigene or must have stayed within the metropolis (any of the selected communities/wards) for a least 10 years. |
| 10 | Community Member (Residing in an informal settlement/slum) | 1 | Community | To be nominated by community gatekeeper(s)/Landlord Association or recruited through previous contact. The individual should preferably be an indigene or must have stayed within the metropolis (any of the selected communities/wards) for a least 10 years. |

**NOTE**: 1) As much as possible, participants especially community stakeholders should be drawn across the various selected wards/communities (Olopomewa, Challenge, Agugu, Bashorun). At least there should be one participant from each of the selected areas/communities. 2) A Stakeholder from State Malaria Elimination Programme (SMEP), State Ministry of Health will serve as a non-participant observer.

#### Guide for multi-stakeholder dialogue

##### A. Classification of settlements

1. What can you say about the housing conditions within this metropolis? (***Probe*** *- basic social services and infrastructural facilities- access roads, electricity supply, piped water/drinking water source; waste disposal/management; housing density; housing types; ownership of most housing units*)
2. What are the various types of settlements within the metropolis?
3. How would you categorise the different types of settlement within the metropolis?
4. What are the parameters that can be used to categorize different types of settlements within the metropolis?
5. (a) What is your understanding of formal settlement?

(b) What is your understanding of informal settlement?

(c) What is your understanding of slums?

6. What are your views about categorizing the different types of settlements within the metropolis into formal settlements, informal settlements, and slums?
7. How will you describe a typical formal settlement within the metropolis? (***Probe*** *- What are the unique features? What makes it different from other types of settlements, what type(s) of person(s) live within formal settlements. Please give us typical examples of places with within the metropolis that can appropriately described as formal settlements*)
8. How will you describe a typical informal settlement within the metropolis? (***Probe*** *- What are the unique features? What makes it different from other types of settlements, what type(s) of person(s) live within informal settlements. Please give us typical examples of places with within the metropolis that can appropriately described as informal settlements*)
9. How will you describe a typical slum within the metropolis? (***Probe*** *- What are the unique features? What makes it different from other types of settlements, what category of persons(s) live within slums. Please give us typical examples of places with within the metropolis that can appropriately described as formal settlements*).

##### B. Validating the clustering model outputs

What are the views of various stakeholders about the various selected study locations/wards in the metropolis (in terms of description and typologies of settlements)? **NOTE**: *Ask each of the stakeholders/participants how best the study ward can be described and the various settlement types within the community.*

*Sample for to be used in each study ward (Olopomewa, Challenge, Agugu, Bashorun in Ibadan; Dorayi, Fagge D2, Tundun Wazurchi, Gobirawa and Zango in Kano)*

| S/N | Category of Participant | General description | Formal settlements (areas within the community/ward) | Informal settlements (areas within the community/ward) | Slums (areas within the community/ward) |
| --- | --- | --- | --- | --- | --- |
| 1 | Town Planner/Quantity Surveyor |  |  |  |  |
| 2 | Building Engineer/ Structural Engineer/Architect |  |  |  |  |
| 3 | Land Surveyor/Geographic Information System Expert |  |  |  |  |
| 4 | Statistician/Demographer/  Population expert |  |  |  |  |
| 5 | Geographer/Urban Regional Planning expert (Academic) |  |  |  |  |
| 6 | Estate Manager (Professional) |  |  |  |  |
| 7 | Estate Agent |  |  |  |  |
| 8 | Intra-City Transport Worker (Taxi cab driver)/Thrift collector |  |  |  |  |
| 9 | Community Member (Residing in formal settlement) |  |  |  |  |
| 10 | Community Member (Residing in an informal settlement/slum) |  |  |  |  |
| 11 | Overall consensus among participants |  |  |  |  |

##### C. Instructions for Participatory Community Mapping

1. Each multi-stakeholder dialogue participant who is very familiar with or resides in each of the selected communities will be required to help sketch the community map with detailed identification of various types of settlements
2. Each Multi-stakeholders’ dialogue participant will be given the opportunity to present and explain his/her community map.
3. For each community map being sketched by each participant, contributions will be welcomed from all the multi-stakeholders’ dialogue participants.
4. Possibly if available, properly validated map of each selected community that may be gotten from relevant organization (National Population Commission, World Health Organization or State Bureau of Statistics) will be given to participants to assist them or provide them with some clues so as to make the process of drawing their own maps easier and well validated.
5. Professionals in the multi-stakeholders’ group like town planner, surveyor and geographer will provide useful guidance and suggestions concerning the mapping process.

#### Sample informed consent form for the multistakeholder dialogue

*Introduction*

My name is ……………and other team members are…………… we are convening this multi-stakeholder dialogue as part of the formative aspect and qualitative components of our study on “field assessment of the burden and determinants of malaria to inform tailoring of interventions (microstratification) in Ibadan and Kano Metropolis” which is being implemented through the collaborative efforts of the University of Ibadan, Nigeria as part of a collaborative project with Northwestern University, USA and Nigeria National Malaria Elimination Programme.

We would like to request for your opinion on various issues relating to settlement types and communities within the urban areas of this metropolis to enable us understand malaria transmission in urban areas. You have been specially invited for this multi stakeholders’ dialogue and we thank you for honouring our invitation.

*Purpose*

The main goal of the multi-stakeholders’ dialogue, which involves different categories of stakeholders/experts, is to facilitate brainstorming and identifying criteria for categorising various settlement types and selecting appropriate communities that can appropriately represent formal and informal settlements and slums within the selected wards being considered for the study in Ibadan metropolis. Furthermore, the multi-stakeholders’ dialogue will complement and/or validate the information about the selected wards and settlements obtained through quantitative analysis and site visits.

The information we will be collecting through the multi-stakeholders’ dialogue will help to develop appropriate operational definitions and categorizations for settlement types suitable for the social milieu of the study sites which can be relied upon for guiding for different components of the study we are planning to conduct. Our study findings will potentially contribute to malaria control efforts in Nigeria.

*Procedure*

As part of this study, you will be placed in a group of 10 individuals who represent various stakeholders and experts for the purpose of brainstorming and having cross-fertilization of ideas on pertinent issues. The multi-stakeholders’ dialogue which will last for 90 - 120 minutes will entail asking you some open-ended questions which will be discussed in form of a group discussion. We will be asking you to share your opinions on settlements typologies and the various communities being considered for the study. During this dialogue, your views will be respected and will not be used against you in any way. The multi-stakeholders’ dialogue will be taped, so please speak up and speak clearly. We ask for your consent to record the multi-stakeholders’ dialogue so that we will not miss out anything from the information you will be providing to us through this dialogue. Please do share your views without mentioning people’s names. We want the multi-stakeholders’ dialogue to be anonymous and to be confidential as possible. You can choose whether to participate in the multi-stakeholders’ dialogue, and you may stop at any time during the dialogue. There is no right or wrong view, so feel free to express yourself. Out of respect, please refrain from interrupting others. However, feel free to be honest even when your responses counter those of other dialogue participants. Remember your participation in this dialogue is voluntary. Your decision not to be involved or drop out at any point will not attract any penalty.

*Benefits*

Your participation in this study may not provide any personal benefit to you. However, should you decide to participate in this study, you will be doing society a great service because the findings of this study will be useful in the design of interventions and programmes for the control and prevention of malaria.

*Risks*

There are no known or anticipated risks associated with participation in this study beyond those experienced during an average conversation. If a question, or the discussion, makes you uncomfortable, you can choose not to answer.

*Confidentiality*

Should you choose to participate in the study you will be asked to respect the privacy of other participants of the multi-stakeholders’ dialogue by not disclosing any content discussed during the study. The information you share will be kept confidential. Identifying information will be removed from the transcript and report of the dialogue. The tape-recording of the dialogue will be retained for a maximum of 5 years, after which they will be destroyed. Data will be stored in an encrypted folder on protected laptop. Only the research team will have access to study data. No identifying information will be used in any presentations or publications based on this research. Although we will ask all participants in the multi-stakeholders’ dialogue to maintain confidentiality, we cannot guarantee that they will do so.

*Contact*

If you have any questions or concerns regarding this study, please contact:

Professor IkeOluwapo Ajayi

IMARAT, College of Medicine, University of Ibadan

Thank you for choosing to participate in the study. Kindly show by using any of the following two boxes, that your participation in this study was voluntary.

I will participate	I will not participate

### Focus Group Discussions and Key Informant interview Instruments

#### Focus group discussion informed consent form

*Who to interview*: Groups (8 – 12 persons) of community members that are homogenous- These include:

1. Male adults
2. Female adults
3. Mothers of under-five children

*Introduction*

My name is ……………and my colleagues are…………… We work with the University of Ibadan. We would like your opinion on various issues to enable us understand malaria transmission in urban areas as part of a collaborative project with Northwestern University, USA and Nigeria National Malaria Elimination Programme. The information we collect will help the government to plan health services to prevent malaria infections by ensuring you receive suitable interventions. You have been specially invited for this focus group discussion and we thank you for honouring our invitation.

*Purpose*

The purpose of this focus group discussion is to investigate the following

1. How community members manage suspected malaria infections
2. Where community members seek care for malaria
3. Factors that influence care seeking for malaria in a hospital,
4. How health seeking behaviour and source of care differ by socioeconomic group
5. Local names for malaria medications
6. Whether showing of pictures or physical packaging of malaria medications can improve participants’ recall and answers about usage of malaria medication
7. Factors that influence decisions about what malaria medicine to use
8. Common methods that people use to protect themselves from themselves from malaria
9. Understand facilitators and barriers to participating in a community-based disease reporting programme for malaria

The information learned in this focus group will be used to guide the development of other components of the study that we are planning to conduct.

*Procedure*

As part of this study, you will be placed in a group of 8 – 12 individuals. The focus group discussion which will last for 45 – 90 minutes will entail asking you some open-ended questions. In addition to the discussion questions, we will be asking for some of your socio-demographic information. During this discussion, your views will be respected and will not be used against you in any way. This discussion will be taped, so please speak up and speak clearly. We ask for your consent to record the discussion so that we will not miss out anything from the information you will be providing to us through the discussion. Please do share your views without mentioning people’s names. We want the discussion to be anonymous and to be confidential as possible. You can choose whether to participate in the focus group discussion, and you may stop at any time during the study. There is no right or wrong view, so feel free to express yourself. Out of respect, please refrain from interrupting others. However, feel free to be honest even when your responses counter those of other group members. Remember your participation in this discussion is voluntary. Your decision not to be involved or drop out at any point will not attract any penalty.

*Benefits*

Your participation in this study may not provide any personal benefit to you. However, should you decide to participate in this study, you will be doing society a great service because the findings of this study will be useful in the design of interventions and programmes for the control and prevention of malaria.

*Risks*

There are no known or anticipated risks associated with participation in this study beyond those experienced during an average conversation. If a question, or the discussion, makes you uncomfortable, you can choose not to answer.

*Confidentiality*

Should you choose to participate in the study you will be asked to respect the privacy of other focus group discussion members by not disclosing any content discussed during the study. The information you share will be kept confidential. Identifying information will be removed from the transcripts. The transcripts and other electronic data will be retained for a maximum of 5 years, after which they will be destroyed. Data will be stored in an encrypted folder on protected laptop. Only the research team will have access to study data. No identifying information will be used in any presentations or publications based on this research. Although we will ask all participants in your focus group to maintain confidentiality, we cannot guarantee that they will do so.

Contact

If you have any questions or concerns regarding this study, please contact:

Professor IkeOluwapo Ajayi

IMARAT, College of Medicine, University of Ibadan

Thank you for choosing to participate in the study. Kindly show by using any of the following two boxes, that your participation in this study was voluntary.

I will participate	I will not participate

#### Focus group discussion guide

| S/N | **Main questions** | **Follow up questions or hints** |
| --- | --- | --- |
|  | **Introduction and general questions** | |
| 1 | a) What can you say about the living conditions of community members? | - Settlements or housing conditions in the community   - (**Probe for**, if not mentioned, presence of formal settlements, informal settlements, slums)   - (Probe for their opinions about features of formal settlements, informal settlements, slums) - Financial condition or status of community members   - (**Probe for**, opinions about categorization of community members based on economic or financial status) - Health facilities in the communities - **Probe for**, availability of the health facilities in the communities - **Probe for**, types of health facilities most preferred by community members (private hospital, primary health facilities, secondary health facilities, tertiary health facilities) - **Probe for**, characteristics of preferred health facilities |
|  | b)Where does the community members generally prefer to go for health care? | **Probe** for the following (if not mentioned):   - Health facilities (private hospitals, primary health facility, secondary health facilities, tertiary health facilities). Probe for names of the preferred facilities - Patent medicine stores - Traditional healing home - Drug peddlers) |
|  | c) In this community if someone is pregnant, where would they typically go for antenatal care? | **Probe** for the following (if not mentioned):   - Health facilities (private hospitals, primary health facility, secondary health facilities, tertiary health facilities). Probe for names of the preferred facilities - Traditional Birth Attendants - Traditional doctors - Faith-based maternity homes - Nowhere/prefer to give birth at home - Opinions on why some pregnant women prefer not to attend antenatal care in the hospitals |
|  | d) What are the common diseases in this community | - Which diseases are most common among under-five children? - Which diseases are most common among adult population? - Which diseases are the most severe diseases? - Which diseases are associated with mosquito bites in the community? |
|  | **Basic understanding of malaria** | |
| 2 | Now let us discuss specifically on some basic issues relating to malaria.  a) How common is malaria in this community? | What categories of persons are most affected by malaria?  Probe for:   - Under-five children - Adolescents - Adults - Aged people - Pregnant women - People with sickle cell anemia etc. |
|  | b) How do community members usually get to know that they have malaria? | **Probe** for the following (if not mentioned):   - Through observation of signs and symptoms that are suggestive of malaria - Clinical examination - Lab examination |
|  | c) In your opinion, what are the signs and symptoms of malaria | **______________________________** |
|  | d) Please share with us what you think is the cause of malaria? | **Probe** for the following:   - Mosquito bite - Exposure to hot sunshine - Poor hygiene - Hunger - Spiritual reasons etc. |
|  | **Practices relating to management of malaria in the community** | |
| 3 | a) How do adult members in community manage suspected malaria infections? | **Probe** for the following (if not mentioned):   - Home-based care - Self-medication - Use of herbs - Use of drugs or medicines with prescription |
|  | b) Tell us about how suspected malaria infections in under-five children are being managed in the community | **Probe** for the following (if not mentioned):   - Home-based care - Self-medication - Use of herbs - Use of drugs or medicines with prescription |
|  | c) Tell us about how suspected malaria infections in adolescents are being managed in the community | **Probe for the following (if not mentioned):**   - Home-based care - Self-medication - Use of herbs - Use of drugs or medicines with prescription |
|  | d) If you think someone in your community has malaria, what kind of treatment should they follow? | **Probe** for the following (if not mentioned):   - Home-based care - Self-medication - Use of herbs - Use of drugs or medicines with prescription |
|  | e) Kindly share your experience of how you managed your last or most recent malaria episode | **Probe for**,   - The malaria medication(s) used the last time participants had malaria infection |
|  | **Malaria related health-seeking behaviours of community members** | |
| 4 | Where do community members seek care for malaria? | **Probe** for the following (if not mentioned)   - Traditional healing homes - Patient Medicine Vendors stores - Drug hawkers/peddlers - Pharmacy stores - Herbal drug stores/kiosks - Hospitals (Private hospital, government hospital)   **Probe for** where community members seek care for malaria the most and reasons  **Probe for** where participants treated their own last malaria episodes |
| 5 | What can you say about what influences community members to seek malaria treatment in a hospital? | **Probe** for the following   - Money for paying bills for treatment - Money for transportation to hospital - Distance of hospital - Access to hospital - Availability of hospital - Availability of medications - Waiting time in the hospital/health facility - Attitude of health workers - Preference for traditional medicine - Preference for a particular type of hospital (Private hospital, primary health centre, secondary health care facility) - Cultural norms and beliefs about seeking care in hospital - Cultural norms and beliefs malaria - Perceived seriousness of malaria - Perceived threat of malaria - Frequency of malaria episode |
| 6 | What can you say about what influences community members to seek malaria treatment in a particular hospital/health facility? | Probe for the following   - Attitude of the health workers - Reputation of the provider - Previous experience with provider - Gender of the provider - Distance to the health facility - Quality of services - Ambience of health facility - Waiting times - Availability of medication - Cost of services |
| 7 | Let us briefly discuss how health seeking behaviour and source of care differ by socioeconomic group? | **Probe** for –where each of the following categories of people usually seek malaria care?  **‘**   - Rich people in the community - Poor people in the community - People with formal education - People without formal education   (**For each of the categories, probe for the common source of malaria treatment e.g** - government institutions, private clinics, pharmacy, chemists, drug peddlers/hawkers, traditional healing homes, herbal drug stores/kiosks) |
|  | **Malaria medications being used and their local names** | |
| 8 | a) What can you say about the malaria medications that community members use? | **Probe** for   - Please tell us about malaria medications that are commonly used among adult population in the community - Tell us about the malaria medications that are commonly used among children in the community - What are your views about the cost of the malaria medications |
|  | b) What are the local names for the malaria medications that community members use? |  |
|  | **Showing of pictures or physical packaging of malaria medications to improve recall and answers about usage of malaria medication** | |
| 9 | a) What are your opinions about the showing of pictures or physical packaging of malaria medications with the intention of improving recall and answers about usage of malaria medication? | **Probe** for   - Why they think showing of pictures or physical packaging of malaria medications can possibly improve recall and answers about usage of malaria medication? |
|  | b) In your opinion what other ways or means do you think can help community members to improve recall and answer about usage of malaria medications? |  |
|  | **Factors influencing use of malaria medications and treating malaria at a hospital** | |
| 10 | a) Please share with us the things that do influence community members decisions about choice of malaria medications | **Probe** for   - **Individual factors that influence the decisions about what malaria medicine to use – e.g.,** Age, sex (male or female), marital status, economic status, being pregnant, medical history, perceived seriousness of malaria, perceived threat of malaria, preference for particular drug - **Drug related factors that influence the decisions about what malaria medicine to use –** e.g. Availability of drug in hospital or drug stores, number of days expected to use drugs, number of capsules/dosages, taste of drug, side effects of drugs, drug resistance, proliferation of fake malaria medicines, giving of genuine malaria medications - **Social-cultural factors that influence the decisions about what malaria medicine to use –**e.g support from partners, support family members, support from friends/peers, media influence and drug adverts, cultural norms, and values - **Health system related factors that influence the decisions about what malaria medicine to use-**e.g availability of drugs in hospital/pharmacy, prescription of drug, advice from health workers - **Policy related factors e.g**–governments and regulatory bodies recommendation relating to antimalarial drugs that should be first line of treatment - **Disease pattern related factors e.g**–uncomplicated vs severe malaria - **Economic related factors -** e.g – cost of buying malaria medications, affordability of preferred malaria medications |
|  | a) What are the foremost things or situations that do influence your own decisions about the use of malaria medications? | **Probe for:**   - Cost of treatment in private hospitals - Cost of treatment in primary health facilities - Cost of treatment in secondary health facilities - Cost of treatment in patent medicine store - Cost of treatment using traditional/herbal medicines |
|  | b) What can you say about the cost of treating malaria in the hospital? | - How affordable do you think the cost of treating malaria is for community members in the health facility? |
|  | **Common methods that community members use to protect themselves from malaria** | |
| 11 | a) Please tell us about the common methods that people use to protect themselves from malaria | **Probe** for (if not mentioned):   - Use of insecticide sprays - Use of insecticides treated bed nets - Use of mosquito replants - Use of coils - Use of window and door screens - Wearing of long-sleeved clothing and long pants - Malaria prophylaxis |
|  | **b)** Which method is most commonly used by people to protect themselves from malaria? | **Probe** for   - Reasons for being the most commonly used |
|  | **c)** Which methods do community members usually use to protect under-five children from malaria? |  |
|  | **Participation in community-based malaria programme** | |
| 12 | a) We are planning to put in place a free community-based programme whereby community members will be asked to consistently report malaria cases or symptoms. Our goal is to understand the transmission of malaria in the community to inform where interventions go. What is your opinion about how community members will perceive the programme? |  |
|  | b) If we want community members to be reporting malaria cases or symptoms consistently what would be the preferred way or means for this? | **Probe** for (if not mentioned):   - Face-to-face with a project volunteer - Through community representative - Through SMS - Through WhatsApp - Through mobile application - Through hotline   Through community meeting etc. |
|  | c) Supposed we ask community members to be reporting malaria cases or symptoms via text message as part of our community-based malaria programme, what is your opinion about it? | **Probe** for:   - How easy it will be for community members to participate - How willing community members will be - The barriers that may be associated with it - What can encourage community members to be reporting consistently malaria cases or symptoms via text message - Participants’ willingness to participate to be involved |
|  | d) What is your opinion about asking community members to be reporting malaria cases or symptoms via a mobile application as part of our community-based malaria programme? | **Probe** for:   - How easy it will be for community members to participate - How willing community members will be - The barriers that may be associated with it - What can encourage community members to be reporting consistently malaria cases or symptoms via a mobile application - Participants’ willingness to participate to be involved |
|  | e) What can facilitate the participation of community members in the programme? |  |
|  | **Conclusion and other relevant information** | |
| 13 | a) Please tell us about on-going or existing community-based malaria interventions in this community |  |
|  | b) What suggestions do you have about how malaria can be controlled in this community? |  |
|  | c) What other suggestions do you have that can help us with the community-based malaria programme that we are planning? |  |

**Socio-demographic information**

Ward and LGA………………………………	Name of Community/Area…………………

Age in years (at last birthday) …………	Sex.……………………

Highest level of Education………………	Primary Occupation……………………

Type of community/settlement ………………………………….

#### Key informant interview consent form for formal health sector providers

*Who to interview*: Formal Healthcare Sector Providers - These include:

1. Heads of Primary Health Care (PHC) Centres/facilities
2. PHC coordinators /Medical officer of Health at LGA level
3. Roll-back malaria focal persons at LGA level
4. Pharmacists and doctors working in private and public health facilities
5. Malaria programme officers at State level

*Introduction*

My name is ……………and my colleagues are…………… We work with the University of Ibadan. We would like your opinion on various issues to enable us understand malaria transmission in urban areas as part of a collaborative project with Northwestern University, USA and Nigeria National Malaria Elimination Programme. The information we collect will help the government to plan health services to prevent malaria infections by ensuring you receive suitable interventions. You have been specially invited for this key informant interview and we thank you for honouring our invitation.

*Purpose*

The purpose of this key informant interview is to investigate the following

1. How community members manage suspected malaria infections
2. Where community members seek care for malaria
3. Factors that influence care seeking for malaria in a hospital,
4. How health seeking behaviour and source of care differ by socioeconomic group
5. Local names for malaria medications
6. Whether showing of pictures or physical packaging of malaria medications can improve participants’ recall and answers about usage of malaria medication
7. Factors that influence decisions about what malaria medicine to use
8. Common methods that people use to protect themselves from themselves from malaria
9. Understand facilitators and barriers to participating in a community-based disease reporting programme for malaria

The information learned in this key informant interview will be used to guide the development of other components of the study that we are planning to conduct.

*Procedure*

The key informant interview, which will last for 40 – 60 minutes, will entail asking you some open-ended questions. In addition to the interview questions, we will be asking for some of your socio-demographic information. During this interview, your views will be respected and will not be used against you in any way. This interview will be taped, so please speak up and speak clearly. We ask for your consent to record the interview so that we will not miss out anything from the information you will be providing to us through the interview. Please do share your views without mentioning people’s names. We want the interview to be anonymous and to be confidential as possible. You can choose whether to participate in the interview, and you may stop at any time during the study. There is no right or wrong view, so feel free to express yourself. Please note that your participation in this interview is voluntary. Your decision not to be involved or drop out at any point will not attract any penalty.

*Benefits*

Your participation in this study may not provide any personal benefit to you. However, should you decide to participate in this study, you will be doing society a great service because the findings of this study will be useful in the design of interventions and programmes for the control and prevention of malaria.

*Risks*

There are no known or anticipated risks associated with participation in this study beyond those experienced during an average conversation. If a question, or the discussion, makes you uncomfortable, you can choose not to answer.

*Confidentiality*

The information you share will be kept confidential. Identifying information will be removed from the transcripts. The transcripts and other electronic data will be retained for a maximum of 5 years, after which they will be destroyed. Data will be stored in an encrypted folder on protected laptop. Only the research team will have access to study data. No identifying information will be used in any presentations or publications based on this research.

*Contact*

If you have any questions or concerns regarding this study, please contact:

Professor IkeOluwapo Ajayi

Director, Institute for Advanced Medical Research and Training (IAMRAT),

College of Medicine, University of Ibadan

Thank you for choosing to participate in the study. Kindly show by using any of the following 2 boxes, that your participation in this study was voluntary.

I will participate	I will not participate

#### Key informant interview guide for formal health sector providers

| S/N | **Main questions** | **Follow up questions or hints** |
| --- | --- | --- |
|  | **Introduction and general questions** | |
| 1 | a) Please tell us about the health facilities that are available in this community | **Probe for:**   - Primary health centres - Secondary health facilities - tertiary health facilities - Private hospitals |
|  | b)Where does the community members generally prefer to go for health care? | **Probe**-(if not mentioned):   - Health facilities (private hospitals, primary health facility, secondary health facilities, tertiary health facilities). - Probe for, opinion about the types of health facilities most preferred by community members - Patent medicine stores - Traditional healing home - Drug peddlers) |
|  | c) In this community if someone is pregnant, where would they typically go for antenatal care? | **Probe** for the following (if not mentioned):   - Health facilities (private hospitals, primary health facility, secondary health facilities, tertiary health facilities). Probe for names of the preferred facilities - Traditional Birth Attendants - Traditional doctors - Faith-based maternity homes - Nowhere/prefer to give birth at home - Opinions on why some pregnant women prefer not to attend antenatal care in the hospitals |
|  | d) What are the common diseases in this community | - Which diseases are most common among under-five children? - Which diseases are most common among adult population? - Which diseases are the most severe diseases? |
|  | **Basic understanding of malaria** | |
| 2 | Now let us discuss specifically on some basic issues relating to malaria.  a)How common is malaria in this community? | What categories of persons are most affected by malaria?  Probe for:   - Under-five children - Aged people - Pregnant women - People with sickle cell anemia etc. |
|  | b) How do community members usually get to know that they have malaria? | **Probe** for the following (if not mentioned):   - Through observation of signs and symptoms that are suggestive of malaria - Clinical examination - Lab examination |
|  | c) In your opinion, what are the signs and symptoms of malaria |  |
|  | **Practices relating to management of malaria in the community** | |
| 3 | a) How do adult members in community manage suspected malaria infections? | **Probe** for the following (if not mentioned):   - Home-based care - Self-medication - Use of herbs - Use of drugs or medicines with prescription |
|  | b) Tell us about how suspected malaria infections of under-five children are being managed in the community? | **Probe** for the following (if not mentioned):   - Home-based care - Self-medication - Use of herbs - Use of drugs or medicines with prescription |
|  | c) Tell us about how suspected malaria infections in adolescents are being managed in the community | **Probe** for the following (if not mentioned):   - Home-based care - Self-medication - Use of herbs - Use of drugs or medicines with prescription |
|  | d) Kindly describe to us how malaria infection is being managed in the health facility |  |
|  | **Malaria related health-seeking behaviours of community members** | |
| 4 | Where do community members seek care for malaria? | **Probe** for the following (if not mentioned)   - Traditional healing homes - Patient Medicine Vendors stores - Drug hawkers/peddlers - Pharmacy stores - Herbal drug stores/kiosks - Hospitals (Private hospital, government hospital)   **Probe for** where community members seek care for malaria the most and reasons |
| 5 | What do you think usually influence community members to seek malaria in a hospital? | **Probe** for the following   - Money for paying bills for treatment - Money for transportation to hospital - Distance of hospital - Access to hospital - Availability of hospital - Availability of medications - Waiting time in the hospital/health facility - Attitude of health workers - Preference for traditional medicine - Preference for a particular type of hospital (Private hospital, primary health centre, secondary health care facility) - Cultural norms and beliefs about seeking care in hospital - Cultural norms and beliefs malaria - Perceived seriousness of malaria - Perceived threat of malaria - Frequency of malaria episode |
| 6 | Let us briefly discuss how health seeking behaviour and source of care differ by socioeconomic group? | **Probe** for –where each of the following categories of people usually seek malaria care?  **‘**   - Rich people in the community - Poor people in the community - People with formal education - People without formal education   (**For each of the categories, probe for the common source of malaria treatment e.g.** - government institutions, private clinics, pharmacy, chemists, drug peddlers/hawkers, traditional healing homes, herbal drug stores/kiosks) |
|  | **Malaria medications being used and their local names** | |
| 7 | a)What can you say about the malaria medications that community members use? | - Please tell us about malaria medications that are commonly used among adult population in the community. - **Probe for**, the local names for the malaria medications being used by adults in the community - Tell us about the malaria medications that are commonly used among children in the community. - **Probe for**, the local names for the malaria medications being used by children in the community - What are your views about the cost of the malaria medications? - How affordable do you think the cost of malaria medications is for community members? |
|  | b)Please think about it if you were sick, which malaria medications would you take | **Probe for**,   - The local names for the malaria medications - The malaria medication(s) used the last time participant had malaria infection |
|  | **Showing of pictures or physical packaging of malaria medications to improve recall and answers about usage of malaria medication** | |
| 8 | a)What are your opinions about the showing of pictures or physical packaging of malaria medications with the intention of improving recall and answers about usage of malaria medication? | **Probe** for:   - Why showing of pictures or physical packaging of malaria medications can possibly improve recall and answers about usage of malaria medication? |
|  | b) In your opinion what other ways or means do you think can help community members to improve recall and answer about usage of malaria medications? |  |
|  | **Factors influencing use of malaria medications and treating malaria at a hospital** | |
| 9 | a)Please share with us the things that you think do influence community members decisions about choice of malaria medications | **Probe** for   - **Individual factors that influence the decisions about what malaria medicine to use – e.g.,** Age, sex (male or female), marital status, economic status, being pregnant, medical history, perceived seriousness of malaria, perceived threat of malaria, preference for particular drug type - **Drug related factors that influence the decisions about what malaria medicine to use –** e.g., Availability of drug in hospital or drug stores, number of days expected to use drugs, number of capsules/dosages, taste of drug, side effects of drugs, drug resistance, proliferation of fake malaria medicines, recognizing genuine malaria medications - **Social-cultural factors that influence the decisions about what malaria medicine to use –** e.g. support from partners, support family members, support from friends/peers, media influence and drug adverts, cultural norms, and values - **Health system related factors that influence the decisions about what malaria medicine to use-** e.g., availability of drugs in hospital/pharmacy, prescription of drug, advice from health workers - **Policy related factors e.g.**–governments and regulatory bodies recommendation relating to antimalarial drugs that should be first line of treatment - **Disease pattern related factors e.g.**–uncomplicated vs severe malaria - **Economic related factors** e.g. – cost of buying malaria medications, affordability of preferred malaria medications |
|  | c) What is your view about the cost of treating malaria in the hospital/health facility? | - How affordable do you think the cost of treating malaria is for community members in the health facility or hospitals? |
|  | **Common methods that community members use to protect themselves from malaria** | |
| 10 | a) Please tell us about the common methods that people use to protect themselves from malaria in this community | **Probe** for (if not mentioned):   - Use of insecticide sprays - Use of insecticides treated bed nets - Use of mosquito replants - Use of coils - Use of window and door screens - Wearing of long-sleeved clothing and long pants - Malaria prophylaxis |
|  | b) Which method is most commonly used by people to protect themselves from malaria? | **Probe** for   - Reasons for being the most commonly used |
|  | c) Which methods do community members usually use to protect under-five children from malaria? |  |
|  | **Participation in community-based malaria programme** | |
| 11 | a) We are planning to put in place a free community-based programme whereby community members will be asked to consistently malaria cases or symptoms. Our goal is to understand the transmission of malaria in the community to inform where interventions go. What is your opinion about how community members will perceive the programme. |  |
|  | b) If we want community members to be reporting malaria cases or symptoms consistently what do you think would be the preferred way or means for this? | **Probe** for (if not mentioned):   - Face-to-face with a project volunteer - Through community representative - Through SMS - Through WhatsApp - Through mobile application - Through hotline   Through community meeting etc. |
|  | c) Supposed we ask community members to be reporting malaria cases or symptoms via text message as part of our community-based malaria programme, what is your opinion about it? | **Probe** for:   - How easy do you think it will be for community members to participate? - How willing do you think community members will be? - What barriers do you think may be associated with it? - What do you think we can encourage community members to be reporting consistently malaria cases or symptoms via text message? |
|  | d) What is your opinion about asking community members to be reporting malaria cases or symptoms via a mobile application as part of our community-based malaria programme? | **Probe** for:   - How easy do you think it will be for community members to participate? - How willing do you think community members will be? - What barriers do you think may be associated with it? - What do you think we can encourage community members to be reporting consistently malaria cases or symptoms via a mobile application? |
|  | e) What can facilitate the participation of community members in the programme? |  |
|  | **Conclusion and other relevant information** | |
| 12 | a) Please tell us about on-going or existing community-based malaria interventions in this community |  |
|  | b) What suggestions do you have about how malaria can be controlled in this community? |  |
|  | c) What other suggestions do you have that can help us with the community-based malaria programme that we are planning? |  |

**Socio-demographic information**

Ward and LGA ………………………………	Sex: ……………………

Age in years (at last birthday) …………	Highest level of Education………………

Designation ……………………	Position ……………………

Number of years spent in present position ……………………

Number of years spent in health facility……………………….

Type of settlement ……………………………….………….......

Name of Community/Area……………………………………….

#### Key informant interview consent form for informal health care workers

*Who to interview*: Informal Healthcare Providers- These include:

1. Patient Medicine Vendors
2. Drug peddlers/hawkers
3. Herbal drug sellers
4. Traditional doctors

*Introduction*

My name is ……………and my colleagues are…………… We work with the University of Ibadan. We would like your opinion on various issues to enable us understand malaria transmission in urban areas as part of a collaborative project with Northwestern University, USA and Nigeria National Malaria Elimination Programme. The information we collect will help the government to plan health services to prevent malaria infections by ensuring you receive suitable interventions. You have been specially invited for this key informant interview and we thank you for honouring our invitation.

*Purpose*

The purpose of this key informant interview is to investigate the following

1. How community members manage suspected malaria infections
2. Where community members seek care for malaria
3. Factors that influence care seeking for malaria in a hospital,
4. How health seeking behaviour and source of care differ by socioeconomic group
5. Local names for malaria medications
6. How showing of pictures or physical packaging of malaria medications can improve participants’ recall and answers about usage of malaria medication
7. Factors that influence decisions about what malaria medicine to use
8. Common methods that people use to protect themselves from themselves from malaria
9. Understand facilitators and barriers to participating in a community-based disease reporting programme for malaria

The information learned in this key informant interview will be used to guide the development of other components of the study that we are planning to conduct.

*Procedure*

The key informant interview, which will last for 40 – 60 minutes, will entail asking you some open-ended questions. In addition to the interview questions, we will be asking for some of your socio-demographic information. During this interview, your views will be respected and will not be used against you in any way. This interview will be taped, so please speak up and speak clearly. We ask for your consent to record the interview so that we will not miss out anything from the information you will be providing to us through the interview. Please do share your views without mentioning people’s names. We want the interview to be anonymous and to be confidential as possible. You can choose whether to participate in the interview, and you may stop at any time during the study. There is no right or wrong view, so feel free to express yourself. Please note that your participation in this interview is voluntary. Your decision not to be involved or drop out at any point will not attract any penalty.

*Benefits*

Your participation in this study may not provide any personal benefit to you. However, should you decide to participate in this study, you will be doing society a great service because the findings of this study will be useful in the design of interventions and programmes for the control and prevention of malaria.

*Risks*

There are no known or anticipated risks associated with participation in this study beyond those experienced during an average conversation. If a question, or the discussion, makes you uncomfortable, you can choose not to answer.

*Confidentiality*

The information you share will be kept confidential. Identifying information will be removed from the transcripts. The transcripts and other electronic data will be retained for a maximum of 5 years, after which they will be destroyed. Data will be stored in an encrypted folder on protected laptop. Only the research team will have access to study data. No identifying information will be used in any presentations or publications based on this research.

*Contact*

If you have any questions or concerns regarding this study, please contact:

Professor IkeOluwapo Ajayi

Director, Institute for Advanced Medical Research and Training (IMARAT),

College of Medicine, University of Ibadan

Thank you for choosing to participate in the study. Kindly show by using any of the following 2 boxes, that your participation in this study was voluntary.

I will participate	I will not participate

#### Key informant interview guide for informal health care workers

| S/N | **Main questions** | **Follow up questions or hints** |
| --- | --- | --- |
|  | **Introduction and general questions** | |
| 1 | a)What can you say about the living conditions of community members? | - Financial condition or status of community members   - (**Probe for**, opinions about categorization of community members based on economic or financial status) - Health facilities available in the communities - **Probe for**, types of health facilities (private hospital, primary health facilities, secondary health facilities, tertiary health facilities) - **Probe for**, the most preferred by community members (private hospital, primary health facilities, secondary health facilities, tertiary health facilities) |
|  | b)Where does the community members generally prefer to go for health care? | **Probe**-(if not mentioned):   - Health facilities (private hospitals, primary health facility, secondary health facilities, tertiary health facilities). - Patent medicine stores - Traditional healing home - Drug peddlers etc. |
|  | c) In this community if someone is pregnant, where would they typically go for antenatal care? | **Probe** for the following (if not mentioned):   - Health facilities (private hospitals, primary health facility, secondary health facilities, tertiary health facilities). - Traditional Birth Attendants - Traditional doctors - Faith-based maternity homes - Nowhere/prefer to give birth at home - Opinions on why some pregnant women prefer not to attend antenatal care in the hospitals |
|  | d) What are the common diseases in this community | - Which diseases are most common among under-five children? - Which diseases are most common among adult population? - Which diseases are the most severe diseases? |
|  | **Basic understanding of malaria** | |
| 2 | Now let us discuss specifically on some basic issues relating to malaria.  a)How common is malaria in this community? | What categories of persons are most affected by malaria?  Probe for:   - Under-five children - Aged people - Pregnant women - People with sickle cell anemia etc. |
|  | b) How do community members usually get to know that they have malaria? | **Probe** for the following (if not mentioned):   - Through observation of signs and symptoms that are suggestive of malaria - Clinical examination - Lab examination |
|  | c) In your opinion, what are the signs and symptoms of malaria |  |
|  | **Practices relating to management of malaria in the community** | |
| 3 | a) How do adult members in community manage suspected malaria infections? | **Probe** for the following (if not mentioned):   - Home-based care - Self-medication - Use of herbs - Use of drugs or medicines with prescription |
|  | b) Tell us about how suspected malaria infections of under-five children are being managed in the community? | **Probe** for the following (if not mentioned):   - Home-based care - Self-medication - Use of herbs - Use of drugs or medicines with prescription |
|  | c) Tell us about how suspected malaria infections in adolescents are being managed in the community | **Probe for the following (if not mentioned):**   - Home-based care - Self-medication - Use of herbs - Use of drugs or medicines with prescription |
|  | **(For herbal drug sellers and traditional doctors)**  d) Kindly describe to us how malaria infection should be managed or treated using herbal malaria drugs? | **Probe for:**   - Malaria infections involving adults - Malaria infections involving under-five children - Malaria infection involving pregnant women - Complicated malaria infections - Individuals having frequent malaria episodes |
|  | **(For Patient Medicine Vendors and Drug peddlers/hawkers involved in selling over-the-counter malaria medicines)**  d ii) Kindly describe to us how malaria infection should be managed or treated using malaria medicines (orthodox medicine)? | **Probe for:**   - Malaria infections involving adults - Malaria infections involving under-five children - Malaria infection involving pregnant women - Complicated malaria infections - Individuals having frequent malaria episodes |
|  | **Malaria related health-seeking behaviours of community members** | |
| 4 | Where do community members seek care for malaria? | **Probe** for the following (if not mentioned)   - Traditional healing homes - Patient Medicine Vendors stores - Drug hawkers/peddlers - Pharmacy stores - Herbal drug stores/kiosks - Hospitals (Private hospital, government hospital)   **Probe for** where community members seek care for malaria the most and reasons |
| 5 | a) What do you think usually influence community members to seek malaria care in a hospital? | **Probe** for the following   - Money for paying bills for treatment - Money for transportation to hospital - Distance of hospital - Access to hospital - Availability of hospital - Attitude of health workers - Preference for traditional medicine - Preference for a particular type of hospital (Private hospital, primary health centre, secondary health care facility) - Cultural norms and beliefs about seeking care in hospital - Cultural norms and beliefs malaria - Perceived seriousness of malaria - Perceived threat of malaria - Frequency of malaria episode |
|  | b)What do you think usually influence community members to seek malaria treatment in Patient Medicine Vendors stores or from drug peddlers?  **(For Patient Medicine Vendors and Drug peddlers/hawkers involved in selling over-the-counter malaria medicines)** | **Probe** for the following   - Money for paying bills for treatment - Money for transportation to hospital - Distance of hospital - Access to hospital - Availability of hospital - Attitude of health workers - Cultural norms and beliefs about seeking care in hospital - Cultural norms and beliefs malaria - Perceived seriousness of malaria - Perceived threat of malaria - Frequency of malaria episode |
|  | c)What do you think usually influence community members to seek malaria in a hospital?  **(For herbal drug sellers and traditional doctors)** | **Probe** for the following   - Money for paying bills for treatment - Money for transportation to hospital - Distance of hospital - Access to hospital - Availability of hospital - Attitude of health workers - Preference for traditional medicine - Cultural norms and beliefs about seeking care in hospital - Cultural norms and beliefs malaria - Perceived seriousness of malaria - Perceived threat of malaria - Frequency of malaria episode |
| 6 | Let us briefly discuss how health seeking behaviour and source of care differ by socioeconomic group? | **Probe** for –where each of the following categories of people usually seek malaria care?   - Rich people in the community - Poor people in the community - People with formal education - People without formal education   (**For each of the categories, probe for the common source of malaria treatment e.g** - government institutions, private clinics, pharmacy, chemists, drug peddlers/hawkers, traditional healing homes, herbal drug stores/kiosks |
|  | **Malaria medications being used and their local names** | |
| 7 | a)What can you say about the malaria medications that community members use? | - Please tell us about malaria medications that are commonly used among adult population in the community. - **Probe for**, the local names for the malaria medications being used by adults in the community - Tell us about the malaria medications that are commonly used among children in the community. - **Probe for**, the local names for the malaria medications being used by children in the community - What are your views about the cost of the malaria medications (orthodox medicine)? - How affordable do you think the cost of malaria medications is for community members? |
|  | b)What can you say about the use of herbal drugs for treating malaria?  **(For herbal drug sellers and traditional doctors)** | - Types of herbal drugs used for treating malaria - **Probe for**, the local names for the herbal malaria drugs - Perceived efficacy of the herbal drugs used for treating malaria - How common the use of herbal malaria drugs is in the community - Characteristics of community women who prefer to use herbal drugs for malaria treatment - Reasons community members prefer to use herbal drugs for malaria treatment - What are your views about the cost of the herbal malaria drugs? - What determines the type of herbal malaria drugs that you give to your patients? - Under what condition should community members who use herbal malaria drugs seek healthcare in the hospital for malaria treatment? |
|  | c)What can you say about your competency to treat and manage malaria infections? | **Probe** for the competency to manage:   - Malaria infections involving adults - Malaria infections involving under-five children - Malaria infection involving pregnant women - Complicated malaria infections - Individuals having frequent malaria episodes |
|  | **Showing of pictures or physical packaging of malaria medications to improve recall and answers about usage of malaria medication** | |
| 8 | a)What are your opinions about the showing of pictures or physical packaging of malaria medications with the intention of improving recall and answers about usage of malaria medication? | **Probe** for   - Why showing of pictures or physical packaging of malaria medications can possibly improve recall and answers about usage of malaria medication? |
|  | b) In your opinion what other ways or means do you think can help community members to improve recall and answer about usage of malaria medications? |  |
|  | **Factors influencing use of malaria medications and treating malaria at a hospital** | |
| 9 | a)Please share with us the things that you think do influence community members decisions about choice of malaria medications | **Probe** for   - **Individual factors that influence the decisions about what malaria medicine to use – e.g** Age, sex (male or female), marital status, economic status, being pregnant, medical history, perceived seriousness of malaria, perceived threat of malaria, preference for particular type of malaria medications (orthodox drugs), preference for herbal malaria drugs - **Drug related factors that influence the decisions about what malaria medicine to use –** e.g Availability of drug in hospital or drug stores, number of days expected to use drugs, number of capsules/dosages, taste of drug, side effects of drugs, drug resistance, availability of herbal malaria drugs, preference for herbal malaria drugs, proliferation of fake malaria medicines, giving of genuine malaria medications - **Social-cultural factors that influence the decisions about what malaria medicine to use –** e.g support from partners, support family members, support from friends/peers, media influence and drug adverts, cultural norms, and values - **Health system related factors that influence the decisions about what malaria medicine to use-** e.g., availability of drugs in hospital/pharmacy, prescription of drug, advice from health workers - **Policy related factors e.g.** – governments and regulatory bodies recommendation relating to antimalarial drugs that should be first line of treatment - **Disease pattern related factors e.g.**–uncomplicated vs severe malaria - **Economic related factors** e.g. – cost of buying malaria medications, affordability of preferred malaria medications |
|  | d) What is your view about the cost of treating malaria in the hospital? | - How affordable do you think the cost of treating malaria is for community members in the health facility? |
|  | **Common methods that community members use to protect themselves from malaria** | |
| 10 | a) Please tell us about the common methods that people use to protect themselves from malaria in this community | **Probe** for (if not mentioned):   - Use of insecticide sprays - Use of insecticides treated bed nets - Use of mosquito replants - Use of coils - Use of window and door screens - Wearing of long-sleeved clothing and long pants - Malaria prophylaxis |
|  | b) Which method is most commonly used by people to protect themselves from malaria? | **Probe** for   - Reasons for being the most commonly used |
|  | c) Which methods do community members usually use to protect under-five children from malaria? |  |
|  | **Participation in community-based malaria programme** | |
| 11 | a) We are planning to put in place a free community-based programme where community members will be asked to consistently malaria cases or symptoms. Our goal is to understand the transmission of malaria in the community to inform where interventions go. What is your opinion about how community members will perceive the programme |  |
|  | b) If we want community members to be reporting malaria cases or symptoms consistently what do you think would be the preferred way or means for this? | **Probe** for (if not mentioned):   - Face-to-face with a project volunteer - Through community representative - Through SMS - Through WhatsApp - Through mobile application - Through hotline   Through community meeting etc. |
|  | c) Supposed we ask community members to be reporting malaria cases or symptoms via text message as part of our community-based malaria programme, what is your opinion about it? | **Probe** for:   - How easy do you think it will be for community members to participate? - How willing do you think community members will be? - What barriers do you think may be associated with it? - What do you think we can encourage community members to be reporting consistently malaria cases or symptoms via text message? |
|  | d) What is your opinion about asking community members to be reporting malaria cases or symptoms via a mobile application as part of our community-based malaria programme? | **Probe** for:   - How easy do you think it will be for community members to participate? - How willing do you think community members will be? - What barriers do you think may be associated with it? - What do you think we can encourage community members to be reporting consistently malaria cases or symptoms via a mobile application? |
|  | e) What can facilitate the participation of community members in the programme? |  |
|  | **Conclusion and other relevant information** | |
| 12 | a) Please tell us about on-going or existing community-based malaria interventions in this community |  |
|  | b) What suggestions do you have about how malaria can be controlled in this community? |  |
|  | c) What other suggestions do you have that can help us with the community-based malaria programme that we are planning? |  |

**Socio-demographic information**

Ward and LGA ………………………………	Sex: ……………………

Age in years (at last birthday) …………	Highest level of Education………………

Primary occupation ……………………	Other occupation(s) ……………………..

Number of years spent as informal healthcare provider……………………

Type of settlement ……………………………….…………..…………….

Name of Community/Area…………………………………….…………...

#### Key informant interview consent form for community leaders

*Who to interview*: Community leaders - These include:

1. Opinion leaders
2. Traditional leaders
3. Religious leaders
4. Women leaders

*Introduction*

My name is ……………and my colleagues are…………… We work with the University of Ibadan. We would like your opinion on various issues to enable us understand malaria transmission in urban areas as part of a collaborative project with Northwestern University, USA and Nigeria National Malaria Elimination Programme. The information we collect will help the government to plan health services to prevent malaria infections by ensuring you receive suitable interventions. You have been specially invited to this key informant interview and we thank you for honouring our invitation.

*Purpose*

The purpose of this key informant interview is to investigate the following

1. How community members manage suspected malaria infections
2. Where community members seek care for malaria
3. Factors that influence care seeking for malaria in a hospital,
4. How health seeking behaviour and source of care differ by socioeconomic group
5. Local names for malaria medications
6. How showing of pictures or physical packaging of malaria medications can improve participants’ recall and answers about usage of malaria medication
7. Factors that influence decisions about what malaria medicine to use
8. Common methods that people use to protect themselves from themselves from malaria
9. Understand facilitators and barriers to participating in a community-based disease reporting programme for malaria

The information learned in this key informant interview will be used to guide the development of other components of the study that we are planning to conduct.

*Procedure*

The key informant interview, which will last for 40 – 60 minutes, will entail asking you some open-ended questions. In addition to the interview questions, we will be asking for some of your socio-demographic information. During this interview, your views will be respected and will not be used against you in any way. This interview will be taped, so please speak up and speak clearly. We ask for your consent to record the interview so that we will not miss out anything from the information you will be providing to us through the interview. Please do share your views without mentioning people’s names. We want the interview to be anonymous and to be confidential as possible. You can choose whether to participate in the interview, and you may stop at any time during the course of the study. There is no right or wrong view, so feel free to express yourself. Please note that your participation in this interview is voluntary. Your decision not to be involved or drop out at any point will not attract any penalty.

*Benefits*

Your participation in this study may not provide any personal benefit to you. However, should you decide to participate in this study, you will be doing society a great service because the findings of this study will be useful in the design of interventions and programmes for the control and prevention of malaria.

*Risks*

There are no known or anticipated risks associated with participation in this study beyond those experienced during an average conversation. If a question, or the discussion, makes you uncomfortable, you can choose not to answer.

*Confidentiality*

The information you share will be kept confidential. Identifying information will be removed from the transcripts. The transcripts and other electronic data will be retained for a maximum of 5 years, after which they will be destroyed. Data will be stored in an encrypted folder on protected laptop. Only the research team will have access to study data. No identifying information will be used in any presentations or publications based on this research.

*Contact*

If you have any questions or concerns regarding this study, please contact:

Professor IkeOluwapo Ajayi

Director, Institute for Advanced Medical Research and Training (IMARAT),

College of Medicine, University of Ibadan

Thank you for choosing to participate in the study. Kindly show by using any of the following 2 boxes, that your participation in this study was voluntary.

I will participate	I will not participate

#### Key informant interview guide for community leaders

| S/N | **Main questions** | **Follow up questions or hints** |
| --- | --- | --- |
|  | **Introduction and general questions** | |
| 1 | a)What can you say about the living conditions of community members? | - Financial condition or status of community members   - (**Probe for**, opinions about categorization of community members based on economic or financial status) - Health facilities available in the communities - **Probe for**, types of health facilities (private hospital, primary health facilities, secondary health facilities, tertiary health facilities) - **Probe for**, the most preferred by community members (private hospital, primary health facilities, secondary health facilities, tertiary health facilities) |
|  | b)Where does the community members generally prefer to go for health care? | **Probe**-(if not mentioned):   - Health facilities (private hospitals, primary health facility, secondary health facilities, tertiary health facilities). - Patent medicine stores - Traditional healing home - Drug peddlers etc. |
|  | c) In this community if someone is pregnant, where would they typically go for antenatal care? | **Probe** for the following (if not mentioned):   - Health facilities (private hospitals, primary health facility, secondary health facilities, tertiary health facilities). - Traditional Birth Attendants - Traditional doctors - Faith-based maternity homes - Nowhere/prefer to give birth at home - Opinions on why some pregnant women prefer not to attend antenatal care in the hospitals |
|  | d) What are the common diseases in this community | - Which diseases are most common among under-five children? - Which diseases are most common among adult population? - Which diseases are the most severe diseases? |
|  | **Basic understanding of malaria** | |
| 2 | Now let us discuss specifically on some basic issues relating to malaria.  a)How common is malaria in this community? | What categories of persons are most affected by malaria?  Probe for:   - Under-five children - Aged people - Pregnant women - People with sickle cell anemia etc. |
|  | b) How do community members usually get to know that they have malaria? | **Probe** for the following (if not mentioned):   - Through observation of signs and symptoms that are suggestive of malaria - Clinical examination - Lab examination |
|  | c) In your opinion, what are the signs and symptoms of malaria |  |
|  | **Practices relating to management of malaria in the community** | |
| 3 | a) How do adult members in community manage suspected malaria infections? | **Probe** for the following (if not mentioned):   - Home-based care - Self-medication - Use of herbs - Use of drugs or medicines with prescription |
|  | b) Tell us about how suspected malaria infections of under-five children are being managed in the community? | **Probe** for the following (if not mentioned):   - Home-based care - Self-medication - Use of herbs - Use of drugs or medicines with prescription |
|  | c) Tell us about how suspected malaria infections in adolescents are being managed in the community | **Probe for the following (if not mentioned):**   - Home-based care - Self-medication - Use of herbs - Use of drugs or medicines with prescription |
|  | c) What efforts are being put in place by the community relating to the prevention and control of malaria? |  |
|  | **Malaria related health-seeking behaviours of community members** | |
| 4 | a) Where do community members seek care for malaria? | **Probe** for the following (if not mentioned)   - Traditional healing homes - Patient Medicine Vendors stores - Drug hawkers/peddlers - Pharmacy stores - Herbal drug stores/kiosks - Hospitals (Private hospital, government hospital)   **Probe for** where community members seek care for malaria the most and reasons |
| 5 | b) What do you think usually influence community members to seek malaria in a hospital? | **Probe** for the following   - Money for paying bills for treatment - Money for transportation to hospital - Distance of hospital - Access to hospital - Availability of hospital - Attitude of health workers - Preference for traditional medicine - Preference for a particular type of hospital (Private hospital, primary health centre, secondary health care facility) - Cultural norms and beliefs about seeking care in hospital - Cultural norms and beliefs malaria - Perceived seriousness of malaria - Perceived threat of malaria - Frequency of malaria episode |
| 6 | c) Let us briefly discuss how health seeking behaviour and source of care differ by socioeconomic group | **Probe** for –where each of the following categories of people usually seek malaria care?   - Rich people in the community - Poor people in the community - People with formal education - People without formal education   (**For each of the categories, probe for the common source of malaria treatment e.g** - government institutions, private clinics, pharmacy, chemists, drug peddlers/hawkers, traditional healing homes, herbal drug stores/kiosks) |
|  | **Malaria medications being used and their local names** | |
| 7 | a) What can you say about the malaria medications that community members use? | - Please tell us about malaria medications that are commonly used among adult population in the community. - **Probe for**, the local names for the malaria medications being used by adults in the community - Tell us about the malaria medications that are commonly used among children in the community. - **Probe for**, the local names for the malaria medications being used by children in the community - What are your views about the cost of the malaria medications (orthodox medicine)? - How affordable do you think the cost of malaria medications is for community members? |
|  | a) Please think about it If you were sick, which malaria medications would you take | **Probe for**,   - The local names for the malaria medications - The malaria medication(s) used the last time participant had malaria infection |
|  | b) What can you say about the use of herbal drugs for treating malaria by community members? | - Types of herbal drugs used for treating malaria - **Probe for**, the local names for the herbal malaria drugs - Perceived efficacy of the herbal drugs used for treating malaria - How common the use of herbal malaria drugs is in the community - Characteristics of community women who prefer to use herbal drugs for malaria treatment - Reasons community members prefer to use herbal drugs for malaria treatment - What are your views about the cost of the herbal malaria drugs? - Under what condition should community members who use herbal malaria drugs seek healthcare in the hospital for malaria treatment? |
|  | **Showing of pictures or physical packaging of malaria medications to improve recall and answers about usage of malaria medication** | |
| 8 | a)What are your opinions about the showing of pictures or physical packaging of malaria medications with the intention of improving recall and answers about usage of malaria medication? | **Probe** for   - Why showing of pictures or physical packaging of malaria medications can possibly improve recall and answers about usage of malaria medication? |
|  | b) In your opinion what other ways or means do you think can help community members to improve recall and answer about usage of malaria medications? |  |
|  | **Factors influencing use of malaria medications and treating malaria at a hospital** | |
| 9 | a) Please share with us the things that you think do influence community members decisions about choice of malaria medications | **Probe** for   - **Individual factors that influence the decisions about what malaria medicine to use – e.g.,** Age, sex (male or female), marital status, economic status, being pregnant, medical history, perceived seriousness of malaria, perceived threat of malaria, preference for particular type of malaria medications (orthodox drugs), preference for herbal malaria drugs - **Drug related factors that influence the decisions about what malaria medicine to use –** e.g. Availability of drug in hospital or drug stores, number of days expected to use drugs, number of capsules/dosages, taste of drug, side effects of drugs, drug resistance, availability of herbal malaria drugs, preference for herbal malaria drugs, proliferation of fake malaria medicines, giving of genuine malaria medications - **Social-cultural factors that influence the decisions about what malaria medicine to use –** e.g. support from partners, support family members, support from friends/peers, media influence and drug adverts, cultural norms, and values - **Health system related factors that influence the decisions about what malaria medicine to use-** e.g., availability of drugs in hospital/pharmacy, prescription of drug, advice from health workers - **Policy related factors e.g.** –governments and regulatory bodies recommendation relating to antimalarial drugs that should be first line of treatment - **Disease pattern related factors e.g.** –uncomplicated vs severe malaria - **Economic related factors** e.g. – cost of buying malaria medications, affordability of preferred malaria medications |
|  | b) What is your view about the cost of treating malaria in the hospital? | - How affordable do you think the cost of treating malaria is for community members in the health facility? |
|  | **Common methods that community members use to protect themselves from malaria** | |
| 10 | a) Please tell us about the common methods that people use to protect themselves from malaria in this community | **Probe** for (if not mentioned):   - Use of insecticide sprays - Use of insecticides treated bed nets - Use of mosquito replants - Use of coils - Use of window and door screens - Wearing of long-sleeved clothing and long pants - Malaria prophylaxis |
|  | b)Which method is most commonly used by people to protect themselves from malaria? | **Probe** for   - Reasons for being the most commonly used |
|  | **c)** Which methods do community members usually use to protect under-five children from malaria? |  |
|  | **Participation in community-based malaria programme** | |
| 11 | a) We are planning to put in place a free community-based programme where community members will be asked to consistently report malaria cases or symptoms. Our goal is to understand the transmission of malaria in the community to inform where interventions go. What is your opinion about how community members will perceive the programme. What is your opinion about how community members will perceive the programme? |  |
|  | b) If we want community members to be reporting malaria cases or symptoms consistently what do you think would be the preferred way or means for this? | **Probe** for (if not mentioned):   - Face-to-face with a project volunteer - Through community representative - Through SMS - Through WhatsApp - Through mobile application - Through hotline   Through community meeting etc. |
|  | c) Supposed we ask community members to be reporting malaria cases or symptoms via text message as part of our community-based malaria programme, what is your opinion about it? | **Probe** for:   - How easy do you think it will be for community members to participate? - How willing do you think community members will be? - What barriers do you think may be associated with it? - What do you think we can encourage community members to be reporting consistently malaria cases or symptoms via text message? |
|  | d) What is your opinion about asking community members to be reporting malaria cases or symptoms via a mobile application as part of our community-based malaria programme? | **Probe** for:   - How easy do you think it will be for community members to participate? - How willing do you think community members will be? - What barriers do you think may be associated with it? - What do you think we can encourage community members to be reporting consistently malaria cases or symptoms via a mobile application? |
|  | e) What can facilitate the participation of community members in the programme? |  |
|  | **Conclusion and other relevant information** | |
| 12 | a) Please tell us about on-going or existing community-based malaria interventions in this community |  |
|  | b) What suggestions do you have about how malaria can be controlled in this community? |  |
|  | c) What other suggestions do you have that can help us with the community-based malaria programme that are planning? |  |

**Socio-demographic information**

Ward and LGA ………………………………	Sex: ……………………

Age in years (at last birthday) …………	Highest level of Education………………

Primary occupation……………………	Other occupation(s)……………………..

Position…………………………………

Type of settlement ……………………………….…………..…………….

Name of Community/Area…………………………………….…………...

#### Cognitive testing consent form

*Who to interview:*

1. Mothers of under-five children

*Introduction*

My name is ……………and my colleagues are…………… We work with the University of Ibadan. We would like your opinion on various issues to enable us understand malaria transmission in urban areas as part of a collaborative project with Northwestern University, USA and Nigeria National Malaria Elimination Programme. The information we collect will help the government to plan health services to prevent malaria infections by ensuring you receive suitable interventions. You have been specially invited for this cognitive pretesting and we thank you for honouring our invitation.

*Purpose*

The objective of this cognitive pretest is to obtain information about a wide range of questionnaire problems. The cognitive pretest will afford us the opportunity of assessing the comprehensibility of questions, identifying problems that respondents have answering the questionnaire, establishing the causes of these problems, and generating suggestions for improvement based on these findings.

The information learned in this cognitive pretesting will be used to guide the development of other components of the study that we are planning to conduct.

**Procedure**

During this interview which will last for 30 – 45 minutes, we will be reading out to you a few questions that researchers commonly ask parents during community surveys. Some of these questions may sound familiar to you. We would like you to think about how easy or difficult it would be for you to answer each question and provide suggestions for how it could be better asked. For each question, we would like you to imagine that you are being asked to answer this about one of your children. We encourage you to feel free to ask any question or ask for clarification during the interview. During this interview, your views will be respected and will not be used against you in any way. This interview will be taped, so please speak up and speak clearly. We employ you to allow us to record the interview so that we will not miss out anything from the information you will be providing to us through the interview. Please do share your views without mentioning people’s names. We want the interview to be anonymous and to be confidential as possible. There is no right or wrong view, so feel free to express yourself. Please note that your participation in this interview is voluntary. Your decision not to be involved or drop at any point will not attract any penalty.

**Benefits**

Your participation in this study may not provide any personal benefit to you. However, should you decide to participate in this study, you will be doing society a great service because the findings of this study will be useful in the design of interventions and programmes for the control and prevention of malaria.

**Risks**

There are no known or anticipated risks associated with participation in this study beyond those experienced during an average conversation. If a question, or the discussion, makes you uncomfortable, you can choose not to answer.

**Confidentiality**

The information you share will be kept confidential. Identifying information will be removed from the transcripts. The transcripts and other electronic data will be retained for a maximum of 5 years, after which they will be destroyed. Data will be stored in an encrypted folder on protected laptop. Only the research team will have access to study data. No identifying information will be used in any presentations or publications based on this research.

**Contact**

If you have any questions or concerns regarding this study, please contact:

Professor IkeOluwapo Ajayi

Director, Institute for Advanced Medical Research and Training (IMARAT),

College of Medicine, University of Ibadan

Thank you for choosing to participate in the study. Kindly show by using any of the following 2 boxes, that your participation in this study was voluntary.

I will participate	I will not participate

#### Cognitive testing discussion guide

| S/N | **Main questions** | **Follow up questions/Probes** |
| --- | --- | --- |
|  | **Objective: Identify the challenges that survey participants encounter when responding to questions on malaria-related illness and treatment seeking from the Demographic and Health Survey and ascertain mitigating factors** | |
| 1 | a) Has your child been ill with a fever at any time in the last two weeks? | - Please tell me if you would be able to recall this accurately - Reasons for being able to recall accurately - Reasons for not being able to recall accurately - How comfortable are you answering this question? - How confident are you to easily recall your child’s malaria fever experience in the last two weeks? - How clear is this question to you? - Could you let me know if there is anything confusing about this question? - In case you feel this question is confusing, how will you suggest that we ask this question? - I will appreciate if you can tell me (by putting it in our own words) if asking whether your child had fever would capture a malaria episode. If not, what do you think we should inquire about to capture a malaria episode. |
| 2 | Did you seek advice or treatment for the illness from any source? | - Please tell me if you would be able to recall this accurately - Reasons for being able to recall accurately - Reasons for not being able to recall accurately - How comfortable are you answering this question? - How clear is this question to you? - Could you let me know if there is anything confusing about this question? - In case you feel this question is confusing, how will you suggest that we ask this question? |
| 3 | Where did you seek advice or treatment? Public sector government hospital? Private medical center? Chemist? Other? [response not required] | - Please tell me if you would be able to recall this accurately - Reasons for being able to recall accurately - Reasons for not being able to recall accurately - Do you understand the difference between public/government hospitals and private ones? - How comfortable are you answering this question? - How clear is this question to you? - Could you let me know if there is anything confusing about this question? - In case you feel this question is confusing, how will you suggest that we ask this question? |
| 4 | At the Pharmacy/Chemist/Patent Medicine Stores (PMS):  Was your child examined?  Did you get advice on the type of medication to buy, or did you already know which one to buy?  What is the name of the medication recommended or purchased?  (Probe separately for each of the three questions here) | - Please tell me if you would be able to recall this accurately - Reasons for being able to recall accurately - Reasons for not being able to recall accurately - How comfortable are you answering this question? - How clear is this question to you? - Could you let me know if there is anything confusing about this question? - In case you feel this question is confusing, how will you suggest that we ask this question? |
| 5 | At any time during the illness, did your child take any drugs for the illness?  What drugs did your child take? Artemisinin Combination Therapy? SP/Fansidar? Chloroquine? Amodiaquine? Quinine Pills? Injection/IV? Artesunate Rectal? Other Antimalarial (specify) | - Please tell me if you would be able to recall this accurately - Reasons for being able to recall accurately - Reasons for not being able to recall accurately - How comfortable are you answering this question? - How clear is this question to you? - Tell me if you are familiar with all the drugs that listed as options - Tell me the drugs you are unfamiliar to you - Tell me you if you familiar with what ACTS are - Could you let me know if there is anything confusing about this question? - In case you feel this question is confusing, how will you suggest that we ask this question? - How would you answer this question if you tested negative for malaria? |
| 6 | Here are a few photos of common medications that are commonly prescribed for malaria. Which one did your child take when they were ill? | - Please tell me if you would be able to recall this accurately - Reasons for being able to recall accurately - Reasons for not being able to recall accurately - How comfortable are you answering this question? - How clear is this question to you? - Between this question (question 6) and the previous one on medication use (question 5) which one is clearer to you? - Between this question (question 6) and the previous one medication (question 5), which one is easier to remember, and which question do you prefer? - What other ways can we ask this question to make it easy for you to remember which medication your child took? - Please think about it If you were sick, which medication would you take? |
| 7 | How long after the fever started did your child first take an artemisinin combination therapy? Same day? Next day? Two days after fever? Three or more days after fever? Don’t know? | - Please tell me if you would be able to recall this accurately - Reasons for being able to recall accurately - Reasons for not being able to recall accurately - How comfortable are you answering this question? - How clear is this question to you? - Could you let me know if there is anything confusing about this question? - In case you feel this question is confusing, how will you suggest that we ask this question? |
|  | **Objective: Assess comfort with answering questions about financial status** | |
| 8 | What is your total household income? (per month) | - Please tell me if you would be able to recall this accurately - Reasons for being able to recall accurately - Reasons for not being able to recall accurately - How clear is this question to you? - How comfortable are you answering this question? - Share your opinion whether people in your area will feel comfortable answering this question? - Who would be the most appropriate person to ask this question? - Could you let me know if there is anything confusing about this question? - In case you feel this question is confusing, how will you suggest that we ask this question |
| 9 | How many members of the household earn income? | - Please tell me if you would be able to recall this accurately - Reasons for being able to recall accurately - Reasons for not being able to recall accurately - How clear is this question to you? - How comfortable are you answering this question? - Share your opinion whether people in your area will feel comfortable answering this question? - Who would be the most appropriate person to ask this question? - Could you let me know if there is anything confusing about this question? - In case you feel this question is confusing, how will you suggest that we ask this question |
| 10 | How do members of your household earn income? [salaried/hourly wage or intermittent work] | - Please tell me if you would be able to recall this accurately - Reasons for being able to recall accurately - Reasons for not being able to recall accurately - How clear is this question to you? - How comfortable are you answering this question? - Share your opinion whether people in your area will feel comfortable answering this question? - Who would be the most appropriate person to ask this question? - Could you let me know if there is anything confusing about this question? - In case you feel this question is confusing, how will you suggest that we ask this question |
| 11 | How much do you spend on personal expenses, such as food and transportation? On a weekly basis? On a monthly basis? | - Please tell me if you would be able to recall this accurately - Reasons for being able to recall accurately - Reasons for not being able to recall accurately - How clear is this question to you? - How comfortable are you answering this question? - Share your opinion whether people in your area will feel comfortable answering this question? - Who would be the most appropriate person to ask this question? - Could you let me know if there is anything confusing about this question? - In case you feel this question is confusing, how will you suggest that we ask this question - In terms of personal expenses such as food and transportation, will it easier to recall expenses in terms of weekly expenses or monthly expenses? |
| 12 | What is your primary mode of transportation? | - How clear is this question to you? - How comfortable are you answering this question? - Share your opinion whether people in your area will feel comfortable answering this question? - Who would be the most appropriate person to ask this question? - Could you let me know if there is anything confusing about this question? - In case you feel this question is confusing, how will you suggest that we ask this question |
| 13 | Are your children enrolled in school? | - How clear is this question to you? - How comfortable are you answering this question? - Share your opinion whether people in your area will feel comfortable answering this question? - Who would be the most appropriate person to ask this question? - Could you let me know if there is anything confusing about this question? - In case you feel this question is confusing, how will you suggest that we ask this question |
| 14 | How much do you spend per term on children’s education? | - Please tell me if you would be able to recall this accurately - Reasons for being able to recall accurately - Reasons for not being able to recall accurately - How clear is this question to you? - How comfortable are you answering this question? - Share your opinion whether people in your area will feel comfortable answering this question? - Who would be the most appropriate person to ask this question? - Could you let me know if there is anything confusing about this question? - In case you feel this question is confusing, how will you suggest that we ask this question |
| 15 | Do you receive money from family abroad? How much do you receive on a monthly basis? | - Please tell me if you would be able to recall this accurately - Reasons for being able to recall accurately - Reasons for not being able to recall accurately - How clear is this question to you? - How comfortable are you answering this question? - Share your opinion whether people in your area will feel comfortable answering this question? - Who would be the most appropriate person to ask this question? - Could you let me know if there is anything confusing about this question? - In case you feel this question is confusing, how will you suggest that we ask this question |
| 16 | Do you have financial support from other sources? | - Please tell me if you would be able to recall this accurately - Reasons for being able to recall accurately - Reasons for not being able to recall accurately - How clear is this question to you? - How comfortable are you answering this question? - Share your opinion whether people in your area will feel comfortable answering this question? - Who would be the most appropriate person to ask this question? - Could you let me know if there is anything confusing about this question? - In case you feel this question is confusing, how will you suggest that we ask this question |
| 17 | If you or a member of your household was sick and needed to go the hospital, would you: a) have enough funds to cover all costs, b) have enough funds for some of the costs, c) need to borrow money or find other means to pay | - Please tell me if you would be able to recall this accurately - Reasons for being able to recall accurately - Reasons for not being able to recall accurately - How clear is this question to you? - How comfortable are you answering this question? - Share your opinion whether people in your area will feel comfortable answering this question? - Who would be the most appropriate person to ask this question? - Could you let me know if there is anything confusing about this question? - In case you feel this question is confusing, how will you suggest that we ask this question |
| 18 | If a member of this household were sick and needed to go to the hospital, would you have enough money for/access to reliable transportation? | - Please tell me if you would be able to recall this accurately - Reasons for being able to recall accurately - Reasons for not being able to recall accurately - How clear is this question to you? - How comfortable are you answering this question? - Share your opinion whether people in your area will feel comfortable answering this question? - Who would be the most appropriate person to ask this question? - Could you let me know if there is anything confusing about this question? - In case you feel this question is confusing, how will you suggest that we ask this question |
| 19 | How do members of your household manage health expenses when there isn’t enough money? | - Please tell me if you would be able to recall this accurately - Reasons for being able to recall accurately - Reasons for not being able to recall accurately - How clear is this question to you? - How comfortable are you answering this question? - Share your opinion whether people in your area will feel comfortable answering this question? - Who would be the most appropriate person to ask this question? - Could you let me know if there is anything confusing about this question? - In case you feel this question is confusing, how will you suggest that we ask this question |

### Cross-sectional study instruments

#### Household survey consent form and questionnaire

| **BACKGROUND INFORMATION** | | | |
| --- | --- | --- | --- |
| LOCAL GOVT. AREA………………………………………………………………………… | | | |
| WARD……………………………………………………………………………………………………………....... | | | |
| ENUMERATION AREA/CLUSTER NUMBER………………....................................................................... | | | |
| HOUSEHOLD NUMBER …………………………………………………………………………………………. | | | |
| NAME OF HOUSEHOLD HEAD………………………………………………...…………………………........ | | | |
| DATE: DAY……… MONTH………YEAR………… | | | |
| INTERVIEWER'S NAME ……………………………………. | | | |
| INTERVIEWERS PHONE NO………………………………………… | | | |
| INTERVIEWER VISIT | | | |
|  | 1 | 2 | 3 |
| DATE | __________________ | ___________________ | ___________________ |
| RESULT |  |  |  |
| SUPERVISORS NAME | ___________________ | FIELD EDITOR | ___________________ |
| **INTRODUCTION AND CONSENT** My name is ……………and my colleagues are…………… I am working with the University of Ibadan. We would like your opinion on various issues to enable us understand malaria transmission in urban areas part of a collaborative project with Northwestern University, USA and National Malaria Elimination Programme. The information we collect will help the government to plan health services to prevent malaria infections by ensuring you receive suitable interventions. We thank you for honouring our presence. **Purpose** The purpose of this interview is to investigate malaria prevalence in your household and identify what factors predispose community members to malaria infection **Procedure** As part of this study, you will be asked questions about your household characteristics, individuals living within the household, socio-demographic information, knowledge of malaria, risk factors and other relevant questions, which will help us achieve our objectives. You will also be offered rapid diagnostic testing for malaria infection. The interview should last between 30- 45 minutes. Your responses will be inputted into an electronic data-capturing device. The interview is going to be anonymous and confidential as much as possible. You can choose whether to participate in the interview, and you may stop at any time during the study. Your decision not to continue to participate will not attract any penalty. **Benefits** Your participation in this study may not provide any personal benefit to you. However, should you decide to participate in this study, you will be doing society a great service because the findings of this study will be useful in the design of interventions and programmes for the control and prevention of malaria in your community. **Risks** There are no known or anticipated risks associated with participation in this study beyond those experienced during an average conversation. If a question makes you uncomfortable, you can choose not to answer. **Confidentiality** The information you share will be kept confidential. Identifying information will be removed from the electronic data. The electronic data will be retained for a maximum of 5 years, after which they will be destroyed. Data will be stored in an encrypted folder on protected laptop. Only the research team will have access to study data. No identifying information will be used in any presentations or publications based on this research. **Contact** If you have any questions or concerns regarding this study, please contact: Professor IkeOluwapo Ajayi, Tel: 08023268431 Director, Institute for Advanced Medical Research and Training (IMARAT), College of Medicine, University of Ibadan Thank you for choosing to participate in the study. Kindly show by using any of the following 2 boxes, that your participation in this study was voluntary. I will participate I will not participate 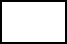Do you have any questions? Yes _______________1 No________________2 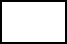May I begin the interview now? Yes _______________1 No________________2 | | | |

| **HOUSEHOLD LINE LISTING** | | | | | | | | | | | | |
| --- | --- | --- | --- | --- | --- | --- | --- | --- | --- | --- | --- | --- |
| Line No | Usual Residents | | Relationship to Household  Head (HH) | | Sex | | Residence | | Age | Marital Status (If Older than 18) | | RDT Eligibility |
|  | Please give me the names of the persons who usually live here | | What is the relationship of (NAME) to the HH | | Is (NAME) male or female?  Male = 1  Female = 2 | | Did (NAME) sleep here last night?  Yes =1  No = 2 | | How old was  (NAME) as at last birthday? | What is (NAME) current marital status | | Selected for RDT?  Yes =1  No = 2 |
| 1 |  | |  | |  | |  | |  |  | |  |
| 2 |  | |  | |  | |  | |  |  | |  |
| 3 |  | |  | |  | |  | |  |  | |  |
| 4 |  | |  | |  | |  | |  |  | |  |
| 5 |  | |  | |  | |  | |  |  | |  |
| 6 |  | |  | |  | |  | |  |  | |  |
| 7 |  | |  | |  | |  | |  |  | |  |
| 8 |  | |  | |  | |  | |  |  | |  |
| 9 |  | |  | |  | |  | |  |  | |  |
| 10 |  | |  | |  | |  | |  |  | |  |
| 11 |  | |  | |  | |  | |  |  | |  |
| 12 |  | |  | |  | |  | |  |  | |  |
| 13 |  | |  | |  | |  | |  |  | |  |
| 14 |  | |  | |  | |  | |  |  | |  |
| 15 |  | |  | |  | |  | |  |  | |  |
| **Information on non-residents** | | | | | | | | | | | | |
| Line No | Non-residents | Relationship to HH | | Sex | | Residence | | Age | | | Remarks | |
|  | Please give me the names of all persons who don’t usually live here | What is the relationship of (NAME) to the HH | | Is (NAME) male or female?  Male = 1  Female = 2 | | Did (NAME) sleep here last night?  Yes = 1  No = 2 | | How old was  (NAME) as at last birthday? | | |  | |
| 1 |  |  | |  | |  | |  | | |  | |
| 2 |  |  | |  | |  | |  | | |  | |
| 3 |  |  | |  | |  | |  | | |  | |
| 4 |  |  | |  | |  | |  | | |  | |
| 5 |  |  | |  | |  | |  | | |  | |

| CODES: |  |
| --- | --- |
| **RELATIONSHIP TO HEAD OF HOUSEHOLD** |  |
| 01 = HEAD | 09 = BROTHER-IN-LAW/SISTER IN-LAW |
| 02 = WIFE OR HUSBAND | 10 = NIECE/NEPHEW BY BLOOD |
| 03 = SON OR DAUGHTER | 11 = NIECE/NEPHEW BY MARRIAGE |
| 04 = SON-IN-LAW OR DAUGHTER-IN-LAW | 12 = OTHER RELATIVE |
| 05 = GRANDCHILD | 13 = ADOPTED/FOSTER/ STEPCHILD |
| 06 = PARENT | 14 = NOT RELATED |
| 07 = PARENT-IN-LAW | 15 = CO-WIFE |
| 08 = BROTHER OR SISTER | 16 – NANNY |
|  | 98 = DON'T KNOW |

**CRITERIA FOR RDT**

(if less than or equal to 5 people in the household, test everyone, if greater than 5 people, test one person in these age groups:

0-5 years

6-10 years

11-17 years

18-30 years

30 years and above

*If age category is missing, test two persons in the youngest category*

| **SECTION 1. HOUSEHOLD RESOURCES** | | | | |
| --- | --- | --- | --- | --- |
| Q100 | What is the main source of drinking water for members of your household?  (enter the number for the most commonly used) | **Improved source**   i. Piped into dwelling/yard/plot --------------------------1 ii. Piped to neighbour ----------------------------------------2 iii. Public tap/standpipe --------------------------------------3 iv. Tube well or borehole ------------------------------------4 v. Protected dug well ----------------------------------------5 vi. Protected spring -------------------------------------------6 vii. Rainwater ---------------------------------------------------7 viii. Tanker truck/cart with small tank ---------------------8 ix. Bottled water -----------------------------------------------9   **Unimproved source**   x. Unprotected dug well ----------------------------------10 xi. Unprotected spring -------------------------------------11 xii. Surface water (River, Pond) --------------------------12 xiii. Sachet water----------------------------------------------13 xiv. **Others (specify)**------------------------------------------14 | |  |
| Q101 | What is the main source of water used by your household for other purposes such as cooking and  handwashing? | **Improved source**   i. Piped into dwelling/yard/plot -------------------------1 ii. Piped to neighbour ---------------------------------------2 iii. Public tap/standpipe -------------------------------------3 iv. Tube well or borehole -----------------------------------4 v. Protected dug well ---------------------------------------5 vi. Protected spring ------------------------------------------6 vii. Rainwater ---------------------------------------------------7 viii. Tanker truck/cart with small tank --------------------8 ix. Bottled water ---------------------------------------------9   **Unimproved source**   x. Unprotected dug well ----------------------------------10 xi. Unprotected spring -------------------------------------11 xii. Surface water (River, Pond) ---------------------------12 xiii. Sachet water-----------------------------------------------13   **Others (specify)**---------------------------------------------------14 | |  |
| Q102 | Where is the main source of water located? | i. In own dwelling ---------------------------------------1 ii. Outside own dwelling -------------------------------2 iii. Public tap-----------------------------------------------3 iv. Elsewhere (specify) ----------------------------------4 | |  |
| Q103 | On the average, how long does it take your household to get to the source of water and back? | ----------- Minutes  ------------Hours  ------------Don’t Know | |  |
| Q104 | What kind of toilet facilities do members of your family usually use? | **Improved sanitation facility**   i. Flush toilet --------------------------------------------------1 ii. Ventilated improved pit (VIP) latrine -----------------2 iii. Pit latrine with slab ---------------------------------------3 iv. Composting toilet -----------------------------------------4   **Unimproved facility**   v. Pit latrine without slab/open pit ----------------------5 vi. Bucket -------------------------------------------------------6 vii. Hanging toilet/hanging latrine ------------------------7 viii. Open defecation (no facility/bush/field) -----------8 ix. Others (specify) ------------------------------------------98 | |  |
| Q105 | Do you have your own toilet or you share toilet with other households? | i. Have own toilet--------------------------------------1 ii. Shared toilet -----------------------------------------2 | |  |
| Q106 | Where is the bathroom of your house located? | i. Inside the house ------------------------------------1 ii. Outside, separated from the house ------------2 iii. No bathroom at all----------------------------------3 | |  |
| Q107 | How many rooms are used by members of your household for sleeping? | Specify the number of rooms used: ….…………...... | |  |
| Q108 | How many members of your household sleep on the floor? | ***Give number, if none write 00***  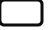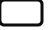 | |  |
| Q109 | What is the main source of power/energy use for cooking in your household? | i. Electricity ------------------------------------------------1 ii. LPG/natural gas/biogas -------------------------------2 iii. Kerosene -------------------------------------------------3 iv. Coal/lignite ----------------------------------------------4 v. Charcoal --------------------------------------------------5 vi. Wood -----------------------------------------------------6 vii. Agricultural crop/straw/shrubs/grass ------------7 viii. Others specify ------------------------------------------9 | |  |
| Q110 | Which of the following items do you, your spouse or your family have? (Circle as appropriate) |  | |  |
|  |  |  | Yes | No |
|  |  | Radio | 1 | 2 |
|  |  | Television | 1 | 2 |
|  |  | Mobile telephone | 1 | 2 |
|  |  | Non-mobile telephone | 1 | 2 |
|  |  | Desktop Computer | 1 | 2 |
|  |  | Laptop Computer |  |  |
|  |  | Refrigerator | 1 | 2 |
|  |  | Table | 1 | 2 |
|  |  | Chair | 1 | 2 |
|  |  | Bed | 1 | 2 |
|  |  | Cupboard | 1 | 2 |
|  |  | Air conditioner | 1 | 2 |
|  |  | Electric iron | 1 | 2 |
|  |  | Generator | 1 | 2 |
|  |  | Fan | 1 | 2 |
| Q111 | Do your household own farmland? | Yes--------------------------------------------------------------------1  No---------------------------------------------------------------------2 | |  |
| Q112 | Does any member of your household have livestock? | Yes -------------------------------------------------------------------1  No--------------------------------------------------------------------2 | |  |
| Q113 | Type of housing | Face to face………………………………..……….……………………1  One Bedroom…………………………………….………………….….2  Two Bedroom Flat…………………………………………………....3  Three Bedroom Flat…………………………………………….…...4  Duplex………………………………………….……………………….….5  Others(specify)………………………………………………………….6 | |  |
| Q114 | Do you share your compound with other households? | Yes……………………………………………..……………………………1  No…………………………………………………………….................2 | |  |
| Q115 | Are the eaves of the house or building occupied by this household open or closed?  **Observe and record** | Completely Open .......................................................1  Partially Open…..........................................................2  Closed.........................................................................3 | |  |
| Q116 | Does the part of the house or building occupied by the household have a ceiling?  **Observe and record** | No, None ....................................................................1  Yes, Partial/Poorly Sealed/Worn Out..........................2  Yes, Complete and Sealed …........................................3 | |  |
| Q117 | Is there a farm within the compound?  **Observe and record** | Yes...............................................................................1  No................................................................................2 | |  |

| **SECTION 2: NEIGHBOURHOOD CHARACTERISTICS** | | | | | | |
| --- | --- | --- | --- | --- | --- | --- |
| Q201 | How would you describe the road type in your neighbourhood | Tarred……………………………………………………………………...1  Untarred…………………………………………………………………..2  Both Tarred/untarred…………………………………………......3 | | | | |
| Q202 | Would you say most houses in your neighbourhood are? | Fenced with gate……………………………………………………..1  Fenced, no gate………………………………………………….……2  Partially Fenced……………………………………………………….3  Partially Fenced……………………………………………………….4  Other(specify)……………………………………………..………....5 | | | | |
| Q203 | If fenced, what is the nature of the fencing material | Cement Block…………………………………………..………..……1  Mud…………………………………………………………..…….…....2  Barbed wire and cement block………………………..……..3  Others(specify)……………………………………………………….4 | | | | |
| Q204 | Are the houses in your neighbourhood painted? | No not painted……………………………………………………...1  Yes, old painting…………………………………………………...2  Yes, recently painted…………………………….……………...3  Others (specify)…………………………………………………….4 | | | | |
| Q205 | Are there open drainages in your neighborhood | Yes............................................................................1  No……………………………………………………......................2 | | | | |
| Q206 | Are the open drainages clogged with dirt or rubbish | Yes............................................................................1  No……………………………………………………......................2 | | | | |
| Q207: Thinking about the environment where you live, how much do you agree with the following statements | | | | | | |
|  |  | Strongly Agree | Agree | Can’t Say | Disagree | Strongly Disagree |
| Q207A | There is a lot of noise in my neighbourhood. | 1 | 2 | 3 | 4 | 5 |
| Q207B | There are sidewalks on most streets in my community. | 1 | 2 | 3 | 4 | 5 |
| Q207C | There is a lot of unpleasant smells in my neighbourhood. | 1 | 2 | 3 | 4 | 5 |
| Q207D | My neighbourhood has heavy human traffic. | 1 | 2 | 3 | 4 | 5 |
| Q207E | There is a lot of trash and litter on the street in my neighbourhood. | 1 | 2 | 3 | 4 | 5 |
| Q207F | There is vandalism in my neighbourhood. | 1 | 2 | 3 | 4 | 5 |
| Q207G | There are too many people hanging around on the streets near my home | 1 | 2 | 3 | 4 | 5 |
| Q207H | I have easy access to medical care in my neighbourhood. | 1 | 2 | 3 | 4 | 5 |

| **SECTION 3: INDIVIDUAL CHARACTERISTICS** | | | | |
| --- | --- | --- | --- | --- |
| Q300 | How long have you been living continuously in (CURRENT PLACE OF RESIDENCE)? | DURATION IN YEARS | |  |
| Q301 | How old were you at your last birthday?  PROBE TO GET ESTIMATE IF NOT SURE | AGE AT LAST BIRTHDAY (IN YEARS)  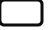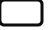 | | |
| Q302 | Sex of Respondent | Male…………………………………………………….1  Female………………………………………………….2 | | |
| Q303 | Have you ever attended school?  (Ask all questions) | Quranic/Islamiyah school?  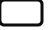1. YES  2. NO | Adult classes?  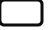1. YES  2. NO | Formal school?  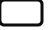1. YES  2. NO |
| Q304 | What is the highest level of formal school you completed? | i. Did not complete Primary School ----------------------1 ii. Primary Completed-----------------------------------------2 iii. Secondary Completed-------------------------------------3 iv. Post-secondary School Completed---------------------4 | | |
| Q305 | If post-secondary school completed: Specify highest level completed in this category: | If completed post-secondary education, please, specify highest level completed:  ______________________________________________ | | |
| Q306 | What is your ethnic group? | i. Igbo ----------------------------------------------------1 ii. Yoruba -------------------------------------------------2 iii. Hausa --------------------------------------------------3 iv. Others (specify)--------------------------------------4 | | |
| Q307 | What is your religion? | i. Christianity -------------------------------------------1 ii. Islam ---------------------------------------------------2 iii. Traditional religion ---------------------------------3 iv. Other (specify)---------------------------------------4 | | |
| Q308 | What is your current marital status? | i. Never married ------------------------------------------1 ii. Married ---------------------------------------------------2 iii. Co-habiting ----------------------------------------------3 iv. Divorced/Separated -----------------------------------4 v. Widowed -----------------------------------------------5 | | |
| Q309 | How would you describe the family living arrangement in your house? | i. Monogamous Family (a man and wife, with or without children) living in a separate house ----------------1 ii. Polygamous Family (a man, wives, and children) living in a separate house -----------------------------2 iii. Extended family (a family which extends across generations, i.e including grandparents, aunts, and other relatives) living in the same house ……………………………………………………....3 iv. A female-headed household (a woman and her children i.e. absence of a man in a household, thus, making the woman the sole economic provider for the family) ................................................................4   Others (specify) ----------------------------------------------------5 | | |
| Q310 | Who provides the main source of income in your home? | i. Husband/Partner …………………………………………………..1 ii. Both Husband and Wife(s) provide equally…………….2 iii. Wife……………………………………………............................3 iv. Parents ………………………………………………………….……..4 v. Children…………………………………………………………….....5   Others (specify) …………………………………….….............................6 | | |
| Q311 | Do you currently do any work to earn an income? | Yes…………………………………………………….......1  No……………………………………………………….....2 | | |
| Q312 | What kind of work do you do? | i. Professional/Technical/Managerial-------------------------1 ii. Clerical ---------------------------------------------------------2 iii. Sales and Services ----------------------------------------------3 iv. Skilled Manual ---------------------------------------------------4 v. Unskilled Manual -----------------------------------------------5 vi. Agriculture --------------------------------------------------6 vii. Domestic Work---------------------------------------------7 viii. Others (specify):-------------------------------------------8 | | |
| Q313 | If you are an agricultural worker, what type of agricultural produce do you work with? | i. Rice .......................................................................1  ii. Other plants...........................................................2  iii. Poultry..................................................................3  iv. Fish and other seafood........................................4  v. Cows....................................................................5  vi. Other livestock....................................................6  vii. Other produce....................................................7 | | |
| Q314 | If you are an agricultural worker, is your work seasonal? | Yes...............................................................................1  No.................................................................................2 | | |
| Q315 | If your work is seasonal, what months of the year do you work? | ................................................................................... | | |
| Q317 | Where do you do your work? | State..........................................................................  LGA...........................................................................  Ward.......................................................................... | | |
| Q318 | Do you work indoors, outdoors or both? | Indoors......................................................................  Outdoors...................................................................  Both.......................................................................... | | |
| Q319 | How long do you spend indoor or outdoor at work?  **Ask for times and write response in hours** | At work (Overall)......................................................  Indoor at Work.........................................................  Outside at Work....................................................... | | |
| Q320 | How much do you earn? | Daily........................................................................  Weekly....................................................................  Monthly................................................................... | | |
| Q321 | Do you have children enrolled in school | Yes……………………………………………………….1  No………………………………………………………..2 | | |
| Q322 | How many children are enrolled? | /________/ | | |
| Q323 | Do your children attend school in the same neighbourhood that you reside in? | Yes………….............................................................1  No……………………………………………………….2 | | |
|  | If, your children school outside this neighbourhood, can you provide information about? | LGA............................................................................  Ward........................................................................ | | |

|  | **IF RESPONDENT IS MALE, SKIP SECTION 4 AND GO TO SECTION 5** | | | |
| --- | --- | --- | --- | --- |
| **SECTION 4 : REPRODUCTION** | | | | |
| Q400 | Have you ever given birth | Yes…………………………..………………………….1  No………………………………………………….……2 | | |
| Q401 | Do you have any sons or daughters to whom you have given birth to who are now living with you | Yes…………………………………………...…………1  No…………………………………………………..…...2 | | |
| Q402 | How many sons live with you? | /_________________/ | | |
| Q403 | How many daughters live with you? | /________________/ | | |
|  | Do you have any sons or daughters to whom you have given birth who are alive but do not live with you | Yes………………………………………………………1  No…………………………………………..…………...2 | | |
| Q404 | How many sons are alive but do not live with you | /_____________________/ | | |
| Q405 | How many daughters are alive but do not live with you | /____________________/ | | |
| Q406 | Have you ever given birth to a boy or girl who was born alive and later died? | Yes……………………………………….…………….1  No………………………………………………………2 | | |
| Q407 | How many boys have died? | /___________/ | | |
| Q408 | How many girls have died? | /__________/ | | |
| Q408b | Just to make sure that I am correct you have had a Total of -------births in your lifetime | /__________/ | | |
| Q409 | Have you ever heard of anything that can be done to prevent pregnancy? | Yes………………………………………..…………….1  No……………………………………….………………2 | | |
| Q410 | If yes, what was your main source of information |  | | |
| Q411 | Have you ever done anything to prevent pregnancy? | Yes……………………………………………………….1  No……………………………………………………...…2 | | |
| Q412 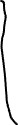 | If yes, which of these methods have you done? |  | Yes | No |
|  |  | Female sterilization | 1 | 2 |
|  |  | Male sterilization | 1 | 2 |
|  |  | Pill | 1 | 2 |
|  |  | IUD | 1 | 2 |
|  |  | Injectables | 1 | 2 |
|  |  | Implants | 1 | 2 |
|  |  | Male condom | 1 | 2 |
|  |  | Female condom | 1 | 2 |
|  |  | Emergency contraception | 1 | 2 |
|  |  | Standard days method | 1 | 2 |
|  |  | Lactational amenorrhea (LAM) | 1 | 2 |
|  |  | Rhythm | 1 | 2 |
|  |  | Withdrawal | 1 | 2 |
|  |  | Other method | 1 | 2 |
| Q413 | Are you currently pregnant? | Yes……………………………………………………...1  No…………………………………………………….…2 | | |

| What name was given to your most recent/previous baby | Is (NAME) a boy or girl? | Was it a single of multiple pregnancy | On what day, month and year was(NAME) born | Is (NAME) still alive | How old was (NAME) as at last birthday? | Is  (NAME) living with you | Was (NAME)  Selected |
| --- | --- | --- | --- | --- | --- | --- | --- |
|  | Boy …1  Girl… 2 | Sing…..1  Mult……2 | Day/__/  Month /__/  Year/_____/ | Yes…1  No….2 | /____/  years | Yes…1  No…..2 | Yes…1  No…..2 |
|  | Boy …1  Girl… 2 | Sing…..1  Mult……2 | Day/__/  Month /__/  Year/_____/ | Yes…1  No….2 | /____/  years | Yes…1  No…..2 | Yes…1  No…..2 |
|  | Boy …1  Girl… 2 | Sing…..1  Mult……2 | Day/__/  Month /__/  Year/_____/ | Yes…1  No….2 | /____/  years | Yes…1  No…..2 | Yes…1  No…..2 |
|  | Boy …1  Girl… 2 | Sing…..1  Mult……2 | Day/__/  Month /__/  Year/_____/ | Yes…1  No….2 | /____/  years | Yes…1  No…..2 | Yes…1  No…..2 |
|  | Boy …1  Girl… 2 | Sing…..1  Mult……2 | Day/__/  Month /__/  Year/_____/ | Yes…1  No….2 | /____/  years | Yes…1  No…..2 | Yes…1  No…..2 |
|  | Boy …1  Girl… 2 | Sing…..1  Mult……2 | Day/__/  Month /__/  Year/_____/ | Yes…1  No….2 | /____/  years | Yes…1  No…..2 | Yes…1  No…..2 |
|  | Boy …1  Girl… 2 | Sing…..1  Mult……2 | Day/__/  Month /__/  Year/_____/ | Yes…1  No….2 | /____/  years | Yes…1  No…..2 | Yes…1  No…..2 |
|  | Boy …1  Girl… 2 | Sing…..1  Mult……2 | Day/__/  Month /__/  Year/_____/ | Yes…1  No….2 | /____/  years | Yes…1  No…..2 | Yes…1  No…..2 |
|  | Boy …1  Girl… 2 | Sing…..1  Mult……2 | Day/__/  Month /__/  Year/_____/ | Yes…1  No….2 | /____/  years | Yes…1  No…..2 | Yes…1  No…..2 |
|  | Boy …1  Girl… 2 | Sing…..1  Mult……2 | Day/__/  Month /__/  Year/_____/ | Yes…1  No….2 | /____/  years | Yes…1  No…..2 | Yes…1  No…..2 |
|  | Boy …1  Girl… 2 | Sing…..1  Mult……2 | Day/__/  Month /__/  Year/_____/ | Yes…1  No….2 | /____/  years | Yes…1  No…..2 | Yes…1  No…..2 |
|  | Boy …1  Girl… 2 | Sing…..1  Mult……2 | Day/__/  Month /__/  Year/_____/ | Yes…1  No….2 | /____/  years | Yes…1  No…..2 | Yes…1  No…..2 |
|  | Boy …1  Girl… 2 | Sing…..1  Mult……2 | Day/__/  Month /__/  Year/_____/ | Yes…1  No….2 | /____/  years | Yes…1  No…..2 | Yes…1  No…..2 |
|  | Boy …1  Girl… 2 | Sing…..1  Mult……2 | Day/__/  Month /__/  Year/_____/ | Yes…1  No….2 | /____/  years | Yes…1  No…..2 | Yes…1  No…..2 |

| Now I would like to ask you about your last pregnancy that resulted in a life birth | | |
| --- | --- | --- |
| Q415 | While you were pregnant for (NAME) did you see anyone for antenatal care? | Yes……………………………………………………………..1  No…………………………..…………………….…………...2 |
| Q416 | Whom did you see? | Doctor……………………………………………………..…1  Nurse/Midwife…………………………………………..2  Auxiliary Midwife…………………………………..…..3  Traditional Birth Attendant…………………..…...4  Community Health Worker………………….…….5  Others(specify)_______________________6 |
| Q417 | Where did you receive antenatal care? | Government Hospital…………………………………1  Government Health Center………………………..2  Government Health Post……………………………3  Other Public Sector(specify).......………………..4  Private Hospital………………………………………….5  Private Clinic………………………………………………6  Other Private Sector(specify)……………………..7  NGO Hospital……………………………………………..8  NGO Clinic………………………………………………….9  Other NGO Center(specify)…………………………10  Home………………………………………………………….11  Traditional Birth Homes………………………………12  Others(specify)________________________13 |
| Q418 | How many weeks or months were you when you first received antenatal care? | Weeks_____________________  Months_____________________ |
| Q419 | How many times did you receive antenatal care during the last pregnancy? |  |
| Q420 | During the last pregnancy, did you take SP/Fansidar to prevent malaria | Yes………………………………………………….……….1  No……………………………………………………………2 |
| Q421 | How many times did you take SP/Fansidar | /___________________/ |
| Q422 | Where did you get the SP/Fansidar | During Antenatal Visit……………………………………..1  Another Facility Visit………………………………………..2  Other Source(Specify).........................................3 |

| **SECTION 5: KNOWLEDGE OF MALARIA TRANSMISSION, CAUSES AND PREVENTIVE PRACTICES** | | | | |
| --- | --- | --- | --- | --- |
| Q500 | Have you ever heard of malaria | Yes………………………………………………………1  No……………………………………………………….2 | | |
| Q501 |  |  | Yes | No |
|  | What was your source of information on malaria | Radio | 1 | 2 |
|  |  | Television | 1 | 2 |
|  |  | Newspaper | 1 | 2 |
|  |  | Friend | 1 | 2 |
|  |  | Health Care Worker | 1 | 2 |
|  |  | Religious leader | 1 | 2 |
|  |  | Colleague | 1 | 2 |
|  |  | Others (Specify)____________ |  | |
| Q502 | What are the common symptoms of malaria?  **(DO NOT READ OUT)** |  | Mentioned | Not Mentioned |
|  |  | Fever | 1 | 2 |
|  |  | Chills/Shivering | 1 | 2 |
|  |  | Headache | 1 | 2 |
|  |  | Joint Pain | 1 | 2 |
|  |  | Poor Appetite | 1 | 2 |
|  |  | Vomiting | 1 | 2 |
|  |  | Convulsion | 1 | 2 |
|  |  | Cough | 1 | 2 |
|  |  | Catarrh/Nasal Congestion | 1 | 2 |
|  |  | Don’t know any | 3 |  |
|  |  | Others(Specify) |  | |
| Q503 | What are the causes of malaria you know?  **(DO NOT READ OUT)** |  |  | |
|  |  | Mosquitoes bite | 1 | 2 |
|  |  | Dirty Environment | 1 | 2 |
|  |  | Stagnant Water | 1 | 2 |
|  |  | Lakes, pits, dams around surroundings | 1 | 2 |
|  |  | Bushes around the house | 1 | 2 |
|  |  | Ill ventilated houses | 1 | 2 |
| Q504 | Do you think malaria can be prevented? | Yes………………………………………………………1  No……………………………………………………….2 | | |
| Q505 | Which was do you think malaria can be prevented? |  | Yes | No |
|  |  | Sleeping under insecticide treated nets | 1 | 2 |
|  |  | Cutting bush around the house | 1 | 2 |
|  |  | Spraying mosquito insecticide in homes | 1 | 2 |
|  |  | Taking malaria preventive drug | 1 | 2 |
|  |  | Door/Window Screens | 1 | 2 |
|  |  | Taking herbs | 1 | 2 |
|  |  | By praying | 1 | 2 |

| **SECTION 6: HISTORY OF MALARIA INFECTION; HEALTH SEEKING BEHAVIOUR FOR MOST RECENT EPISODE OF MALARIA AND ANTIMALARIAL DRUG USE PATTERN** | | | |
| --- | --- | --- | --- |
| Q600 | Has anyone been ill with a fever at any time in the last 2 weeks? | | Yes………………………..1  No…………………………2 |
| Q601 | Has anyone been ill with malaria at any time in the last 2 weeks? | | Yes………………………..1  No…………………………2 |
| Q602 | How many people in your household had fever in the last 2 weeks? | |  |
| Q603 | Can you list all people that have had a fever in the last 2 weeks? | | NAME______________________  LINE NO___________________  NAME______________________  LINE NO____________________  NAME______________________  LINE NO____________________  NAME_____________________  LINE NO____________________ |
| Q604 | Did you seek advice or treatment for the illness for (NAME) from any source? | | Yes…………………………..…..1  No………………………..………2 |
| Q605 | Where did you first seek advice or treatment? | **PUBLIC SECTOR**  Government hospital ……………………………………………………. 1  Government health center.................................................2  Government health post....................................................3  Mobile clinic.......................................................................4  Fieldworker/CHW...............................................................5  Other public sector (specify)..............................................6  **PRIVATE MEDICAL SECTOR**  Private hospital/clinic........................................................7  Pharmacy.........................................................................8  Chemist/PMV...................................................................9  Private doctor..................................................................10  Mobile clinic ....................................................................11  Other private medical sector (specify) ……………..............12  **OTHER SOURCE**  Shop..................................................................................13  Traditional practitioner.....................................................14  Market..............................................................................15  Itinerant drug seller..........................................................16  Community-oriented resource person…………………………..17  Other(specify)..................................................................18 | |
| Q606 | How many days after the illness began did you first seek advice or treatment for (NAME)? | | (00 if same day) |
| Q607 | At any time during the illness, did (NAME) have blood taken to diagnose malaria | | Yes…………………………..1  No……………………………2 |
| Q608 | Were you told by a healthcare provider that (NAME) had malaria? | | Yes…………………………..1  No……………………………2 |
| Q609 | Was (NAME) advised to take antimalarial drugs? | Yes…………………………………………………..……..1  No………………………………………………….……….2 | |
| Q610 | What drugs did (NAME) take?  Probe: Any other drugs? | Artemisinin Combination Therapy (Act)....…………1  SP/Fansidar………………………………………………………..2  Chloroquine……………………………………………………….3  Amodiaquine……………………………………………………..4  Quinine Pills……………………………………………………….5  Quinine Injection/IV…………………………………………..6  Artesunate Rectal..................................................7  Artesunate Injection/IV…..……………………...........8  Other Antimalarial….……………………………………….9  .Drug of unknown type ………………………….........10 | |
| Q611 | If (NAME) took antimalarial, where did you get the antimalarial drug(NAME) took? | Same facility visited as Q604.............................1  Other(specify)__________________________2 | |
| Q612 | How long after the fever started did(NAME) first take an artemisinin combination therapy? | Same Day...........................................................0  Next Day............................................................1  Two Days After Fever.........................................2  Three Or More Days After Fever........................3  Don't Know.........................................................4 | |
| **Note: If more than one household member was ill (Q601 >1) , kindly repeat Q603 – Q612 for each member** | | | |

| **SECTION 7: VECTOR CONTROL** | | |
| --- | --- | --- |
| Q700 | Have you ever heard of a mosquito net | Yes…………………………………….……………………1  No………………………………………….…………….…2 |
| Q701 | Does your household own a mosquito net | Yes……………………………………….……………….…1  No………………………………………….………………..2 |
| Q702 | How many mosquito nets does your household have? | /_____________/ |
| Q703 | How many months ago did your household get the mosquito net? | _____________ (00 if less than one month) |
| Q704 | Where did you get the net? | Distribution campaign.................................1  ANC..............................................................2  Immunization visit.......................................3  Govt. Health facility....................................4  Private health facility..................................5  Pharmacy....................................................6  Shop/market ..............................................7  Community health worker..........................8  Religious institution....................................9  School.........................................................10 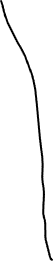 Other...........................................................11  Don’t know..................................................12 |
| Q705 | Did you pay for the net? | Yes…………………………………………………………...1  No………………………………….………………..……....2  Not Sure…………………………………………………...3 |
| Q706 | If you paid for the net, how much did you pay? | ______________________ |
| Q707 | Is the mosquito net you have treated? | Yes……………………………………………………………1  No…………………………………………..……………….2  Not Sure……………………………………………..……3 |
| Q708 | If yes, what was it treated with? |  |
| Q709 | Did anyone sleep inside this  mosquito net last night? | Yes………………………………………………………….1  No……………………………………………...............2  Not Sure………………………………………….….....3 |
| Q710 | ***For each net in the household, ask the following question***  Who slept inside this mosquito  net last night? | NAME.............................................................. |
| Q711 | Why did noone sleep  inside this net? | No mosquitoes…………………………...................1  No malaria.................................................... 2  Too hot..........................................................3  Difficult to hang...……………………………...........4  Don't like smell.............................................5  Feel `closed in' or constrained.....……………...6  Net too old/torn.……………………………..………..7  Net too dirty.................................................8  Net not available last night (washing)..……..9  Feelitn chemicals are unsafe ......................10  ITN provokes cough……………………….............11  Users did not sleep here last night…..………..12  Net not needed last night.............................13  No space to hang …………………….………………..14  Other (specify) .............................................15  Don’t know…………………………………………………99 |
| Q712 | Have you ever heard of indoor residual spraying (IRS)?  **If no, explain what IRS means to study participant** | Yes……………………………………..………1  No……………………………………….……..2 |
| Q713 | If yes, what does it mean?  **If not explained accurately, explain what IRS means to study participant** |  |
| Q714 | What is the importance of IRS? | Prevention of mosquito bite……………………..1  Prevention of other insect bite………………….2  Prevention of scorpion stings…………………...3  Others (specify)……………………………………..…4 |
| Q715 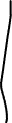 | What do you think are the disadvantages of IRS | Respiratory disorder…………………………………1  Headache………………………………………….……..2  Food contamination…………………………………3  Discoloration of surfaces and walls………….4  Unpleasant odor……………………………………..5  Others(Specify)_____________ |
| Q716 | Have IRS ever been conducted in this household | Yes……………………………………………..............1  No………………………………………………………….2 |
| Q717 | If yes, when was the last time this was done | Month /______/ Year/_____/ |
| Q718 | If yes, who conducted the IRS | Government……………………………………….…1  Self…………………………………………………..……2  Community…………………………………………..3  Health Facility…………………………..………….4  Others(Specify) …………………………………...5 |
| Q719 | Do you have window/door screens? | Yes………………………………………………………1  No……………………………………………............2 |
| Q720 | Do you use spray insecticides? | Yes………………………………………………………1  No…………………………………………………….…2 |
| Q721 | If yes, how often do you use it? |  |
| Q722 | Do you use mosquito coil? | Yes……………………………………………………...1  No……………………………………………………….2 |
| Q723 | If yes, how often do you use it? | Daily………………………………….………………..1  Sometimes………………………………………....2  Rarely…………………………………………..…….3 |
| Q724 | What other things do you do to prevent malaria infection in your household? |  |
| Q725 | Have you heard of seasonal malaria chemoprevention (SMC)?  **If no, explain what SMC means to study participant** | Yes……………………………………………….…...1  No……………………………………………..………2 |
| Q726 | If yes, what does it mean?  **If not explained accurately, explain what SMC means to study participant** |  |
| Q727 | ***(Check line listing for how many children aged 3-59 months)***  Just to confirm you have /____/  children aged 3-59 months in this household? | Yes………………………………………………...1  No………………………………………………….2 |
| Q728 | If yes, has any of them ever taken SMC | Yes………………………………………………..1  No…………………………………………………2 |
| Q729 | If they have ever taken SMC, how long ago did they take it? | Less than 28 days ago……………………1  29-42 days ago………………………………2  43 days or more……………………………3  Can’t remember……………………………4 |
| Q730 | If they have ever taken SMC, how many doses have they taken in the last four months? | ........................................................................... |
| Q731 | If they have taken SMC, who provided the SMC? | Government....................................1  NGO. ……………………………………………2  Community…………………………………..3  Health Facility……………………………….4  Others(Specify) _______________ 5 |

| **SECTION 8: MOBILITY PATTERNS AND INSIDE/OUTDOOR ACTIVITIES** | | |
| --- | --- | --- |
| Q800 | How long have you been living continuously in this community?  ***Ask for duration in years*** | (00 If less than one year) |
| Q801 | Just before you moved here, how would you describe where you lived? | City ……………………......................................……………….1  Town……………………………………………………….......………...2  Rural………………………………………………………..……………...3 |
| ***For each person in the household ask the following questions*** | | |
| Q802 | Have you travelled out of your current residence in the last 4 weeks | Yes………………………………………………………..…..……………1  No…………………………………………………………..………………2 |
| Q803 | How many times have you travelled out of your current location in the last 4 weeks? | ................................................................................................ |
| Q804 | If yes, where did you travel to? | Outside my community…………………………………………..1  Outside my State…………………………………………………….2  Others(Specify)…………………………………………………….…3 |
| Q805a | Kindly provide the state, LGA and the ward of place visited, if known? | State ……....................................  LGA ………..................................  Ward……..................................... |
| Q805b | If Q805a isn’t known, Can you provide the address of the place visited? | Address:  ________________________________________________________ |
| Q806 | What was the main reason for your travel? | Business/Work-Related………………………………………….…1  Holiday……………………………………………………….…….........2  Visiting Family/Relatives…………………………………………..3  Visit Friends……………………………………………….……….......4  Others(Specify)______________________________5 |
| Q807 | How best would you describe your travel location? | Urban area - formal settlement ……………………………….1  Urban area - Informal settlement or slum…………………2  Rural area - village………………………………………………..3  Rural area – farm ………………..……………………………….4  Others(Specify)____________________________5 |
| Q808 | Did you stay overnight during your last travel? | Yes………………………………………………………………………1  No……………………………………………………………………….2 |
| Q809 | How long did you stay at this location? | .................................................................................................... |
| Q810 | Did you sleep under a mosquito net during the last night you travelled? | Yes…………………….,…………………………………………......1  No………………………………………………………………………2 |
| Q811 | If no, what was the reason? | No mosquitoes...……………………………………………………..1  No malaria....……………………………………………................2  Too hot……………………..…………………………………………....3  Difficult to hang . …………………………………………………....4  Don't like smell . ………………………………………………………5  Feel “closed in’' or constrained ………………………………..6  Net too old/torn ……………………………………………….........7  Net too dirty. . . . . . . . …………………………………………......8  Net not available last night (washing)...…………………….9  Feel ITN chemicals are unsafe.....……………………….........10  ITN provokes cough…………………………………………….......11  Net not needed last night. . . . …………………………..........12  No space to hang .………………………………………….............13  Other (specify) _______________________________14  Don’t know.............………………………………………..............99 |
| Q812 | Did you treat malaria during your most recent travel? | Yes…………………….,………………………………………….............1  No……………………………………………………………………………..2 |
| Q813 | After you returned from your most recent trip, did you treat malaria? | Yes…………………….,………………………………………….............1  No……………………………………………………………………..........2 |
| Q814 | How many days after your most recent trip did you treat malaria? | _______________________________________________ |
| Q815 | Did you do any other thing to prevent malaria during your travel? | Yes…………………….,………………………………………….............1  No……………………………………………………………………..........2 |
| Q816 | If yes, what did you do? | Used Mosquito Coil……………………………………................1  Sprayed the room with insecticide ………………………..….2  Use Mosquito Repellant Cream………………………...........3  Others(Specify)_______________________________4 |
| Q817 | How long do you spend outside the home daily | /___________________/hours  /___________________/ minutes |
| Q818 | Do you do any chores outside your home | Yes………………………….…………………………………………........1  No……………………………………………………………………..........2 |
| Q819 | If yes, what kind of chores do you do? |  |
| Q820 | How long do you spend outside doing these chores | /___________________/hours  /___________________/ minutes |
| Q821 | What time do you usually go to bed? | 6:00pm- 7:00pm……………………………………………………....1  7:00pm – 8:00pm……………………………………………………...2  9:00pm-10:00pm……………………………………………………...3  10pm and above……………………………………………………....4 |
| Q822 | On a daily basis, would you say | I spend same duration of time indoor/outdoor…………................1  I spend more time indoor than outdoor…………………...................2  I spend more time outdoor than indoor…………………...................3  I cannot estimate the time I spend indoor/outdoor………............4  Others__________ |
| **If there are non-residents in the household who slept in the HH the night before, ask the following questions** | | |
| Q823 | How long in days has (NAME) been staying at your home | ______________________ |
| Q824 | Where is (NAME) visiting from? | State__________________  LGA___________________  Ward __________________ |
| Q825 | Has (NAME) had malaria at any time since they arrived? | Yes………………………………………………….……….…....1  No……………………………………………………………….…2 |
| Q826 | Was (NAME) diagnosed by a health professional? | Yes………………………………………………….…….,….....1  No…………………………………………………………………2 |

| **MALARIA TESTING ALGORITHM** | | | | |
| --- | --- | --- | --- | --- |
|  | SELECTED FOR MALARIA RDT TEST | Yes……………………………………………………..1  No………………………………………………………2 | | |
|  | ASK CONSENT FOR MALARIA TEST  FROM PARENT/OTHER ADULT. | "As part of this survey, we are asking eligible respondents to take a test to see if they have malaria. Malaria is a serious illness caused by a parasite transmitted by a mosquito bite. This survey will assist the government to develop programs to prevent malaria.  We ask all eligible respondents to take part in malaria testing in this survey and give a few drops of blood from a finger or heel. One blood drop will be tested for malaria immediately, and the result will be told to you right away. All results will be kept strictly confidential and will not be shared with anyone other than members of our survey team.  Do you have any questions?  You can say yes or no. It is up to you to decide.  Will you participate in the malaria test?"  Yes………………………………………………………1  No…………………………………………………….…2 | | |
|  | RDT DONE  Yes……1  No…….2 | OUTCOME:  TESTED……..........1  REFUSED…..………2  Other_________3 |  | RESULT:  POSITIVE…….1  NEGATIVE…...2  Other________3 |

### Health Facility Survey Instruments

#### Health facility data capture form

**HEALTH FACILITY INFORMATION**

This form should be completed for every health facility and linked to participant responses from the health facility surveys

NAME OF HEALTH FACILITY . . . . . . . . . . . . . . . . . . . . . . . . . . . . . . . . . . . . . . . . . . . . . . . . . . . . . . .

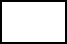
TYPE OF HEALTH FACILITY

Public . . . . . . . . . . . . . . . . . . . . . . . 1

Private. . . . . . . . . . . . . . . . . . . . . . . .2

PHYSICAL ADDRESS OF HEALTH FACILITY_______________________________________

_____________________________________________________________________________________________________________________________________________________

WARD ..............................................................

LGA ..................................................................

NUMBER OF ANC ATTENDEES LAST MONTH: _______________________

NUMBER OF DELIVERIES LAST MONTH: ____________________________

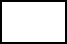
Does your facility conduct mosquito net distribution?

Yes……………………………………………………………………………1

No…………………………………………………………………………….2

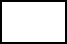
IPTp Available in Health Facility

Yes …………………………………………………………………………...1

No……………………………………………………………………………..2

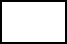
Involved in mass campaigns?

Yes …………………………………………………………………………...1

No……………………………………………………………………………..2

If involved in mass campaigns, list the common campaigns involved in

____________________________________________________________

____________________________________________________________

_____________________________________________________________

#### Health facility questionnaire

| **BACKGROUND INFORMATION** | | | |
| --- | --- | --- | --- |
| LOCAL GOVT. AREA…………………………………………………………………………....................... | | | |
| WARD……………………………………………………………………………………………………………. | | | |
| ENUMERATION AREA/CLUSTER NUMBER……………………………………………………… | | | |
| NAME OF HEALTH FACILITY ............................................................................................................... | | | |
| PHYSICAL ADDRESS OF HEALTH FACILITY...................................................................................... | | | |
| DATE: DAY……… MONTH………YEAR………… | | | |
| INTERVIEWER'S NAME ……………………………………. | | | |
| INTERVIEWERS PHONE NO…………………………………………… | | | |
| INTERVIEWER VISIT | | | |
|  | 1 | 2 | 3 |
| DATE | __________________ | ___________________ | ___________________ |
| RESULT |  |  |  |
| SUPERVISORS NAME | ___________________ | FIELD EDITOR | ___________________ |
| **INTRODUCTION AND CONSENT** My name is ……………and my colleagues are…………… I am working with the University of Ibadan. We would like your opinion on various issues to enable us understand malaria transmission in urban areas part of a collaborative project with Northwestern University, USA and National Malaria Elimination Programme. The information we collect will help the government to plan health services to prevent malaria infections by ensuring you receive suitable interventions. We thank you for honouring our presence. **Purpose** The purpose of this interview is to investigate malaria prevalence among pregnant women and members of their household and identify what factors predispose them to malaria infection **Procedure** As part of this study, you will be asked questions about your household characteristics, individuals living within the household, socio-demographic information, knowledge of malaria, risk factors and other relevant questions, which will help us achieve our objectives. You will also be offered rapid diagnostic testing for malaria infection. The interview should last between 30- 45 minutes. Your responses will be inputted into an electronic data-capturing device. The interview is going to be anonymous and confidential as much as possible. You can choose whether to participate in the interview, and you may stop at any time during the study. Your decision not to continue to participate will not attract any penalty. **Benefits** Your participation in this study may not provide any personal benefit to you. However, should you decide to participate in this study, you will be doing society a great service because the findings of this study will be useful in the design of interventions and programmes for the control and prevention of malaria in your community. **Risks** There are no known or anticipated risks associated with participation in this study beyond those experienced during an average conversation. If a question makes you uncomfortable, you can choose not to answer. **Confidentiality** The information you share will be kept confidential. Identifying information will be removed from the electronic data. The electronic data will be retained for a maximum of 5 years, after which they will be destroyed. Data will be stored in an encrypted folder on protected laptop. Only the research team will have access to study data. No identifying information will be used in any presentations or publications based on this research. **Contact** If you have any questions or concerns regarding this study, please contact: Professor IkeOluwapo Ajayi, Tel: 08023268431 Director, Institute for Advanced Medical Research and Training (IMARAT), College of Medicine, University of Ibadan Thank you for choosing to participate in the study. Kindly show by using any of the following 2 boxes, that your participation in this study was voluntary. I will participate I will not participate 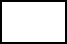Do you have any questions? Yes _______________1 No________________2 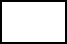May I begin the interview now? Yes _______________1 No________________2 | | | |

| **ELIGIBILITY** | | |
| --- | --- | --- |
| Q001 | Is this your first pregnancy? | Yes...................................................1  No.....................................................2 |
| Q002 | Is this your first ANC visit | Yes...................................................1  No.....................................................2 |
| Q003 | Have you been interviewed by the urban malaria study team before? | Yes……………………….……………1  No……………………………………...2 |
| Q004 | Where do you live? | Within Ibadan metropolis…………....1  Outside Ibadan Metro Area………….2  **Respondents living in Ibadan metropolis are those residing in any of the following LGAs – Ibadan North, Ibadan North West, Ibadan North East, Ibadan South West, Ibadan South West** |
|  | ***(Check, If Q001=1, Q002=1, Q003=2 and Q004 = 1, then woman is eligible for interview*** | Eligible…………………………….......1  Not Eligible………………………........2 |

**IF ELIGIBLE PROCEED TO INTERVIEW**

| **SECTION 1. HOUSEHOLD RESOURCES** | | |
| --- | --- | --- |
| Q100 | What is the main source of drinking water for members of your household?  (Enter the number for the most commonly used) | **Improved sources**  Piped into dwelling/yard/plot -------------------------------1  Piped to neighbour ---------------------------------------------2  Public tap/standpipe -------------------------------------------3  Tube well or borehole -----------------------------------------4  Protected dug well ----------------------------------------------5  Protected spring -------------------------------------------------6  Rainwater ---------------------------------------------------------7  Tanker truck/cart with small tank --------------------------8  Bottled water ----------------------------------------------------9  **Unimproved source**  Unprotected dug well -----------------------------------------10  Unprotected spring --------------------------------------------11  Surface water (River, Pond) ----------------------------------12  Sachet water-----------------------------------------------------13  Others (specify)-------------------------------------------------14 |
| Q101 | What is the main source of water used by your household for other purposes such as cooking and  handwashing? | **Improved source**  Piped into dwelling/yard/plot ------------------------------1  Piped to neighbour --------------------------------------------2  Public tap/standpipe ------------------------------------------3  Tube well or borehole ----------------------------------------4  Protected dug well --------------------------------------------5  Protected spring -----------------------------------------------6  Rainwater -------------------------------------------------------7  Tanker truck/cart with small tank ------------------------8  Bottled water -------------------------------------------------9  **Unimproved source**  Unprotected dug well --------------------------------------10  Unprotected spring -----------------------------------------11  Surface water (River, Pond) ------------------------------12  Sachet water--------------------------------------------------13  **Others (specify)**----------------------------------------------14 |
| Q102 | Where is the main source of water located? | In own dwelling ----------------------------------------------1  Outside own dwelling --------------------------------------2  Public tap------------------------------------------------------3  Elsewhere (specify) -----------------------------------------4 |
| Q103 | On the average, how long does it take your household to get to the source of water and back? | ----------- Minutes  ------------Hours  ----------------Don’t Know |
| Q104 | What kind of toilet facilities do members of your family usually use? | **Improved sanitation facility**  Flush toilet ----------------------------------------------------1  Ventilated improved pit (VIP) latrine -------------------2  Pit latrine with slab -----------------------------------------3  Composting toilet -------------------------------------------4  **Unimproved facility**  Pit latrine without slab/open pit ------------------------5  Bucket ---------------------------------------------------------6  Hanging toilet/hanging latrine --------------------------7  Open defecation (no facility/bush/field) -------------8  Others (specify) --------------------------------------------98 |
| Q105 | Do you have your own toilet, or you share toilet with other households? | iii. Have own toilet------------------------------- 1 iv. Shared toilet -----------------------------------2 |
| Q106 | Where is the bathroom of your house located? | iv. Inside the house ----------------------------------1 v. Outside, separated from the house ----------2 vi. No bathroom at all--------------------------------3 |
| Q107 | How many rooms are used by members of your household for sleeping? | Specify the number of rooms used: ….………… |
| Q108 | How many members of your household sleep on the floor? | ***Give number, if none write 00***  _________________ |
| Q109 | What is the main source of power/energy use for cooking in your household? | Electricity -------------------------------------------1  LPG/natural gas/biogas -------------------------2  Kerosene -------------------------------------------3  Coal/lignite ----------------------------------------4  Charcoal --------------------------------------------5  Wood -----------------------------------------------6  Agricultural crop/straw/shrubs/grass ------7  Animal dung --------------------------------------8  Others specify ------------------------------------9 |
| Q110 | Which of the following items do you, your spouse or your family have? (Circle as appropriate) | \| Yes \| No \|; Radio \| 1 \| 2 \|; Television \| 1 \| 2 \|; Mobile telephone \| 1 \| 2 \|; Non-mobile telephone \| 1 \| 2 \|; Desktop Computer \| 1 \| 2 \|; Laptop Computer \| 1 \| 2 \|; Refrigerator \| 1 \| 2 \|; Table \| 1 \| 2 \|; Chair \| 1 \| 2 \|; Bed \| 1 \| 2 \|; Cupboard \| 1 \| 2 \|; Air conditioner \| 1 \| 2 \|; Electric iron \| 1 \| 2 \|; Generator \| 1 \| 2 \|; Fan \| 1 \| 2 \| |
| Q111 | Do your household own a farm land? | Yes-------------------------------------------------------------1  No--------------------------------------------------------------2 |
| Q112 | Is there a farm within the compound where you reside? | Yes...........................................................................1  No............................................................................2 |
| Q113 | Does any member of your household have livestock? | Yes -------------------------------------------------------------1  No--------------------------------------------------------------2 |
| Q114 | Type of housing | Face to face……………………………………………………..…1  One Bedroom………………………………………..………..…2  Two Bedroom Flat……………………………………………..3  Three Bedroom Flat……………………..……………………4  Duplex………………………………………………………….…...5  Other(specify)________________________________ |
| Q115 | Do you share your compound with other households? | Yes……………………………………………………………………1  No…………………………………………….………………..…...2 |

| **SECTION 2: NEIGHBOURHOOD CHARACTERISTICS** | | | | | | |
| --- | --- | --- | --- | --- | --- | --- |
| Q200 | How would you describe the road type in your neighbourhood | Tarred…………………………………………………………….1  Untarred…………………………………………………........2  Both Tarred/untarred…………………………………....3 | | | | |
| Q201 | Would you say most houses in your neighbourhood are? | Fenced with gate……………………………………………1  Fenced, no gate…………………………………………..…2  Partially Fenced………………………………………..……3  Partially Fenced………………………………………..……4  Other(specify)……………………………………………....5 | | | | |
| Q202 | If fenced, what is the nature of the fencing material | Cement Block………………………………………..……...1  Mud………………………………………………….…………..2  Barbed wire and cement block………………….....3  Others(specify)……………………………………………..4 | | | | |
| Q203 | Are the houses in your neighbourhood painted? | No not painted……………………………………………..1  Yes, old painting…………………………………………..2  Yes, recently painted…………………………………...3  Others (specify)…………………………………………...4 | | | | |
| Q204 | Are there open drainages in your neighborhood | Yes...................................................................1  No…………………………………………………….............2 | | | | |
| Q205 | Are the open drainages clogged with dirt or rubbish | Yes...................................................................1  No…………………………………………………….............2 | | | | |
| Q206: Thinking about the environment where you live, how much do you agree with the following statements | | | | | | |
|  |  | Strongly Agree | Agree | Can’t Say | Disagree | Strongly Disagree |
| Q206A | There is a lot of noise in my neighbourhood. | 1 | 2 | 3 | 4 | 5 |
| Q206B | There are sidewalks on most streets in my community. | 1 | 2 | 3 | 4 | 5 |
| Q206C | There is a lot of unpleasant smells in my neighbourhood. | 1 | 2 | 3 | 4 | 5 |
| Q206D | My neighbourhood has heavy human traffic. | 1 | 2 | 3 | 4 | 5 |
| Q206E | There is a lot of trash and litter on the street in my neighbourhood. | 1 | 2 | 3 | 4 | 5 |
| Q206F | There is vandalism in my neighbourhood. | 1 | 2 | 3 | 4 | 5 |
| Q206G | There are too many people hanging around on the streets near my home | 1 | 2 | 3 | 4 | 5 |
| Q206H | I have easy access to medical care in my neighbourhood. | 1 | 2 | 3 | 4 | 5 |

| **SECTION 3: INDIVIDUAL CHARACTERISTICS** | | | | | | |
| --- | --- | --- | --- | --- | --- | --- |
| Q300 | How long have you been living continuously in (CURRENT PLACE OF RESIDENCE)? ***(Criteria for selection is 1 year and above)*** | DURATION IN YEARS  _____________ | | | | |
| Q301 | How old were you at your last birthday?  PROBE TO GET ESTIMATE IF NOT SURE | AGE AT LAST BIRTHDAY (IN YEARS)  __________ | | | | |
| Q302 | Have you ever attended school?  (Ask all questions) | Quranic/Islamiyah school?  1. YES  2. NO | Adult classes?  1. YES  2. NO | | Formal school?  ]1. YES  2. NO | |
| Q303 | What is the highest level of formal school you completed? | Did not complete Primary School------------------------1  Primary Completed------------------------------------------2  Secondary Completed--------------------------------------3  Post-secondary School Completed----------------------4 | | | | |
| Q304 | If Post-secondary School completed: Specify highest level completed in this category: | If with Post-secondary Education, please, specify highest level completed: ________________________________ | | | | |
| Q305 | What is your ethnic group? | Igbo ------------------------------------------------1  Yoruba ---------------------------------------------2  Hausa ----------------------------------------------3  Others (specify)____________________ 4 | | | | |
| Q306 | What is your religion? | Christianity ---------------------------------------1  Islam -----------------------------------------------2  Traditional religion -----------------------------3  Other (specify) _____________________4 | | | | |
| Q307 | What is your current marital status? | Never married -------------------------------------1  Married --------------------------------------------- 2  Co-habiting ----------------------------------------- 3  Divorced/separated ------------------------------4  Widowed---------------------------------------------5 | | | | |
| Q308 | How would you describe the family living arrangement in your house? | Monogamous Family (a man and wife, with or without children) living in a separate house ---------1  Polygamous Family (a man, wives, and children) living in a separate house ---------------------------------2  Extended family (a family which extends across generations, i.e including grandparents, aunts, and other relatives) living in the same house ….3  A female-headed household (a woman and her children i.e. absence of a man in a household, thus, making the woman the sole economic provider for the family) .....................................................................4  Others (specify) ----------------------------- | | | | |
| Q309 | Who provides the main source of income in your home? | Self (the woman) …………………………………………….1  Husband/Partner ………………………………………….…2  Both Husband and Wife(s) provide equally………3  Parents ……………………………………………………………4  Children……………………………………………………….…..5  Others (specify) …………………………….…………….…..6 | | | | |
| Q310 | Do you currently do any work to earn an income? | Yes…………………………………….………………...1  No……………………………………………………......2 | | | | |
| Q311 | What kind of work do you do? | Professional/Technical/Managerial-------------------1  Clerical ---------------------------------------------------2  Sales and Services ---------------------------------------3  Skilled Manual --------------------------------------------4  Unskilled Manual ----------------------------------------5  Agriculture ------------------------------------------------6  Domestic Work-------------------------------------------7  Others (specify):------------------------------------------8 | | | | |
| Q312 | If you are an agricultural worker, what type of agricultural produce do you work with? | Rice .......................................................................1  Other plants...........................................................2  Poultry....................................................................3  Fish and other seafood...........................................4  Cows.......................................................................5  Other livestock.......................................................6  Other produce........................................................7 | | | | |
| Q313 | If you are an agricultural worker, is your work seasonal? | Yes..........................................................................1  No...........................................................................2 | | | | |
| Q314 | If your work is seasonal, what months of the year do you work? | ...................................................................................... | | | | |
| Q315 | Is there a farm within your household’s compound? | Yes...........................................................................1  No............................................................................2 | | | | |
| Q316 | Where do you do the work? | State.............................................................................  LGA..............................................................................  Ward............................................................................  _________________________________ | | | | |
| Q317 | How long do you spend indoor or outdoor at work?  **Ask for times and write response in hours** | At work (Overall)_____________________  Indoor at Work_______________________  Outside at Work______________________ | | | | |
| Q318 | How much do you earn? | Daily..............................................................  Weekly..........................................................  Monthly......................................................... | | | | |
| ***If currently, living with spouse/partner*** | | | | | | |
| Q317 | How old was your partner as at last birthday |  | | | | |
| Q318 | Has your partner ever attended school?  (Ask all questions) | Quranic/Islamiyah school?   1. YES 2. NO | | Adult classes?  1. YES  2. NO | | Formal school?  1. YES  2. NO |
| Q319 | What is the highest level of formal school you completed? | Did not complete Primary School ----1  Primary Completed-----------------------2 Secondary Completed--------------------3  Post-secondary School Completed----4 | | | | |
| Q320 | If post-secondary school completed: Specify highest level completed in this category: | If completed post-secondary education, please, specify highest level completed:  _________________________________ | | | | |
| Q320 | What is your husband’s/partner’s ethnic group? | Igbo --------------------------------------------1  Hausa------------------------------------------2  Yoruba ----------------------------------------3  Others (specify) -----------------------------4 | | | | |
| Q320 | What is your husband’s/partner’s religion? | Christianity -----------------------------------1  Islam -------------------------------------------2  Traditional African religion ----------------3  Others(specify)-------------------------------4 | | | | |
| Q321 | What is your partners current work status? | Working ---------------------------------------------1  Unemployed/looking for a job -----------------2  Retired ----------------------------------------------3  Studying --------------------------------------------4  Others (specify)-----------------------------------5 | | | | |
| Q322 | If working, what kind of work does your partner do? | Professional/Technical/Managerial----------1  Clerical----------------------------------------------2 Sales and Services -------------------------------3  Skilled Manual------------------------------------4  Unskilled Manual --------------------------------5  Agriculture ----------------------------------------6 Domestic Work-----------------------------------7  Other(specify)------------------------------------8 | | | | |
| Q323 | If your partner is an agricultural worker, is their work seasonal? | Yes..............................................................1  No...............................................................2 | | | | |
| Q324 | If your partner’s work is seasonal, what months of the year do they work? | ............................................................................ | | | | |
| Q325 | Where does your partner do their work? | State...................................................................  LGA....................................................................  Ward................................................................... | | | | |
| Q326 | How long does your partner spend indoor or outdoor at work? | At Work(Overall)________________________  Indoor at Work__________________________  Outside at Work_________________________  Don’t Know____________________________99 | | | | |

| **SECTION 4: KNOWLEDGE OF MALARIA TRANSMISSION, CAUSES AND PREVENTIVE PRACTICES** | | | | |
| --- | --- | --- | --- | --- |
| Q400 | Have you ever heard of malaria | Yes………………………………………………………1  No……………………………………………………….2 | | |
| Q401 |  |  | Yes | No |
|  | What was your source of information on malaria | Radio | 1 | 2 |
|  |  | Television | 1 | 2 |
|  |  | Newspaper | 1 | 2 |
|  |  | Friend | 1 | 2 |
|  |  | Health Care Worker | 1 | 2 |
|  |  | Religious leader | 1 | 2 |
|  |  | Colleague | 1 | 2 |
|  |  | Others (Specify)____________ |  | |
| Q402 | What are the common symptoms of malaria?  **(DO NOT READ OUT)** |  | Mentioned | Not Mentioned |
|  |  | Fever | 1 | 2 |
|  |  | Chills/Shivering | 1 | 2 |
|  |  | Headache | 1 | 2 |
|  |  | Joint Pain | 1 | 2 |
|  |  | Poor Appetite | 1 | 2 |
|  |  | Vomiting | 1 | 2 |
|  |  | Convulsion | 1 | 2 |
|  |  | Cough | 1 | 2 |
|  |  | Catarrh/Nasal Congestion | 1 | 2 |
|  |  | Don’t know any | 3 |  |
|  |  | Others (Specify) |  | |
| Q403 | What are the causes of malaria you know?  (**DO NOT READ OUT**) |  |  | |
|  |  | Mosquitoes bite | 1 | 2 |
|  |  | Dirty Environment | 1 | 2 |
|  |  | Stagnant Water | 1 | 2 |
|  |  | Lakes, pits, dams around surroundings | 1 | 2 |
|  |  | Bushes around the house | 1 | 2 |
|  |  | Ill ventilated houses | 1 | 2 |
| Q404 | Do you think malaria can be prevented? | Yes………………………………………………………1  No……………………………………………………….2 | | |
| Q405 | Which was do you think malaria can be prevented? |  | Yes | No |
|  |  | Sleeping under insecticide treated nets | 1 | 2 |
|  |  | Cutting bush around the house | 1 | 2 |
|  |  | Spraying mosquito insecticide in homes | 1 | 2 |
|  |  | Taking malaria preventive drug | 1 | 2 |
|  |  | Door/Window Screens | 1 | 2 |
|  |  | Taking herbs | 1 | 2 |
|  |  | By praying | 1 | 2 |

| **SECTION 5: MALARIA HISTORY AND TREATMENT SEEKING PRACTICES** | | | | |
| --- | --- | --- | --- | --- |
| Q500 | Have you been ill with fever at any time in the last 2 weeks? | | Yes-----------------------------------------------------------------1  No------------------------------------------------------------------2 | |
| Q501 | Have you been ill with malaria at any time in the last 2 weeks? | | Yes-----------------------------------------------------------------1  No------------------------------------------------------------------2 | |
| Q501 | Have anyone in your household been ill with fever in the last two weeks | | Yes-----------------------------------------------------------------1  No------------------------------------------------------------------2 | |
| Q502 | At any time during the illness, did anyone in your household have blood taken from them to diagnose malaria? | | Yes-----------------------------------------------------------------1  No------------------------------------------------------------------2 | |
| Q503 | Were you told by a healthcare provider that anyone in your household had malaria? | | Yes-----------------------------------------------------------------1  No------------------------------------------------------------------2 | |
| Q504 | How many people in your household were diagnosed with malaria? | | ..................................................................................... | |
| Q505 | Have you ever had malaria in this pregnancy | | Yes-----------------------------------------------------------------1  No------------------------------------------------------------------2 | |
| Q506 | If yes, at what gestational age was malaria diagnosed | | _________________________ | |
| Q507 | Regarding your last episode of malaria, where did you first seek advice or treatment | | **PUBLIC SECTOR**  Government hospital …………………………………….............1  Government health center...................…………..………....2  Government health post...............................................3  Mobile clinic...................................................................4  Fieldworker/CHW...........................................................5  Other public sector(specify)...........................................6  **PRIVATE MEDICAL SECTOR**  Private hospital/ private hospital/clinic..........................7  Pharmacy........................................................................8  Chemist/PMV..................................................................9  Private doctor.................................................................10  Mobile clinic ...................................................................11  Fieldworker/CHW............................................................12  Other private ………………………………….……….....................13  Medical sector ………………………………………..…..................14  **OTHER SOURCE**  Shop...................................................................................15  Traditional practitioner......................................................16  Market................................................................................17  Itinerant drug seller............................................................18  Community-oriented resource person…………………............19  Other(specify)……...............................................................20 | |
| Q508 | How many days after the illness began did you first seek advice or treatment? | | | (00 if same day) |
| Q509 | At any time during the illness, did (NAME) have blood taken to diagnose malaria | | | Yes…………………………..1  No……………………………2 |
| Q510 | What was the result? | | | Positive……………………....1  Negative…………………..…2  Indeterminate……………..3 |
| Q511 | Were you advised to take antimalarial drugs? | Yes………………………………………………..…..………..1  No……………………………………………………………….2 | | |
| Q512 | Where did you get the antimalarial drug you took? | Same facility visited as Q504.............................................1  Other(specify)__________________________________2 | | |
| Q513 | What drugs did you take?  Probe: Any other drugs? | Artemisinin Combination Therapy (Act)...........................1  SP/Fansidar.......................................................................2  Chloroquine……………………………………….………………………...3  Amodiaquine…………..…………………....................................4  Quinine Pills……………………………..…………………….……………5  Quinine Injection/IV…………..…………………………………….....6  Artesunate Rectal………………..……………………...…………......7  Artesunate Injection/IV……….……………………..………….......8  Other Antimalarial……………………………..………………….......9  Drug of unknown type …………………………………..……………10 | | |
| Q514 | How long after the fever started did you first take an antimalarial drug? | /________________________/ | | |

| **SECTION 6: VECTOR CONTROL** | | |
| --- | --- | --- |
| Q600 | Have you ever heard of a mosquito net | Yes……………………………………………..1  No………………………………………………2 |
| Q601 | Does your household own a mosquito net | Yes……………………………………………..1  No………………………………………………2 |
| Q602 | How many mosquito nets does your household have? | _____________ |
| Q603 | How many months ago did your household get the mosquito net? | _____________ (00 if less than one month) |
| Q604 | Where did you get the net? | Distribution campaign................................1  ANC.............................................................2  Immunization visit......................................3  Govt. Health facility....................................4  Private health facility.................................5  Pharmacy...................................................6  Shop/market..............................................7  Community health worker..........................8  Religious institution....................................9  School........................................................10  Other..........................................................11  Don’t know.................................................12 |
| Q605 | Did you pay for the net? | Yes……………………………………………………….….1  No………………………………..………………………....2  Not Sure………………………………………………..…3 |
| Q606 | If you paid for the net, how much did you pay? | ______________________ |
| Q607 | Is the mosquito net you have treated? | Yes……………………………………………………..……1  No………………………………..……………………….…2  Not Sure…………………………….……………………3 |
| Q608 | If yes, what was it treated with? |  |
| Q609 | Did you sleep under a  mosquito net last night? | Yes………………………………………………………….1  No…………………………………………….………….…2  Not Sure………………………………………..………..3 |
| Q610 | Did any member of your household sleep under a net last night | Yes………………………………………………………….1  No…………………………………………………..………2 |
| Q611 | How many members of your household slept under a net last night? | ..................................................... |
| Q612 | How many people live in your household? | ..................................................... |
| Q613 | If you did not sleep under a mosquito net last night, what are the reasons? | No mosquitoes………………………………………………………….1  No malaria…………………………………………………………….…..2  Too hot……………………………………………………………………..3  Difficult to hang…………………………………………………………4  Don't like smell…..……………………………………………..........5  Feel “closed in’' or constrained.…………………………………6  Net too old/torn………………………………………………………...7  Net too dirty……………..………………………………………………..8  Net not available last night (washing)………………………….9  Feel ITN chemicals are unsafe……………………………………..10  ITN provokes cough……………………………………………………..11  Net not needed last night…………..………………………................12  No space to hang……………………………………………………………....13  Other (specify) ____________________________________14  Don’t know…………….…………….…………………………....................99 |
| Q614 | Have you ever heard of indoor residual spraying(IRS)?  **If no, explain what IRS means to study participant** | Yes…………………………………………………….………1  No…………………………………………………….……….2 |
| Q615 | If yes, what does it mean?  **If not explained accurately, explain what IRS means to study participant** |  |
| Q616 | What is the importance of IRS? | Prevention of mosquito bite…………………………………….1  Prevention of other insect bite……………………….……….2  Prevention of scorpion stings…………………………………..3  Others (specify)……………………………………………………….4 |
| Q617 | What do you think are the disadvantages of IRS | Respiratory disorder…………………………….…………………1  Headache……………………………………….…………………..….2  Food contamination………………………….……………..…….3  Discoloration of surfaces and walls…………………………4  Unpleasant odor………………………………….………………...5  Others (specify)_____________________________6 |
| Q618 | Have IRS ever been conducted in your household | Yes……………………………..………….…………………………..1  No………………………………………………………………………2  Not sure……………………………………………………………..3 |
| Q619 | If yes, when was the last time this was done | Month /______/ Year/_____/ |
| Q620 | If yes, who conducted the IRS | Government………………………………………………………1  Self…………………………………………………………………….2  Community………………………………………………………..3  Health Facility…………………………………………………….4  Others(Specify) ___________________________5 |
| Q621 | Do you have window/door screens? | Yes……………………………………………………………...1  No……………………………………………………………… 2 |
| Q622 | Do you use spray insecticides? | Yes………………………………………………………………1  No……………………………………………………………….2 |
| Q623 | If yes, how often do you use it? | Daily…………………………………………………………….1  Sometimes…………………………………………………..2  Rarely…………………………………………………………..3 |
| Q624 | Do you use mosquito coil? | Yes……………………………………………………………....1  No…………………………………………………….…………..2 |
| Q625 | If yes, how often do you use it? | Daily…………………………………………………………….1  Sometimes……………………………………………………2  Rarely…………………………………………………………..3 |
| Q626 | What other things do you do to prevent malaria infection in your household? |  |
| Q627 | Have you heard of seasonal malaria chemoprevention (SMC)?? | Yes…………………………………………………….………...1  No………………………………………………….…………….2 |
| Q628 | If yes, what does it mean?  **If not explained accurately, explain what IRS means to study participant** |  |
| Q629 | Do you have children aged 3-59 months in your household? | Yes……………………………………………….……………...1  No…………………………………………………..……………2 |
| Q630 | If yes, have they ever taken SMC | Yes……………………………………………………………..1  No…………………………………………………………….…2 |
| Q631 | If they have ever taken SMC, how long ago did they take it? | Less than 28 days ago………………………………….…1  29-42 days ago…………………………………………….…2  43 days or more……………………………………………..3  Can’t remember………………………….………………….4 |
| Q632 | If yes, who provided the SMC? | Government………………………………………………..…. 1  NGO.………………………………………………………………..2  Community……………………………………………………...3  Health Facility…………………………………………………..4  Others(Specify) __________________________ 5 |

| **SECTION 7: MOBILITY PATTERNS AND INSIDE/OUTDOOR ACTIVITIES** | | |
| --- | --- | --- |
| Q700 | How long have you been living continuously in this community?  ***Ask for duration in years*** | (00 If less than one year) |
| Q701 | Just before you moved here, how would you describe where you lived? | City …………………….....................................………………..1  Town……………………………………………………….......….2  Rural………………………………………………………..……….3 |
| Q702 | Have you travelled out of your current location in the last 4 weeks | Yes………………………………………………………..…..……..1  No…………………………………………………………..…………2 |
| Q703 | How many times have you travelled out of your current location in the last 4 weeks? |  |
| **Ask participant to provide responses to the following questions based on their most recent travel** | | |
| Q704 | If yes, for your most recent travel, where did you travel to? | Outside my community…………………………………1  Outside my State…………………………………………..2  Others(Specify)………………………………………..……3 |
| Q705a | Kindly provide the state, LGA and the ward of place visited, if known? | State ……..  LGA ……….  Ward……… |
| Q705b | If Q805a isn’t known, can you provide the address of the place visited? | Address:  ________________________________________________________ |
| Q706 | What was the main reason for your travel? | Business/Work-Related………………………………….…1  Holiday…………………………………….……………….……...2  Visiting Family/Relatives…………………………………...3  Visit Friends……………………………………………….………4  Others(Specify)___________________________5 |
| Q707 | How best would you describe your travel location? | Formal setting…………………………………………………. 1  Informal setting…………………………………………….....2  Slum………………………………………………………………….3  Farm………………………………………………………………… 4  Open Field……………………………..............................5  Others(Specify)___________________________6 |
| Q708 | Did you stay overnight during your last travel? | Yes…………………………………………………………………..1  No……………………………………………………………………2 |
| Q709 | How long did you stay at this location? |  |
| Q710 | Did you sleep under a mosquito net during the last night you travelled? | Yes…………………….,………………………………………....1  No…………………………………………………………………..2 |
| Q711 | If no, what was the reason? | No mosquitoes…………….………………………………...1  No malaria............………………………………………....2  Too hot……………..……………………………………………3  Difficult to hang……………………………………………..4  Don't like smell…………………………………………......5  Feel “closed in’' or constrained………………………6  Net too old/torn ……………………………………………7  Net too dirty…….…………………………………………...8  Net not available last night (washing)…………….9  Feel ITN chemicals are unsafe……………….........10  ITN provokes cough………………………………………11  Net not needed last night……………………….......12  No space to hang…………..……………………………..13  Other (specify)_________________________14  Don’t know….…………………………………...............99 |
| Q712 | Did you treat malaria during your most recent travel? | Yes…………………….……………………………………........1  No…………………………………………………………………..2 |
| Q713 | After you returned from your most recent trip, did you treat malaria? | Yes……………………..……………………………………….... 1  No……………………………………………………………………2 |
| Q714 | How many days after your most recent trip did you treat malaria? | __________________________________________ |
| Q715 | Did you do any other thing to prevent malaria during your travel? | Yes…………………….……………………………………….......1  No……………………………………………………………………2 |
| Q716 | If yes, what did you do? | Used Mosquito Coil……………………………………......1  Sprayed the room with insecticide ………………….2  Use Mosquito Repellant Cream……………………….3  Others(Specify)__________________________4 |
| Q717 | How long do you spend outside the home daily | /___________________/hours  /___________________/ minutes |
| Q718 | Do you do any chores outside your home | Yes…………………….………………………………………......1  No……………………………………………………………………2 |
| Q719 | If yes, what kind of chores do you do? |  |
| Q720 | How long do you spend outside doing these chores | /___________________/hours  /___________________/ minutes |
| Q721 | What time do you usually go to bed? | 6:00pm- 7:00pm………………………………………………1  7:00pm – 8:00pm…………………………………………….2  9:00pm-10:00pm…………………………………………….3  10pm and above……………………………………………..4 |
| Q722 | On a daily basis, would you say | I spend same duration of time indoor/outdoor…….1  I spend more time indoor than outdoor…...............2  I spend more time outdoor than indoor……………..3  I cannot estimate the time I spend indoor/outdoor......................................................4  Others___________________________________ |

| **MALARIA SCREENING** | | | |
| --- | --- | --- | --- |
|  | ASK CONSENT FOR MALARIA TEST  FROM PARENT/OTHER ADULT. | "As part of this survey, we are asking eligible respondents to take a test to see if they have malaria. Malaria is a serious illness caused by a parasite transmitted by a mosquito bite. This survey will assist the government to develop programs to prevent malaria.  We ask all eligible respondents to take part in malaria testing in this survey and give a few drops of blood from a finger or heel. One blood drop will be tested for malaria immediately, and the result will be told to you right away. All results will be kept strictly confidential and will not be shared with anyone other than members of our survey team.  Do you have any questions?  You can say yes or no. It is up to you to decide.  Will you participate in the malaria test?"  Yes……………………………………………………………….…1  No………………………………………………………………..….2 | |
|  | RDT DONE  Yes……1  No…….2 | OUTCOME:  TESTED……...…..1  REFUSED…..……2  Other________3 | RESULT:  POSITIVE…….1  NEGATIVE…..2  Other_____________3 |

### Longitudinal Study Instruments

#### Longitudinal Survey Baseline Questionnaire

| **BACKGROUND INFORMATION** | | | |
| --- | --- | --- | --- |
| LOCAL GOVT. AREA………………………………………………………………………..................... | | | |
| WARD……………………………………………………………………………………………………………... | | | |
| ENUMERATION AREA/CLUSTER NUMBER………………………………………………........... | | | |
| HOUSEHOLD NUMBER.......................................................................................... | | | |
| NAME OF HOUSEHOLD HEAD_________________________________________________________________ | | | |
| DATE: DAY………….. MONTH…………..YEAR………… | | | |
| INTERVIEWER'S NAME ……………………………………. | | | |
| INTERVIEWERS PHONE NO……………………………… | | | |
| INTERVIEWER VISIT | | | |
|  | 1 | 2 | 3 |
| DATE | __________________ | ___________________ | ___________________ |
| RESULT |  |  |  |
| SUPERVISORS NAME | ___________________ | FIELD EDITOR | ___________________ |
| **INTRODUCTION AND CONSENT** My name is ……………and my colleagues are…………… I am working with the University of Ibadan. We would like your opinion on various issues to enable us understand malaria transmission in urban areas as part of a collaborative project with Northwestern University, USA and Nigeria National Malaria Elimination Programme. The information we collect will help the government to plan health services to prevent malaria infections by ensuring you receive suitable interventions. We thank you for honouring our presence. **Purpose** The purpose of this interview is to investigate malaria cases among children and identify what factors predispose them to malaria infection. **Procedure** As part of this study, you will be asked questions about your household characteristics, children living within your household, socio-demographic information, knowledge of malaria, risk factors and other relevant questions which will help us achieve our objectives. In addition, one child aged between 0-10 years will be selected in your household for rapid diagnostic testing for malaria infection. We shall visit the child once a month to further ask questions relating to malaria infection and prevention and test them using the same rapid diagnostic tests. During each visit, the interview should last between 30-45 minutes. Your responses will be inputted into an electronic data capturing device. The interview is going to be anonymous and confidential as much as possible. You can choose whether to participate in the interview, and you may stop at any time during the study. There is no right or wrong answer, so feel free to express yourself. Remember your participation in this interview is voluntary. Your decision not to continue to participate will not attract any penalty. **Benefits** Your participation in this study may not provide any personal benefit to you. However, should you decide to participate in this study, you will be doing society a great service because the findings of this study will be useful in the design of interventions and programmes for the control and prevention of malaria in your community. **Risks** There are no known or anticipated risks associated with participation in this study beyond those experienced during an average conversation. If a question, or the discussion, makes you uncomfortable, you can choose not to answer. **Confidentiality** The information you share will be kept confidential. Identifying information will be removed from the electronic data. The electronic data will be retained for a maximum of 5 years, after which they will be destroyed. Data will be stored in an encrypted folder on protected laptop. Only the research team will have access to study data. No identifying information will be used in any presentations or publications based on this research. **Contact** If you have any questions or concerns regarding this study, please contact: Professor IkeOluwapo Ajayi, Tel: 08023268431 Director, Institute for Advanced Medical Research and Training (IMARAT), College of Medicine, University of Ibadan Thank you for choosing to participate in the study. Kindly show by using any of the following 2 boxes, that your participation in this study was voluntary. I will participate I will not participate Do you have any questions? Yes ___________No_________________ May I begin the interview now? Yes ___________No________________ | | | |

| **ELIGIBILITY** | | |
| --- | --- | --- |
| Q001 | Do you have children 0 – 10 years old residing in your household? | Yes...................................................1  No.....................................................2 |
| Q002 | Do you plan to live in your current place of residence for the next one year? | Yes……………………….……………1  No……………………………………...2 |
|  | ***(If Q001=1 and Q002=1, then household is eligible for interview)*** | Eligible…………………………….......1  Not Eligible………………………........2 |

**IF ELIGIBLE PROCEED TO INTERVIEW**

| **HOUSEHOLD LINE LISTING** | | | | | | |
| --- | --- | --- | --- | --- | --- | --- |
| Line No | Usual Residents | Relationship to Household Head (HH) | Sex | Residence | Age | Remarks |
|  | Please give me the names of all persons who usually live here | What is the relationship of (NAME) to the HH | Is (NAME) male or female?  Male = 1  Female = 2 | Did (NAME) sleep here last night?  Yes =1  No = 2 | How old was  (NAME) as at last birthday? |  |
| 1 |  |  |  |  |  |  |
| 2 |  |  |  |  |  |  |
| 3 |  |  |  |  |  |  |
| 4 |  |  |  |  |  |  |
| 5 |  |  |  |  |  |  |
| 6 |  |  |  |  |  |  |
| 7 |  |  |  |  |  |  |
| 8 |  |  |  |  |  |  |
| 9 |  |  |  |  |  |  |
| 10 |  |  |  |  |  |  |
| **Information on non-residents** | | | | | | |
| Line No | Non-residents | Relationship to HH | Sex | Residence | Age | Remarks |
|  | Please give me the names of all persons who don’t usually live here | What is the relationship of (NAME) to the HH | Is (NAME) male or female?  Male = 1  Female = 2 | Did (NAME) sleep here last night?  Yes =1  No = 2 | How old was  (NAME) as at last birthday? |  |
| 1 |  |  |  |  |  |  |
| 2 |  |  |  |  |  |  |
| 3 |  |  |  |  |  |  |
| 4 |  |  |  |  |  |  |

| CODES: |  |
| --- | --- |
| **RELATIONSHIP TO HEAD OF HOUSEHOLD** |  |
| 01 = HEAD | 09 = BROTHER-IN-LAW/SISTER IN-LAW |
| 02 = WIFE OR HUSBAND | 10 = NIECE/NEPHEW BY BLOOD |
| 03 = SON OR DAUGHTER | 11 = NIECE/NEPHEW BY MARRIAGE |
| 04 = SON-IN-LAW OR DAUGHTER-IN-LAW | 12 = OTHER RELATIVE |
| 05 = GRANDCHILD | 13 = ADOPTED/FOSTER/ STEPCHILD |
| 06 = PARENT | 14 = NOT RELATED |
| 07 = PARENT-IN-LAW | 15 = CO-WIFE |
| 08 = BROTHER OR SISTER | 16 = NANNY |
|  | 98 = DON'T KNOW |

| **SECTION 1. HOUSEHOLD RESOURCES** | | | | |
| --- | --- | --- | --- | --- |
| Q100 | What is the main source of drinking water for members of your household?  (enter the number for the most commonly used) | **Improved source**  Piped into dwelling/yard/plot -------------------------1  Piped to neighbour --------------------------------------2  Public tap/standpipe ------------------------------------3  Tube well or borehole ----------------------------------4  Protected dug well --------------------------------------5  Protected spring -----------------------------------------6  Rainwater --------------------------------------------------7  Tanker truck/cart with small tank -------------------8  Bottled water ---------------------------------------------9  **Unimproved source**  Unprotected dug well ----------------------------------10  Unprotected spring -------------------------------------11  Surface water (River, Pond)---------------------------12  Sachet water----------------------------------------------13  Others (specify)------------------------------------------14 | |  |
| Q101 | What is the main source of water used by your household for other purposes such as cooking and  handwashing? | **Improved source**  Piped into dwelling/yard/plot -----------------------1  Piped to neighbour -------------------------------------2  Public tap/standpipe -----------------------------------3  Tube well or borehole ---------------------------------4  Protected dug well -------------------------------------5  Protected spring ----------------------------------------6  Rainwater ------------------------------------------------7  Tanker truck/cart with small tank ------------------8  Bottled water --------------------------------------------9  **Unimproved source**  Unprotected dug well --------------------------------10  Unprotected spring -----------------------------------11  Surface water (River, Pond) ------------------------12  Sachet water--------------------------------------------13  Others (specify)----------------------------------------14 | |  |
| Q102 | Where is the main source of water located? | In own dwelling ---------------------------------------1  Outside own dwelling -------------------------------2  Public tap-----------------------------------------------3  Elsewhere (specify) ----------------------------------4 | |  |
| Q103 | On the average, how long does it take your household to get to the source of water and back? | ----------- Minutes  ------------Hours  ------------Don’t Know | |  |
| Q104 | What kind of toilet facilities do members of your family usually use? | **Improved sanitation facility**  Flush toilet ---------------------------------------------1  Ventilated improved pit (VIP) latrine ------------2  Pit latrine with slab ----------------------------------3  Composting toilet ------------------------------------4  **Unimproved facility**  Pit latrine without slab/open pit -----------------5  Bucket --------------------------------------------------6  Hanging toilet/hanging latrine -------------------7  Open defecation (no facility/bush/field) ------8  Others (specify) --------------------------------------98 | |  |
| Q105 | Do you have your own toilet, or you share toilet with other households? | Have own toilet---------------------------------------1  Shared toilet ------------------------------------------2 | |  |
| Q106 | Where is the bathroom of your house located? | Inside the house -------------------------------------1  Outside, separated from the house -------------2  No bathroom at all-----------------------------------3 | |  |
| Q107 | How many rooms are used by members of your household for sleeping? | Specify the number of rooms used: ….…………...... | |  |
| Q108 | How many members of your household sleep on the floor? | ***Give number, if none write 00*** | |  |
| Q109 | What is the main source of power/energy use for cooking in your household? | Electricity ----------------------------------------------1  LPG/natural gas/biogas ----------------------------2  Kerosene ----------------------------------------------3  Coal/ignite --------------------------------------------4  Charcoal -----------------------------------------------5  Wood --------------------------------------------------6  Agricultural crop/straw/shrubs/grass ---------7  Others specify ---------------------------------------8 | |  |
| Q110 | Which of the following items do you, your spouse or your family have? (Circle as appropriate) |  | |  |
|  |  |  | Yes | No |
|  |  | Radio | 1 | 2 |
|  |  | Television | 1 | 2 |
|  |  | Mobile telephone | 1 | 2 |
|  |  | Non-mobile telephone | 1 | 2 |
|  |  | Desktop Computer | 1 | 2 |
|  |  | Laptop Computer |  |  |
|  |  | Refrigerator | 1 | 2 |
|  |  | Table | 1 | 2 |
|  |  | Chair | 1 | 2 |
|  |  | Bed | 1 | 2 |
|  |  | Cupboard | 1 | 2 |
|  |  | Air conditioner | 1 | 2 |
|  |  | Electric iron | 1 | 2 |
|  |  | Generator | 1 | 2 |
|  |  | Fan | 1 | 2 |
| Q111 | Do your household own a farmland? | Yes------------------------------------1  No-------------------------------------2 | |  |
| Q112 | Does any member of your household have livestock? | Yes -----------------------------------1  No------------------------------------2 | |  |
| Q113 | Type of housing | Face to face………………………….1  One Bedroom……………………….2  Two Bedroom Flat………………..3  Three Bedroom Flat……………..4  Duplex………………………………….5  Others(specify)…………………….6 | |  |
| Q114 | Do you share your compound with other households? | Yes………………………………………1  No……………………………………….2 | |  |
| Q115 | Are the eaves of the house or building occupied by this household open or closed?  **Observe and record** | Completely Open ...................1  Partially Open…......................2  Closed.....................................3 | |  |
| Q116 | Does the part of the house or building occupied by the household have a ceiling?  **Observe and record** | No, None ........................................................................1  Yes, Partial/Poorly Sealed/Worn Out.............................2  Yes, Complete and Sealed …..........................................3 | |  |
| Q117 | Is there a farm within the compound?  **Observe and record** | Yes...............................................................................1  No................................................................................2 | |  |

#### Longitudinal Survey Follow-up Questionnaire

| **BACKGROUND INFORMATION** | | | |
| --- | --- | --- | --- |
| LOCAL GOVT. AREA………………………………………………………………………… | | | |
| WARD……………………………………………………………………………………………. | | | |
| ENUMERATION ……………………………………………………………………………… | | | |
| HOUSEHOLD NUMBER . . . . . . . . . . . . . . . . . . . . . . . . . . . . . . . . . . . . . | | | |
| NAME OF CAREGIVER___________________________________________________________  _____________________________________________________________________________ | | | |
| NAME OF CHILD_______________________________________________________________  _____________________________________________________________________________ | | | |
| DATE: DAY………….. MONTH…………..YEAR………… | | | |
| INTERVIEWER'S NAME ……………………………………. | | | |
| INTERVIEWERS PHONE NO……………………………………………….. | | | |
| MONTH OF VISIT /_____________/ | | | |
| INTERVIEWER VISIT | | | |
|  | 1 | 2 | 3 |
| DATE | __________________ | ___________________ | ___________________ |
| RESULT |  |  |  |
| SUPERVISORS NAME | ___________________ | FIELD EDITOR | ___________________ |

| **SECTION 1: CHILD CHARACTERISTICS** 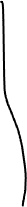 | | |
| --- | --- | --- |
| Q100 | Does (NAME) still attend school | Yes…………………………………...…………………..1  No…………………………………………………..…….2 |
| Q101 | What is (NAME) level of education now? | Preschool………………………………………….……1  Nursery……………………………………………….…2  Primary………………………………………………….3  Post Primary…………………………………………..4  Junior Secondary……………………………………5 |
| Q103 | Does your household have a mosquito net | Yes……………………………………………..……….1  No………………………………………….……………2  Had a net as at last visit………………………3 |
| Q104 | How many mosquito nets does your household now have? | /___________/ |
| Q105 | Where did you get the net? | Mass distribution campaign………………………..1  ANC…………………………………………………………….2  Immunization visit………………………………………3  Govt. Health facility…………………………………….4  Private health facility………………………………….5  Pharmacy…………………………………………………..6  Shop/market……………………………………………..7  Community health worker…………………………8  Religious institution…………………………………..9  School……………………………………………………….10 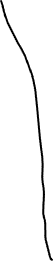 Other…………………………………………………………11  Don’t know……………………………………………….12 |
| Q106 | Did you pay for the net? | Yes………………………………………………………..1  No………………………………….………………….....2  Not Sure………………………………………………..3 |
| Q107 | How much did you pay? | ______________________ |
| Q108 | Is the mosquito net you have treated? | Yes………………………………………………………1  No………………………………………………….……2  Not Sure………………………………..………......3 |
| Q109 | If yes, what was it treated with? | ................................................................................... |
| Q110 | Did (NAME) sleep under the mosquito net last night | Yes……………………………………..……………….1  No…………………………………………………….…2 |
| Q111 | If no, why didn’t (NAME) sleep under a mosquito net | No mosquitoes…………………………………………………..1  No malaria………………………………………………………….2  Too hot………………………………………………………………3  Difficult to hang…………………………………………………4  Don't like smell………………………………………………....5  Feel `closed in' or constrained………………………..…6  Net too old/torn……………………………………………....7  Net too dirty……………………………………………………..8  Net not available last night (washing) ………………9  Feel ITN chemicals are unsafe……………………………10  Net provokes cough………………………………………….11  Users did not sleep here last night …………………..12  Net not needed last night…………………………………13  No space to hang ……………………………………..........14  Other (specify) __________________________15  Don’t know………………………………………………………99 |
| Q112 | Has (NAME) residence been sprayed in the last one months? | Yes……………………………………………………….1  No………………………………………………..………2 |
| Q113 | Who conducted the spraying? | Government………………………………………..1  Self……………………………………………….……..2  Community………………………………………….3  Health Facility………………………………………4  Others(Specify) ……………………………………5 |
| Q114 | What other things do you do to prevent malaria infection in your household? | Use of mosquito coil……………………………………..1  Use of mosquito repellant…………………………….2  Use of window/door screens…………………………3  Others(specify)_________________________4 |

| **SECTION 2: MALARIA IN THE ENROLLED CHILD** | | |
| --- | --- | --- |
| Now I would like to ask you about some questions about the health of your child aged 0 – 10 years  **(This is the child enrolled in the study)** | | |
| Q200 | Has (NAME) been ill with a fever at any time in the last four (4) weeks? | Yes…………………………………………………………….1  No……………………………………………………..………2 |
| Q201 | Has (NAME) been ill with malaria at any time in the last four (4) weeks? | Yes…………………………………………………………….1  No……………………………………………………..………2 |
| Q202 | At any time during the illness, did (NAME) have blood taken from them to diagnose malaria? | Yes…………………………………………………………….1  No………………………………………………..……………2 |
| Q203 | Were you told by a healthcare provider that (NAME) had malaria? | Yes…………………………………………………..……….1  No………………………………………………….…………2 |
| Q204 | Did you seek advice or treatment for (NAME) for the illness from any source? | Yes……………………………………………………………1  No……………………………………………………….……2 |
| Q205 | Where did you seek advice or treatment? | **PUBLIC SECTOR**  Government hospital...............…………………1  Government health centre..........................2  Government health post.............................3  Mobile clinic................................................4 Fieldworker/CHW.......................................5  Other public sector (Specify) ___________6  **PRIVATE MEDICAL SECTOR**  Private hospital/clinic..................................7  Pharmacy.....................................................8  Chemist/PMV...............................................9  Private doctor..............................................10  Mobile clinic ................................................11  Other private medical sector (specify)____12  **OTHER SOURCE**  Shop.............................................................13  Traditional Practitioner................................14  Market..........................................................15  Itinerant drug seller......................................16  Community-oriented resource person……….17  Other (Specify)…………………………………………..18 |
| Q206 | How many days after the illness began did you first seek advice or treatment for (NAME) | ____________________________________ |
| Q207 | Was (NAME) advised to take antimalarial drugs? | Yes………………………………………………………………..1  No…………………………………………………………………2 |
| Q208 | What drugs did (NAME) take?  Probe: Any other drugs? | Artemisinin Combination Therapy (Act)....……1  SP/Fansidar………………………………………………….2  Chloroquine…………………………………………………3  Amodiaquine……………………………………………….4  Quinine Pills…………………………………………………5  Quinine Injection/IV…………………………………….6  Artesunate Rectal………………………………………..7  Artesunate Injection/IV……………………………….8  Other Antimalarial………………………………………9  Drug of unknown type …………………………......10 |
| Q209 | If (NAME) took antimalarials, where did you get the antimalarial drug (NAME) took? | Same facility visited as Q204..................................1  Other(specify)_____________________________2 |
| Q210 | How long after the fever started did (NAME) first take an artemisinin combination therapy? | Same Day…………………………………………………..0  Next Day…………………………………………………….1  Two Days After Fever……………………..............2  Three Or More Days After Fever………………..3  Don't Know………………………………………………..8 |
| Q211 | ***Check If NAME is 3-59 months***  Has (NAME) taken SMC since our last visit? | Yes………………………………………………………………1  No…………………………………………………………….…2 |
| Q212 | How long ago did they take it? | Less than 7 days ago……………………………………1  7 - 14 days ago…………………………………………….2  15 - 28days …………………………..…………………….3  Can’t remember……………………….…………………4 |
| Q213 | If they have taken SMC, who provided the SMC? | Government……………………………………………….1  NGO.…………………………………………………………..2  Community…………………………………………………3  Health Facility……………………………………………..4  Others(Specify) _______________________5 |

| **SECTION 3: MOBILITY PATTERNS AND INSIDE/OUTDOOR ACTIVITIES** | | |
| --- | --- | --- |
| ***For each usual resident of the household ask the following questions*** | | |
| Q300 | Has (NAME) travelled out of your current location in the last 4 weeks | Yes……………………..…................................1  No……………………………………………….……….2 |
| Q301 | How many times has (NAME) travelled out of your current location in the last 4 weeks? | ................................................................. |
| **Ask participant to provide responses to the following questions based on their most recent travel** | | |
| Q302 | If yes, for your most recent travel where did (NAME) travel to? | Outside the community……………………………..…...1  Outside the State………………………………….….……..2  Others(Specify)………………………………………..………3 |
| Q303a | Kindly provide the state, LGA and the ward of place visited, if known? | STATE__________________________________________  LGA____________________________________________  Ward___________________________________________ |
| Q303b | If Q303a isn’t known, can you provide the address of the place visited? | Address:________________________________________ |
| Q304 | What was the main reason for your travel? | Buisness/Work-Related…………………………………….1  Holiday………………………………………………….………….2  Visiting Family/Relatives…………………………………..3  Visit Friends……………………………………………….…….4  Others(Specify)__________________________5 |
| Q305 | How best would you describe the travel location? | Urban area - formal settlement ………………………………....1  Urban area - Informal settlement or slum…………………...2  Rural area - village……………………………………………………….3  Rural area – farm ………………..………………………………………4  Others(Specify)________________________________5 |
| Q306 | Did (NAME) stay overnight during your last travel? | Yes……………………………………………………………..1  No………………………………………………………………2 |
| Q307 | How long did (NAME) stay at this location? | ......................................................................... |
| Q308 | Did (NAME) sleep under a mosquito net during the last night you travelled? | Yes………………………………………………….….,…....1  No………………………………………………………………2 |
| Q309 | If no, what was the reason? | No mosquitoes......……………………………………..1  No malaria...............……………………………………2  Too hot...........................................................3  Difficult to hang…………………………………………..4  Don't like smell……………………………………………5  Feel `closed in' or constrained…………………….6  Net too old/torn…………………………………………7  Net too dirty……………………………………………….8  Net not available last night (washing) ………..9  Feel ITN chemicals are unsafe……………………..10  ITN provokes cough…………………………………….11  Net not needed last night..............................12  No space to hang . ………………………………………13  Other (specify) _______________________14  Don’t know…………………………………………………99 |
| Q310 | Did (NAME) treat malaria during your most recent travel? | Yes…………………………………………………………….1  No……………………………………………………………..2 |
| Q311 | After (NAME) returned from the most recent trip, did (NAME) treat malaria? | Yes…………………………………………………………….1  No……………………………………………………………..2 |
| Q312 | How many days after (NAME) returned from the most recent trip did (NAME) treat for malaria | ___________________________ |
| Q313 | Did you do any other thing to prevent malaria during your travel? | Yes……………………………………………………………1  No…………………………………………………………….2 |
| Q314 | If yes, what did you do | Used Mosquito Coil…………………………………..1  Sprayed the room with insecticide …………...2  Use Mosquito Repellant Cream…………………3  Others(Specify)_______________________4 |
| Q315 | How long in hours does (NAME) spend outside the home daily? | ____________________________________ |
| Q316 | Does (NAME) do any chores outside the house now? | Yes…………………………………………………………….1  No……………………………………………………………..2 |
| Q317 | If yes, what kind of chores does (NAME) do? | ____________________________________ |
| Q318 | Approximately how long in minutes or hours does (NAME) spend outside doing these chores | ____________________________________ |
| Q319 | What time does (NAME) usually go to bed now? | 6.00pm-7.00pm. …………………………………………………1  8.00pm-9.00pm. …………………………………………………2  10.00pm above ………………………………………………… 3  Others(specify)________________________________________4 |
| Q320 | On a daily basis, would you say | I spend same duration of time indoor/outdoor…………................1  I spend more time indoor than outdoor…………………...................2  I spend more time outdoor than indoor…………………...................3  I cannot estimate the time I spend indoor/outdoor………............4  Others__________ |
| **If there are non-residents in the household who slept in the HH the night before, ask the following questions** | | |
| Q321 | How long in days has (NAME) been staying at your home | ______________________ |
| Q322 | Where is (NAME) visiting from? | State__________________  LGA___________________  Ward __________________ |
| Q323 | Has (NAME) had malaria at any time since they arrived? | Yes………………………………………………….……..…....1  No…………………………………………………………….…..2 |
| Q324 | Was (NAME) diagnosed by a health professional? | Yes………………………………………………….……..…....1  No…………………………………………………………….…..2 |

| **SECTION 4: MEASUREMENTS** | | |
| --- | --- | --- |
| Q400 | Weight |  |
| Q401 | Height |  |
| Q402 | Mid Upper Arm Circumference |  |
|  | ASK CONSENT FOR MALARIA TEST  FROM PARENT/OTHER ADULT. | "As part of this survey, we are asking eligible respondents to take a test to see if they have malaria. Malaria is a serious illness caused by a parasite transmitted by a mosquito bite. This survey will assist the government to develop programs to prevent malaria.  We ask all eligible respondents to take part in malaria testing in this survey and give a few drops of blood from a finger or heel. One blood drop will be tested for malaria immediately, and the result will be told to you right away. All results will be kept strictly confidential and will not be shared with anyone other than members of our survey team.  Do you have any questions?  You can say yes or no. It is up to you to decide.  Will you participate in the malaria test?"  Yes………………………………………………………1  No…………………………………………………….…2 |
| Q403 | Malaria RDT Done | Yes..........................................................1  No...........................................................2 |
| Q404 | Result | |
|  |  | Positive....................................................1  Negative..................................................2  Indeterminate.........................................3 |

### Entomological survey instruments

#### Community and Household Consent Form for Entomological Survey

We bring warm greetings from the management of Osun State University. This is an entomological study for the determination of indoor and outdoor biting rates of malaria in Kano and Ibadan to inform strategic plans to control malaria in your communities as part of a collaborative project with Northwestern University, USA and Nigeria National Malaria Elimination Programme.

**What does being a participating community involve?**

A team of Public Health Entomologists will work in the selected communities/households to collect both indoor and outdoor mosquitoes over a period of eight months. Their activities will include mounting of CDC light trap in selected rooms indoor and outdoor and between 6pm and 6am on each catching days for the collection of mosquitoes and indoor residual spraying of ten selected rooms in the community to collect indoor resting mosquitoes. The members of the community will be part of the personnel to be recruited for the study.

**What are the risks?**

There are no direct risks for participating in this study. However, there may be a situation of slight inconveniences in the household where the indoor and outdoor CDC light trap is carried out due to the activities of the entomologists collecting mosquitoes throughout the night.

**What are the benefits?**

The benefit is that your community/household would be contributing to the research in advancing efforts towards controlling malaria in urban Nigeria and the community/household may be first among the others to pilot and benefit from the control strategies emanating from the study.

**Confidentiality:**

Our team members are professionally trained, and all information and personal discussion volunteered to them during the study will be kept confidential.

**Voluntariness:**

Your participation in this study is purely voluntary. Your participation or withdrawal from the study will not have any negative effect whatsoever.

**Compensation:**

The community members or households selected as mosquito collectors to participate in the study will be remunerated in accordance with the budget for the study.

This study has been approved by the Federal Ministry of Health, Abuja Nigeria, and all questions should be directed to :

**Prof. M. A. Adeleke,**

Osun State University, Osogbo, Nigeria,

**Statement of consent**

I wish to confirm that I have read and understood the concept of the study. I therefore willingly agree to participate in the project on behalf of my household/community.

DATE: _________________SIGNATURE: _________________________________

NAME: _____________________________________________________________

POSITION IN COMMUNITY/HOUSEHOLD: __________________________________

Entomologist name and signature:

NAME AND SIGNATURE: ________________________________________

#### Sample modified CDC light trap data collection form

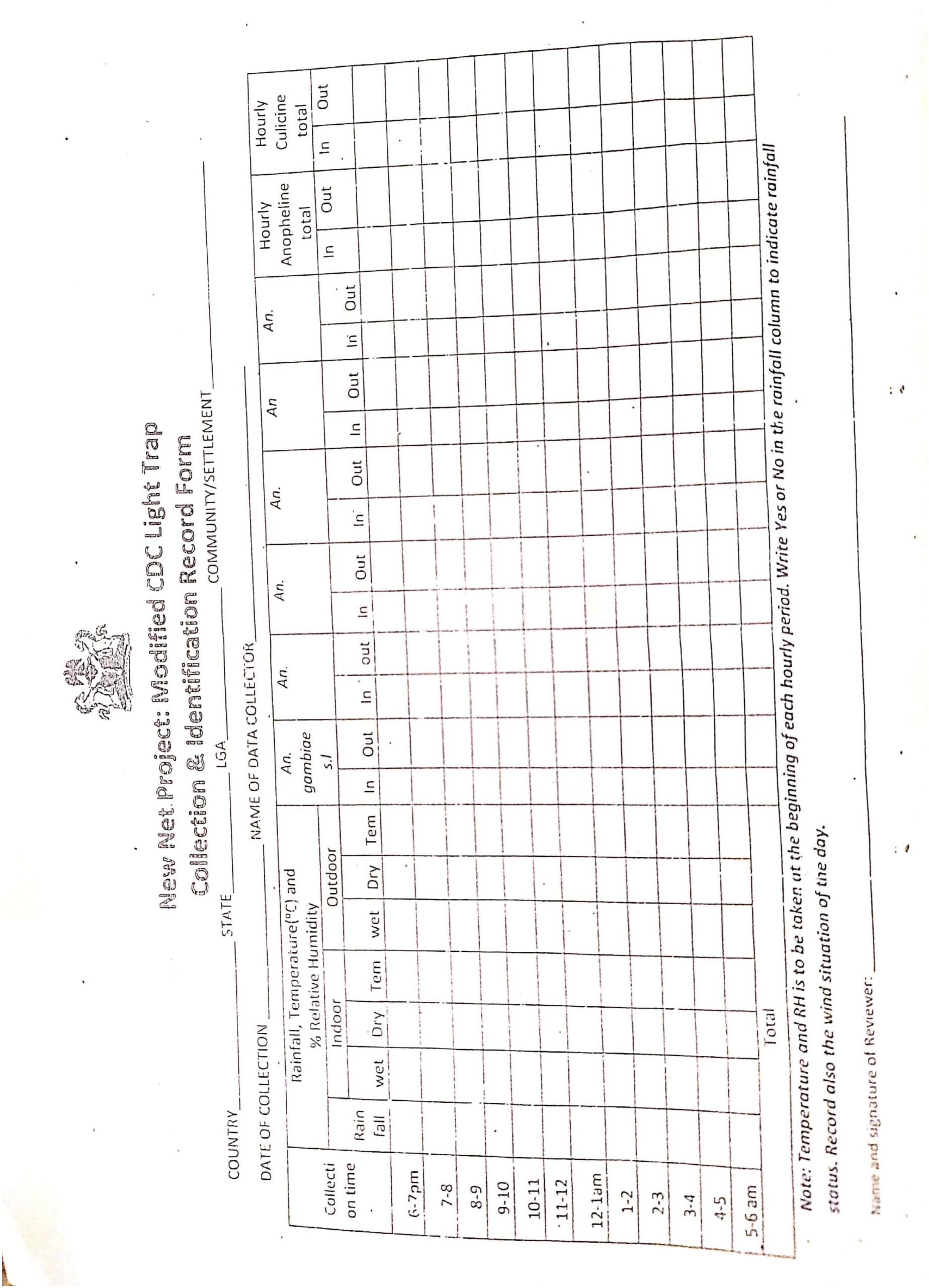

#### Sample pyrethrum spray catch and species identification record form

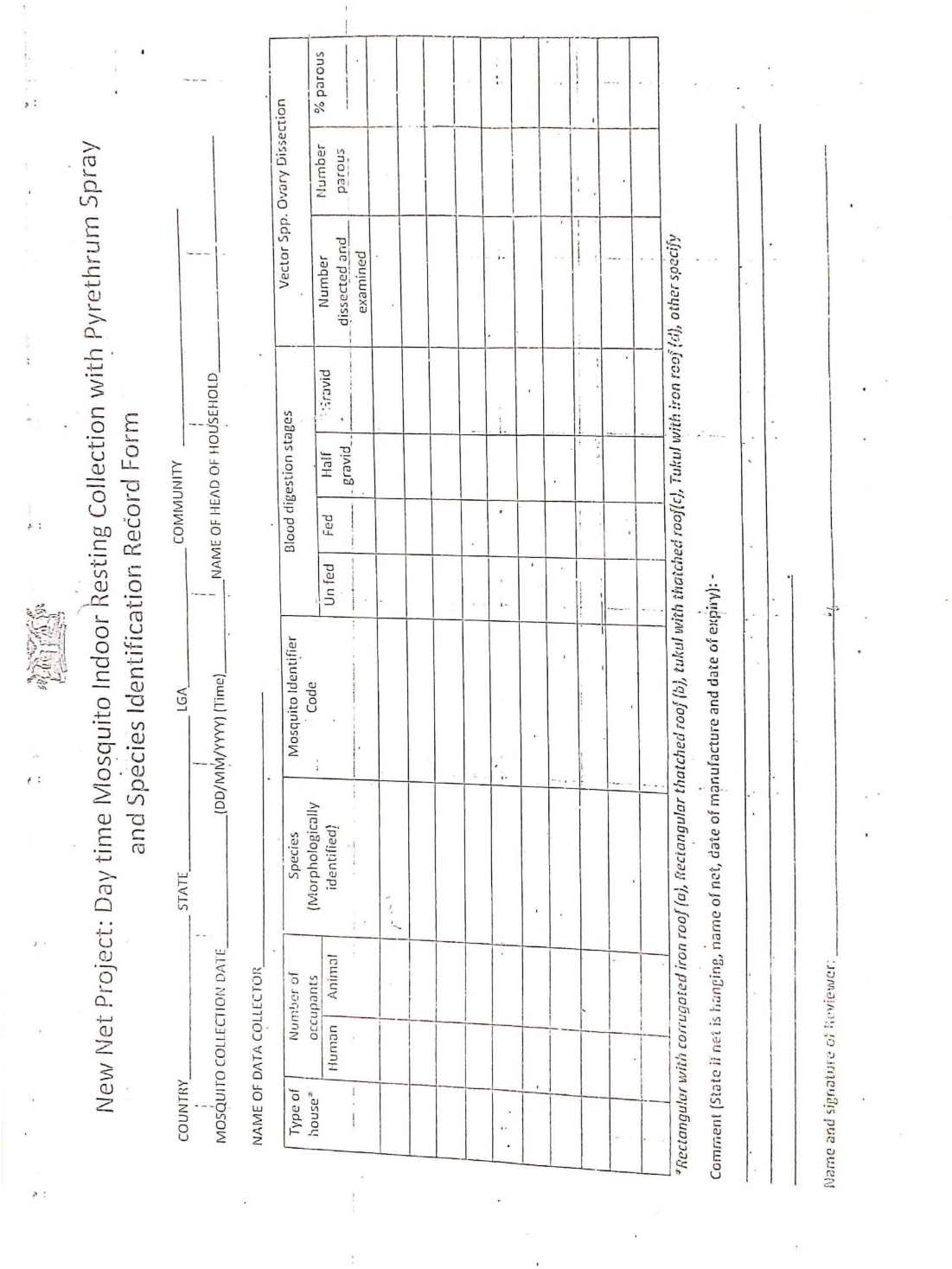

#### Sample morphology identification form

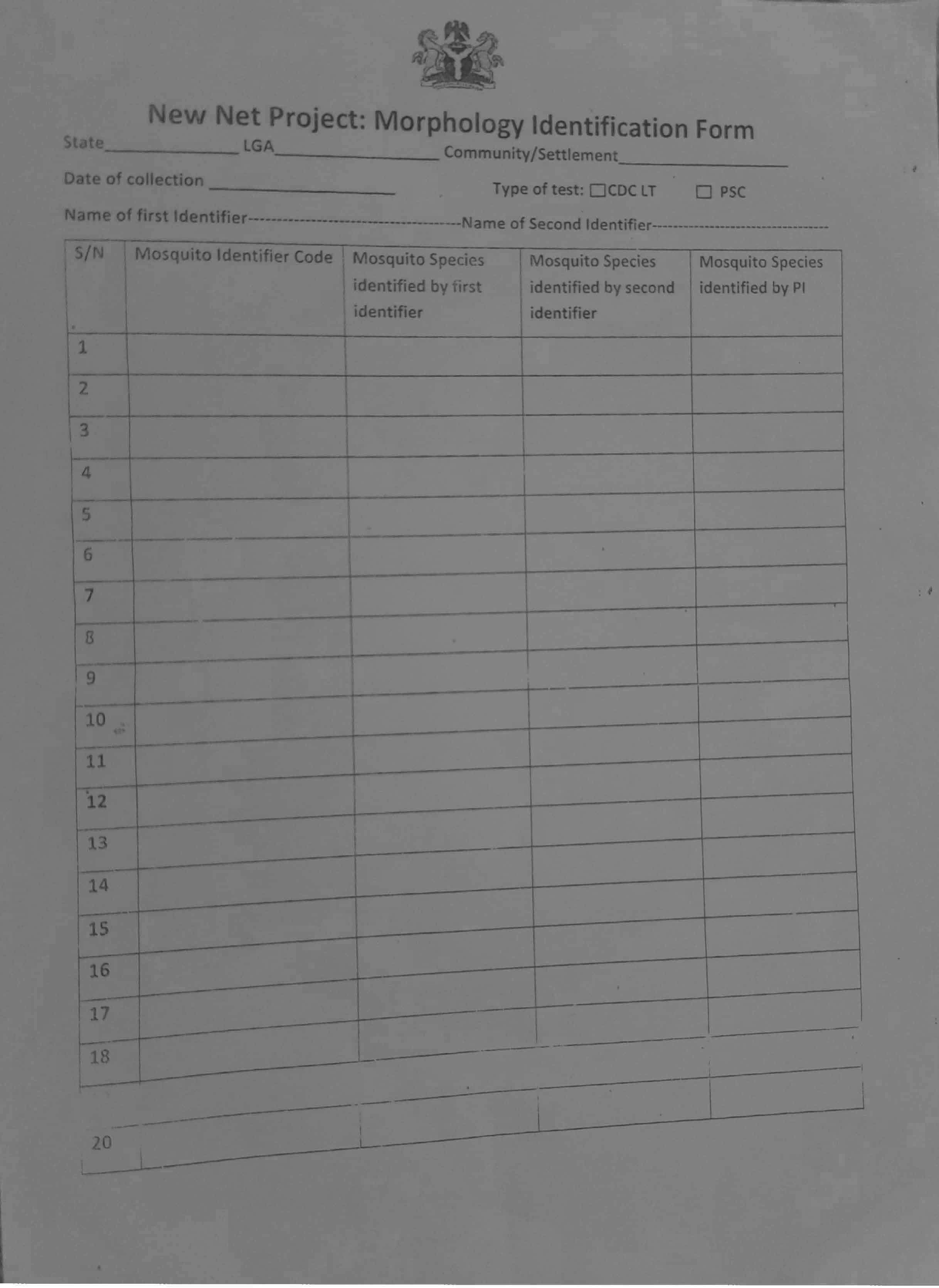

#### Entomological data collection protocol

##### A. Species identification of Anopheles gambiae complex

*DNA extraction*

The DNA will be extracted using Animal DNA Preparation Kit (Jena Bioscience, Germany), following the manufacturer’s guide.

- Place each mosquito sample in 1.5 ml microcentrifuge tube and homogenized in 300 μl Lysis Buffer and 2 μl of RNase A.
- Vortex vigorously for 30-60 seconds and add 8 μl of Proteinase K and mix by pipetting.
- Incubate homogenates at 60^o^C for 20 minutes in a water bath and cool for 5 minutes.
- Add 300 μl of Binding Buffer and vortex briefly.
- Place the tube on ice for 5 minutes and centrifuge at 10,000 rpm for 5 minutes to decant the supernatant.
- Insert a spin column in a 2 ml collection tube then add 100 μl Activation Buffer into the spin column.
- Centrifuge at 10,000 rpm for 30 seconds then the flow-through is discarded.
- Pipette directly into the spin column, centrifuge for 1 minute at 10,000 rpm and the flow-through discarded.
- Wash the DNA twice with 500 μl each of DNA wash buffer follow by centrifugation at 10,000 rpm for 30 seconds and a final centrifugation of the empty column for 2 minutes at 14,500 rpm.
- Elute the DNA in 40 μl of elution buffer and then store at 4^o^C.

*Preparation of PCR Mix*

The PCR mix is composed of a Master mix (dNTPs, Buffer, MgCl_2_ etc.), the primers (both forward and reverse), Taq polymerase (an enzyme), the DNA template to be amplified and finally a PCR grade water (double distilled water or **ddH_2_O**) to bring the volume to the desired volume. For the analysis, a 25 µl PCR mix is prepared as follows;

| **Reagents** | **Volume** |
| --- | --- |
| PCR Master Mix | 8µl |
| UN | 0.5 µl |
| AR | 0.5 µl |
| QD | 0.5 µl |
| GA | 0.5 µl |
| ME | 0.5 µl |
| Taq Polymerase | 0.25 µl |
| DNA Template | 2 µl |
| PCR Grade Water | 12.25 |
| **Total Volume** | **25 µl** |

**Steps in PCR Amplification**

| **S/N** | **Cycle step** | **Temperature** | **Time** | **Cycles** |
| --- | --- | --- | --- | --- |
| 1. | Initial Denaturation | 95°C | 2 minutes | 1 |
| 2. | Denaturation | 95°C | 30 seconds |  |
| 3. | Annealing | 55°C | 30 seconds | 30 cycles |
| 4. | Extension | 72°C | 40 seconds |  |
| 5. | Final Extension | 72°C | 7 minutes | 1 |
| 6. | Hold | 4°C | ∞ | -- |

*Gel Electrophoresis - Preparation and Casting of Agarose Gel for Electrophoresis*

Three percent 1.5 % agarose gel concentrations will be used for gel electrophoresis. The following procedure is required for preparation of the gel.

- The agarose will be melted in a microwave for about 2 mins until it has completely dissolved and allowed to cool sufficiently. Visualizing dye (5µl) (SYBR) will be added to ensure visualization of DNA.
- Upon cooling to about 50-45oC (not too hot to touch). The gel will be poured into a clean agarose well casting chamber in which a clean electrophoresis comb had been inserted as appropriate to create wells into which the amplicons would be loaded.
- After the gel had set, the cast will then be placed in a gel electrophoresis tank containing enough 1X TAE buffer to cover the gel and wells. The comb was then carefully removed from the gel in a way to avoid cracking of wells or gel.

*Loading of Amplicons and Running of the Agarose Gel Electrophoresis*

The samples will be loaded, and the gel electrophoresis run as described below:

- Molecular ladder (5µl) will be carefully dispensed into the first well of the gel or any designated well for ladder.
- 7.5µl of each amplicon (DNA) would be thereafter dispensed into corresponding wells.
- The electrophoresis tank covered with its lid and the tank cables will be appropriately connected to the electric source and set to run at 100 V for 1hour. After this, the gel is viewed to check for the bands.

*Identification of Anopheles coluzzii (Anopheles gambiae M form)*

It involves digestion of PCR product of *Anopheles gambiae s.s*with *Hha 1* Enzyme

*Procedure*

Prepare the mix in titre plate as follows:

- Incubate the mix at 37°C overnight
- Run 7μl of the digested PCR product 1.5% agarose gel containing and visualize in UV transillumination.
- 367bp and 23bp for *Anopheles colluzzii* (M form) and 257 and 110bp and 23bp for *Anopheles gambiae* S. form

| **Reagents** | **Volume** |
| --- | --- |
| Buffer + BSA  (1 x NE Buffer 4 and 1 x BSA) | 0.6µl |
| 1U of HhaI enzyme | 0.2 µl |
| PCR Grade Water | 1.0 |
| PCR product | 10 |
| **Total Volume** | **11.8 µl** |

**PRIMERS**

UN: GTG TGC CCC TTC CTC GAT GT

GA: CTG GTT TGG TCG GCA CGT TT

AR: AAG TGT CCT TCT CCA TCC TA

QD: CAG ACC AAG ATG GTT AGT AT

ME: TGA CCA ACC CAC TCC CTT GA

**Molecular weights of the bands**

*Anopheles merus* ………………………. .464 base pair

*Anopheles gambiae* ……………………	390 base pair

*Anopheles arabiensis*………………….……315 base pair

*Anopheles quadriannulatus*………………….153 base pair

***Anopheles Colluzzii* and An. gambiae S form**

367bp and 23bp for *Anopheles colluzzii* (M form) and 257 and 110bp and 23bp for *Anopheles gambiae* S. form

##### B. Molecular identification of Anopheles funestus complex

*DNA extraction*

Same as *Anopheles gambiae.*

*Preparation of PCR Mix*

Same as *Anopheles gambiae*

| **Reagents** | **Volume** |
| --- | --- |
| PCR Master Mix | 8µl |
| UV | 0.6 µl |
| FUN | 0.6 µl |
| VAN | 0.6 µl |
| RIV | 0.6 µl |
| PAR | 0.6 µl |
| LEES | 0.6 µl |
| Taq Polymerase | 0.25 µl |
| DNA Template | 2 µl |
| PCR Grade Water | 11.15 |
| **Total Volume** | **25 µl** |

**Steps in PCR Amplification**

| **S/N** | **Cycle step** | **Temperature** | **Time** | **Cycles** |
| --- | --- | --- | --- | --- |
| 1. | Initial Denaturation | 95°C | 2 minutes | 1 |
| 2. | Denaturation | 95°C | 30 seconds |  |
| 3. | Annealing | 47°C | 30 seconds | 35 cycles |
| 4. | Extension | 72°C | 40 seconds |  |
| 5. | Final Extension | 72°C | 5 minutes | 1 |
| 6. | Hold | 4°C | ∞ | -- |

*Gel Electrophoresis - Preparation and Casting of Agarose Gel for Electrophoresis*

Three percent 2.0 % agarose gel concentrations will be used for gel electrophoresis. The following procedure is required for preparation of the gel.

1. The agarose will be melted in a microwave for about 2 mins until it has completely dissolved and allowed to cool sufficiently. Visualizing dye (5µl) (SYBR) will be added to ensure visualization of DNA.
2. Upon cooling to about 50-45oC (not too hot to touch), the gel will be poured into a clean agarose well casting chamber in which a clean electrophoresis comb had been inserted as appropriate to create wells into which the amplicons would be loaded.
3. After the gel had set, the cast will then be placed in a gel electrophoresis tank containing enough 1X TAE buffer to cover the gel and wells. The comb was then carefully removed from the gel in a way to avoid cracking of wells or gel.

**Loading of Amplicons and Running of the Agarose Gel Electrophoresis**

Same as *Anopheles gambiae*

##### C. Detection of Plasmodium falciparum in mosquitoes

*PCR detection - Sample preparation*

- The sporozoite is found only at salivary gland between thorax and head
- Separate the abdomen from other part of the body
- Place the thorax-head on the tube for DNA extraction

*DNA extraction*

The DNA will be extracted using Animal DNA Preparation Kit (Jena Bioscience, Germany), following the manufacturer’s guide.

- Place each mosquito sample in 1.5 ml microcentrifuge tube and homogenized in 300 μl Lysis Buffer and 2 μl of RNase A.
- Vortex vigorously for 30-60 seconds and add 8 μl of Proteinase K and mix by pipetting.
- Incubate homogenates at 60^o^C for 20 minutes in a water bath and cool for 5 minutes.
- Add 300 μl of Binding Buffer and vortex briefly.
- Place the tube on ice for 5 minutes and centrifuge at 10,000 rpm for 5 minutes to decant the supernatant.
- Insert a spin column in a 2 ml collection tube then add 100 μl Activation Buffer into the spin column.
- Centrifuge at 10,000 rpm for 30 seconds then the flow-through is discarded.
- Pipette directly into the spin column, centrifuge for 1 minute at 10,000 rpm and the flow-through discarded.
- Wash the DNA twice with 500 μl each of DNA wash buffer follow by centrifugation at 10,000 rpm for 30 seconds and a final centrifugation of the empty column for 2 minutes at 14,500 rpm.
- Elute the DNA in 40 μl of elution buffer and then store at 4^o^C.

*PCR reaction*

Cyt b genes targeting P. falciparum is targeted (Hassan *et al*, 2009). Primers MitF2 (5'-TGAGTTATTGGGGTGCAACTG-3') and MitR2 (5'-TGTTTGCTTGGGAGCTGTAA-3').

*Preparation of PCR mix*

PCR mix for 25 µl PCR reaction:

| **Reagents** | **Volume** |
| --- | --- |
| PCR Master Mix | 8µl |
| MitF2 | 0.5 µl |
| MitR2 | 0.5µl |
| Taq Polymerase | 0.25 µl |
| DNA Template | 2 µl |
| PCR Grade Water | 13.75 |
| **Total Volume** | **25 µl** |

*Steps in PCR Amplification*

| **S/N** | **Cycle step** | **Temperature** | **Time** | **Cycles** |
| --- | --- | --- | --- | --- |
| 1. | Initial Denaturation | 95°C | 4 minutes | 1 |
| 2. | Denaturation | 95°C | 40 seconds |  |
| 3. | Annealing | 61°C | 40 seconds | 35 cycles |
| 4. | Extension | 72°C | 1 minute |  |
| 5. | Final Extension | 72°C | 10 minutes | 1 |
| 6. | Hold | 4°C | ∞ | -- |

*Gel electrophoresis - Preparation and Casting of Agarose Gel for Electrophoresis*

Three percent 2.0 % agarose gel concentrations will be used for gel electrophoresis. The following procedure is required for preparation of the gel.

4. The agarose will be melted in a microwave for about 2 mins until it has completely dissolved and allowed to cool sufficiently. Visualizing dye (5µl) (SYBR) will be added to ensure visualization of DNA.
5. Upon cooling to about 50-45oC (not too hot to touch), the gel will be poured into a clean agarose well casting chamber in which a clean electrophoresis comb had been inserted as appropriate to create wells into which the amplicons would be loaded.
6. After the gel had set, the cast will then be placed in a gel electrophoresis tank containing enough 1X TAE buffer to cover the gel and wells. The comb was then carefully removed from the gel in a way to avoid cracking of wells or gel.

*Loading of Amplicons and Running of the Agarose Gel Electrophoresis*

The samples will be loaded, and the gel electrophoresis run as described below:

1. Molecular ladder (5µl) will be carefully dispensed into the first well of the gel or any designated well for ladder.
2. 7.5µl of each amplicon (DNA) would be thereafter dispensed into corresponding wells.
3. The electrophoresis tank covered with its lid and the tank cables will be appropriately connected to the electric source and set to run at 100 V for 1hour. After this, the gel is viewed to check for the bands.

**Results**

Amplification bands: 729 bp fragment of the Cyt b gene.

##### D. PCR identification of Plasmodium spp from mosquito (Protocol 2)

*Procedure*

Prepare master mix for one 12.5 µl PCR reactions. Add reagents in the order presented

| **Reagents** | **X1 (**µ**l)** |
| --- | --- |
| ddH2O | 7.5 |
| Pre-mix (X5) with BSA | 2.5 |
| Pfr364F | 0.375 |
| Pfr364R | 0.375 |
| Pvr47F | 0.375 |
| Pvr47R | 0.375 |
| DNA | 1.0 |
| **TOTAL** | **12.5** |

*PCR cycle conditions*

- 95^o^C/5mins x 1 cycle
- (95^o^C/2mins, 56^o^C/30 sec, 72^o^C/30 sec) x 35 cycles
- 72^o^C/ 5mins x 1 cycle 4^o^C hold
- Run samples on a 2% agarose EtBr gel., load 10 µl sample.

*Base pairs*

- 700 or 220 bp for *P. falciparum*
- 333bp for *P. vivax*

*Primer sequences for multiplex PCR of Plasmodium vivax and falciparum*

| Pvr47F (F, 25pmol/l) | 5’ CTGATTTTCCGCGTAACAATG 3’ |
| --- | --- |
| Pvr47R (R, 25pmol/l) | 5’ CAAATGTAGCATAAAAATCCAAG 3’ |
| Pfr364F (F, 25pmol/l) | 5’ CCGGAAATTCGGGTTTTAGAC 3’ |
| Pfr364R (R, 25pmol/l) | 5’ GCTTTTGAAGTGCATGTGAATTGTGCAC 3’ |

(Oyedeji et al., 2007;Demas et al., 2011)

##### E. PCR identification of mammalian blood meals in mosquitoes by a multiplexed polymerase chain reaction

*DNA extraction* (deBenedictis, et al., 2003)

- Prepare extraction buffer by adding 0.1M NaCl, 0.2M sucrose, 0.1M Tris-HCl, 0.05M EDTA, pH 9.1 and 0.5% sodium dodecyl sulfate (SDS).
- Place mosquito in an autoclaved 1.5ml tube
- Homogenise mosquito sample in 100µL of extraction Buffer
- Incubate at 65^o^C for one hour.
- Precipitate the SDS by adding 15µL of cold 8M potassium acetate to each homogenate and incubate on ice for 45 minutes.
- Centrifuge for 10 minutes to remove cellular debris.
- Transfer supernatant into a new sterile 1.5 mL microfuge tube.
- Precipitate DNA by adding 250 µL of 100% ethanol to the transferred supernatant.
- Incubate for 5 minutes at room temperature.
- Spin for 15 minutes to pellet the DNA
- Discard the supernatant and dry
- Re-suspend dry pellets in 10 µL of 0.1 X SSC (15mM NaCl, 1.5mM sodium citrate) + 40 µL of double distilled water.

*Procedure*

Prepare master mix for one 12.5 µl PCR reactions. Add reagents in the order presented

| **Reagents** | **X1 (**µ**l)** |
| --- | --- |
| ddH2O | 6.75 |
| Pre-mix (X5) with BSA | 2.5 |
| Pig573F | 0.375 |
| Human741F | 0.375 |
| Goat894F | 0.375 |
| Dog368F | 0.375 |
| Cow121F | 0.375 |
| UNREV1025 | 0.375 |
| Template | 1.0 |
| **TOTAL** | **12.5** |

*PCR cycle conditions*

- 95^o^C/15mins x 1 cycle
- (95^o^C/1mins, 58^o^C/1 min, 72^o^C/I min) x 35 cycles
- 72^o^C/ 5mins x 1 cycle 4^o^C hold
- Run samples on a 2% agarose EtBr gel., load 10 µl sample.

*Base pairs*

- 453bp for pig
- 334bp for human
- 132bp for goat
- 680bp for Dog
- 561bp for Cow

*Primer sequences for the cytochrome b-based PCR blood meal identification assay*

Pig573F	5’ CCTCGCAGCCGTACATCTC 3’

Human 741F 5’ GGCTTACTTCTCTTCATTCTCTCCT 3’

Goat894F	5’ CCTAATCTTAGTACTTGTACCCTTCCTC 3’

Dog368F	5’ GGAATTGTACTATTATTCGCAACCAT 3’

Cow121F	5’ CATCGGCACAAATTTAGTCG 3’

UNREV1025 5’ GGTTGTCCTCCAATTCATGTTA 3’

##### F. Blood meal ELISA protocol

- Ensure that you have autoclaved enough tips, Eppendorf tubes and pestles
- Grind each mosquito with 100 µl of Phosphate Buffer Saline (PBS) in 1.5 ml eppendorf tube to prepare mosquito triturate
- Add 50 µl of mosquito triturate and the respective controls into wells of microtitre plate.
- (positive control: 10 µl of human serum+500 µl of PBS)
- Incubate for 1 hour
- Wash twice with 200 µl PBS – Tween 20. (To 1L of PBS, add 500 µl of Tween 20)
- Add 50 µl of prepared enzyme conjugate solution
- Affinity purified Antibody to human IgG (H+L); 1^o^ antibody 1:500
- Peroxidase-labeled affinity purified antibody to human IgG (H+L) 2^0^ antibody 1:500 i.e. add 1 µl of each to 500 µl of PBS
- Mix both primary and secondary antibody to get enzyme conjugate solution
- Incubate for 1 hour
- Wash three times with 200 µl PBS-Tween 20
- Add 100 µl of ABTS Peroxidase substrate
- Mix solution A and B together (i.e. 5ml+5ml per plate)
- Incubate for 30 minutes
- Wash three times with 200 µl PBS-Tween 20
- Add 50 µl of phosphatase substrate to each well
- Mix substrate A and B (i.e. 2.5ml+2.5ml per plate)
- Incubate for 30mins and read absorbance at 414 nm

*Preparation of Buffer - Preparation of Phosphate Buffer Saline (X1 PBS***)**

- To 800 ml of distilled water dissolve
  - 8 g of NaCl
  - 0.2 g of KCl
  - 1.44 g of Na_2_HPO_4_
  - 0.24 g of KH_2_PO_4_
- Adjust pH to 7.4 with HCl
- Adjust volume to 1 litre with distilled water

### Entomological parameters to be collected using study data

1. *The potential breeding sites of Anopheles mosquitoes*. Habitat occupancy will be computed as the number of aquatic habitats found to harbour Anopheles vector larvae or pupae divided by the number of potential habitats for Anopheles vector egg-laying and immature stage development in an area, by category of habitat.
2. The types of the *Anopheles* found, and nature of habitats utilized by each species in each site in both seasons.
3. *Larval/pupae density (per dip/per person/per time).* Larval density is the number of immature Anopheles vectors collected per dip, per person per unit time. Usually recorded by stage (I–IV instars and pupae) and by habitat and reported by stage category (early instar, late instar, pupae) for an area.
4. *Determination of indoor and outdoor man biting rates:* The man biting rate will be calculated for each species as HBR=N/HxNi where N=Number of female Anopheles mosquitoes, H= Number of persons that slept on the bed (covered with untreated bed net as a bait) at each collection point indoors or outdoors; Ni= Number of Nights.
5. *Calculation of Human Blood Index for indoor and outdoor mosquitoes*: The human blood index represents the total number of *Anopheles* mosquitoes collected indoors or outdoors with human blood divided by the total number of *Anopheles* mosquitoes with blood.
6. *Determination of Indoor Resting Density:* The indoor resting mosquito density will be calculated as the number of female mosquitoes collected for a species divided by the number of houses where PSC was conducted in the community*.*
7. *Calculation of Sporozoite rate for indoor and outdoor mosquitoes*: The *P. falciparum* sporozoite rate is the number of female *Anopheles* mosquitoes collected indoors or outdoors infected with sporozoites divided by the total number of mosquitoes examined for indoor or outdoor collection.
8. In each study location, *Anopheles* habitat will be geo-referenced and a habitat distribution map for each area will be created.
